# Multi-Omic Factor Analysis uncovers immunological signatures with pathophysiologic and clinical implications in coronary syndromes

**DOI:** 10.1101/2023.05.02.23289392

**Authors:** Kami Pekayvaz, Corinna Losert, Viktoria Knottenberg, Irene V. van Blokland, Roy Oelen, Hilde E. Groot, Jan Walter Benjamins, Sophia Brambs, Rainer Kaiser, Luke Eivers, Vivien Polewka, Raphael Escaig, Markus Joppich, Aleksandar Janjic, Oliver Popp, Tobias Petzold, Ralf Zimmer, Wolfgang Enard, Kathrin Saar, Philipp Mertins, Norbert Huebner, Pim van der Harst, Lude H. Franke, Monique G. P. van der Wijst, Steffen Massberg, Matthias Heinig, Leo Nicolai, Konstantin Stark

## Abstract

Acute and chronic coronary syndromes (ACS and CCS) are leading causes of mortality. Inflammation is considered to be a key pathogenic driver, but immune states in humans and their clinical implications remain poorly understood. We hypothesized that Multi-Omic blood analysis combined with Multi-Omic Factor Analysis (MOFA) might uncover hidden sources of variance providing pathophysiological insights linked to clinical needs. Here, we compile a single cell longitudinal dataset of the circulating immune states in ACS & CCS (13x10^3^ clinical & Multi-Omic variables, n=117 subjects, n=838 analyzed samples) from two independent cohorts. Using MOFA, we identify multilayered factors, characterized by distinct classical monocyte and CD4^+^ & CD8^+^ T cell states that explain a large proportion of inter-patient variance. Three factors either reflect disease course or predict outcome in coronary syndromes. The diagnostic performance of these factors reaches beyond established biomarkers highlighting the potential use of MOFA as a novel tool for multilayered patient risk stratification.

## Introduction

Myocardial ischemia is a major driver of mortality and morbidity worldwide^1^. This is caused by atherosclerosis in coronary arteries, which is clinically subdivided into stable chronic coronary syndromes (CCS) and acute coronary syndromes (ACS). Myocardial infarction (MI), the most severe form of ACS, is initiated by an acute disruption of blood flow to the myocardium due to plaque rupture in preexisting CCS^2^. Local and systemic immune responses are a main driver of atherosclerosis and contribute to thrombosis as well as myocardial remodeling after acute myocardial ischemia^3, 4^. However, despite extensive basic and translational research in the past decades, the immunological signatures in these disease entities in humans remain incompletely understood.

Single cell sequencing allows for characterizing immune signatures in an unbiased way with unprecedented resolution. In basic research it has been used to profile immune cells in atherosclerotic plaques^5^ and at sites of MI^6^. In addition, single cell genomics is increasingly taken up in clinical applications^7^. Its potential for diagnostics has been shown for systemic lupus^8^, while the prediction of survival^9^ and of treatment response have been demonstrated for immuno-therapy of different cancers^10–12^.

Systems biology and systems medicine approaches assessing different types of biomolecules simultaneously in multiple disease relevant tissues and cell types hold the potential to capture disease processes in their entirety^13^. Multi-Omics approaches are increasingly taken up in cardiovascular research^14^ and have led to the identification of predictive biomarkers for the stability of atherosclerotic plaques^5, 15^. Analysis of such Multi-Omics data is challenging due to the heterogeneity of data types and the large number of variables measured in relatively small numbers of samples. Multi-Omics factor analysis (MOFA) is an unsupervised approach for exploratory data analysis across multiple data types that enables the identification of the major axes of variation composed of multiple molecular features and to link these with the underlying molecular processes^16^. In contrast to analyzing the predictive potential of single variables, this data driven dimensionality reduction allows for testing single, integrative factors for their potential in diagnosis and prognosis, while retaining the wealth of information contained in the Multi-Omic dataset.

Together single cell and Multi-Omics systems-biological approaches have the potential to reveal the circulating immune response to ACS and CCS, which has not yet been characterized comprehensively in humans. This could provide mechanistic insights as a basis to develop new treatment strategies and provide non-invasive blood-based integrative biomarker signatures for the prediction of treatment outcome of ACS and for the non-invasive diagnosis of CCS.

We hypothesized that the concept of applying MOFA on patient blood samples might allow us to connect the immune signature of CCS and ACS to the diagnosis of coronary artery disease and to prediction of myocardial function after MI. Utilizing a multi-center, prospective, Multi-Omics strategy, we here characterize the circulating immune signatures and their time course in human coronary syndromes using single cell transcriptomics and patient level Multi-Omics in a clinically well-defined patient cohort with minimal co-morbidities. Most importantly, using the MOFA approach, we distil concise immune signatures with clinical and mechanistic implications. These immune signatures describe the disease course and are predictive for treatment outcome. Downstream analysis of the contributions of individual molecular features to each immune signature identified underlying molecular and cell-cell communication pathways. In summary, by linking our Multi-Omics study with MOFA, we unravel the clinical potential of immune signatures for diagnosis of coronary artery disease in CCS and prediction of the functional outcome after ACS in two independent, prospective cohorts.

## Results

### Multi-Omic characterization of the immune profile in acute coronary syndrome

We analyzed the human immune response to myocardial ischemia in a cohort of patients who presented at the cardiology department of the University Hospital, Ludwig-Maximilians-University (LMU) Munich (Munich cohort).

ACS patients all had a ST-elevation myocardial infarction (STEMI) and were only included in the absence of major comorbidities, cardiogenic shock or signs of infection at the time of inclusion (see inclusion and exclusion criteria in methods (Suppl. Table 1)). This cohort was sampled longitudinally to allow a high-resolution characterization of the systemic immune response to STEMI. We aimed to capture all major phases of the immune response during myocardial infarction^3, 4^ at four sampling timepoints in the Munich cohort (TPM): TP1M was the periinterventional timepoint, where sampling was performed during acute presentation at the catheter laboratory for immediate revascularization. TP2M was chosen to pick up the reperfusion injury, and sampling was performed 14 (± 8) h after re-opening of the culprit vessel. Samples at TP3M were collected 60 (± 12) h after the acute event as an intermediate timepoint. TP4M represented the normalization of creatine kinase (CK) values and the beginning of inflammation resolution and healing (prior to discharge, about 6.5 (±1.5) d after the acute event).

The Munich control cohort, without acute coronary ischemia, further allowed comparison between diagnostically secured CCS and CCS rule-out (non-CCS) patients. Samples from patients who presented with suspected coronary artery disease (CAD) were collected at timepoint TP0M before they underwent invasive or computed-tomography based coronary angiography and were subsequently classified as CCS or non-CCS (i.e., no evidence of CAD) (Suppl. Table 1).

Additionally, we validated the key immune signatures of our analysis in a second dataset from a cohort of ACS patients presenting at the University Medical Center Groningen (UMCG) (Groningen cohort). In the second independent dataset, the Groningen cohort^17^, sampling timepoints (TPG) for ACS were chosen in a similar manner: first admission to the catheter unit (TP1G), 24h after catheterization (TP2G), the last timepoint again represented the time when cardiac biomarkers had normalized, and the myocardial repair had been initiated (TP3G). In the Groningen cohort samples from this timepoint were collected 6-8 weeks after first admission.

In the Groningen control cohort healthy samples from the LifeLines DEEP^17^ cohort were included at a single timepoint TP0G. The detailed analysis and cohort description of the Groningen cohort are further described in the corresponding manuscript^17^ (Suppl. Table 2). Overall, we enrolled a combined total of 117 individual subjects and n=838 samples from clinical blood tests (n=125), scRNA Sequencing (scRNA-seq) (total: n=224 samples, derived from M: n=121; G: n=103), flow cytometry (n=122), cytokine multiplex-assay (n=127 samples), plasma proteomics (n=119), and neutrophil prime-Sequencing (prime-seq^18^) (n=121) (Fig. 1a,b, Suppl. Table 3,4).

**Fig. 1.**
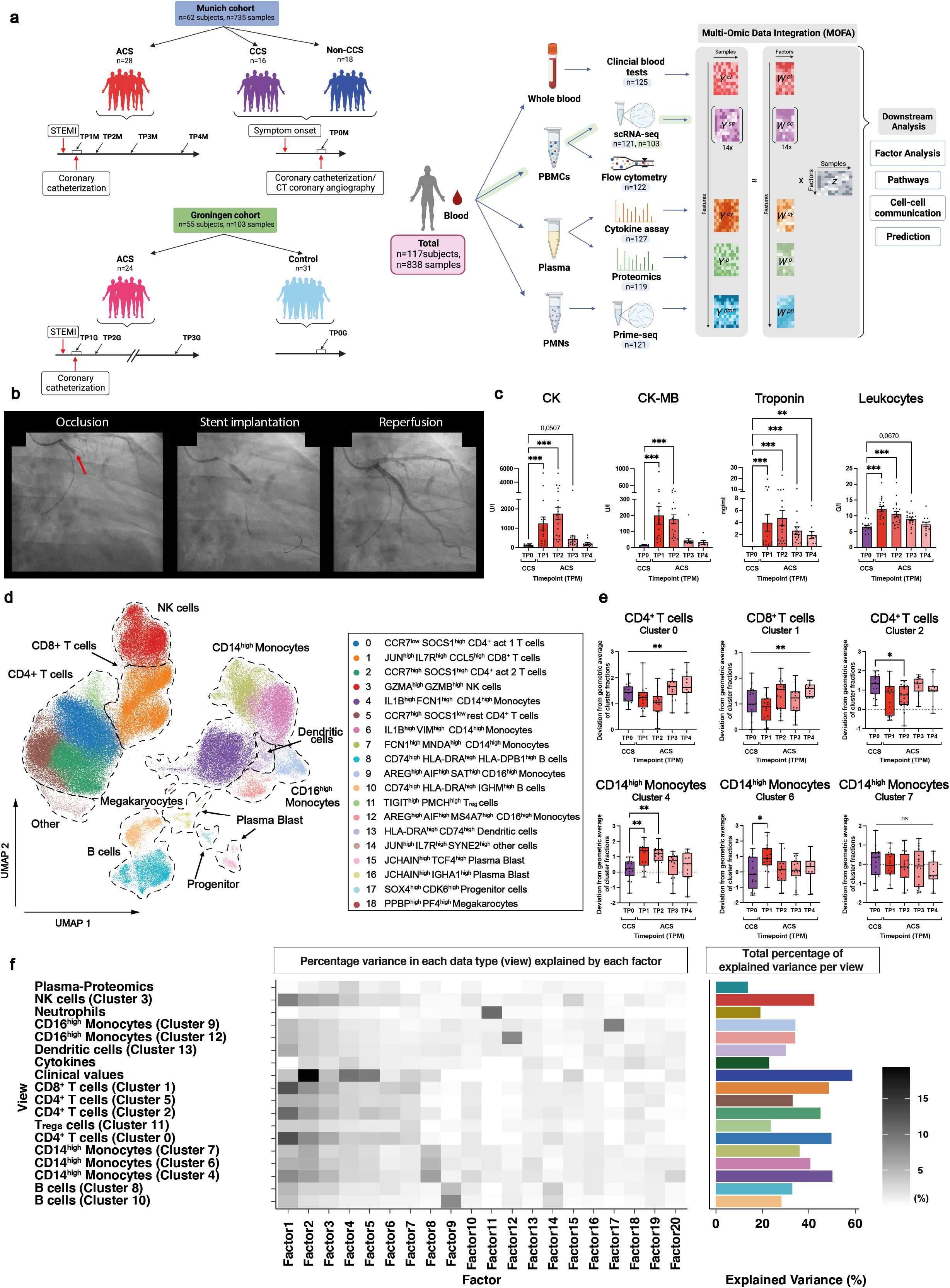
Study overview and longitudinal clinical and cellular characteristics of patients with myocardial ischemia. **(a)** Study design: In the Munich cohort, blood was analyzed longitudinally (TP1-4M) from patients with ACS as well as from patients with CCS and non-CCS (single sampling timepoint TP0M). A joint Multi-Omic dataset was created by using the following methods: clinical blood test (n=125), scRNA-seq (n=121), flow-cytometry (n=122), plasma proteomics (n=119), cytokine assay (n=127) and prime-seq (n=121). Samples were defined as independent Omic measures involving individual patients at different timepoints. This was followed by data integration, MOFA model estimation and subsequent downstream analysis such as factor analysis, pathways, cell-cell communication and prediction. In total, samples from n=62 patients were included in the Munich data cohort. Findings from the Munich data cohort were evaluated in the Groningen data (V2) as an independent validation cohort which measured blood from subjects (n=55) longitudinally (TP1-3G) as well as from a patient control group (TP0G). Samples were defined as independent Omic measures involving different timepoints or subjects. **(b)** Coronary catheterization with reperfusion of the occluded left circumflex artery (LCX) by stent implantation. **(c)** Clinical blood test. Individual timepoints of sterile ACS (TP1- 4M) compared to CCS patients (TP0M). CK (ACS: TP1M n=17, TP2M n=19, TP3M n=17, TP4M=12; CCS: TP0M n=13); CK-MB (ACS: TP1M n=16, TP2M n=19, TP3M n=16, TP4M=6; CCS: TP0M n=4); Troponin (ACS: TP1M n=17, TP2M n=19, TP3M n=16, TP4M=10; CCS: TP0M n=13); Leukocytes (ACS: TP1M n=17, TP2M n=19, TP3M n=17, TP4M n=12; CCS: TP0M n=15). Parametric distributed data were analyzed using the Ordinary One-Way ANOVA with correction for multiple comparisons by Dunnett’s test; non-parametric distributed data were analyzed using the Kruskal-Wallis test with correction for multiple comparisons by Dunn’s test. *p≤0.05, **p≤0.01, ***p≤0.001. Illustration of the mean value with SEM. **(d)** UMAP of scRNA-seq data from PBMCs showing cells of identified and annotated cell type clusters used for subsequent analyses (n=148.275)**. (e)** Analysis of centered log ratio (CLR) transformed cell type abundance based on scRNA-seq dataset. Individual timepoints of sterile ACS (TP1M n=16, TP2M n=19, TP3M n=16, TP4M n=11) compared to CCS patients (TP0M n=16). Parametric distributed data were analyzed using the Ordinary One-Way ANOVA with correction for multiple comparisons by Dunnett’s test; non-parametric distributed data were analyzed using the Kruskal-Wallis test with correction for multiple comparisons by Dunn’s test. *p≤0.05, **p≤0.01. In case only the column factor was significant, graphs are marked with a vertical bar on top. Illustration as a Box-Whiskers plot (minimum to maximum). **(f)** Variance decomposition showing the percentage of explained variance per view and factor of the MOFA model with 20 factors. Heatmap shows for each view the percentage of the variance of that view that is explained by the respective factor. Barplot (right) shows the total percentage of explained variance by all 20 factors.

We first focused on patients with ACS and a classical acute symptom onset, instant recanalization and no infectious complications during their disease course within the hospital (sterile ACS) and compared them to CCS patients (see Methods: Ethics & patient cohort). The analysis of laboratory values (CK, CK-MB isoenzyme, and Troponin T) confirmed the classical course of acute myocardial ischemia. In addition, transient elevations in C-Reactive Protein (CRP) and increased circulating leukocyte counts characterized a systemic immune response in MI (Fig.1c, Suppl. Fig. 1a). Flow cytometry-based phenotyping (n=122 samples) revealed no major quantitative changes in classical circulating leukocyte subsets in ACS compared to the CCS cohort (Suppl. Fig. 1b,c). To allow for a more refined resolution of leukocyte subsets, we made use of scRNA Sequencing (scRNA-seq).

Single-cell RNA sequencing revealed all major peripheral blood mononuclear immune cells (PBMCs) and rare populations such as plasma blasts, progenitor cells and circulating megakaryocytes^19^ (Fig. 1d, Suppl. Fig. 2a,b). We identified multiple previously described^20^ activated, resting and regulatory CD4^+^ T cell clusters. Compared to flow cytometry, the higher resolution of scRNA-seq-defined immune cell subsets revealed shifts in the composition of immune cell subsets across the disease course of MI (Fig. 1d-e, Suppl. Fig. 2a-c). Longitudinal analysis of centered log ratio (CLR) transformed cell type abundance identified marked shifts in the monocyte and T cell compartment during MI. CD14^high^ FCGR3A^low^ VCAN^high^ IL1B^high^ classical monocyte clusters 4 and 6 abundances increased early during acute infarction. Simultaneously, cluster 0 CD4^+^ act 1 (CCR7^low^ SOCS1^high^) T cell as well as cluster 2 CD4^+^ act 2 (CCR7^high^ SOCS1^high^) T cell abundance dropped during the disease course. In line, CD8^+^ T cell cluster 1 showed altered abundance across the immune response to ACS (Fig. 1e, Suppl. Fig. 2c). Overall, ACS is characterized by major, distinct changes in classical monocyte and T cell subsets compared to CCS.

### Multi-variate integration and factor analysis extract comprehensive immune signatures that explain inter-patient variance

To unlock the full potential and to identify overarching immune signatures of our Multi-Omic dataset from the Munich cohort, we integrated and harmonized data across the different types of data. We applied data type specific pre-processing (pseudo-bulk aggregation, library size normalization, log transformation, sample quantile normalization, filtering) and overall feature- quantile normalization (see methods). This resulted in 13,282 variable Multi-Omics and clinical features in the integrated dataset (Suppl. Fig. 3a). The complexity of the data, comprising different omics datasets, clinical variables and disease entities, made the analysis of the dataset with standard methods challenging and impractical. Therefore, we hypothesized that integrative factor analysis could exploit inter-patient variability to discover distinct immune signatures (i.e. multicellular programmes^21^) potentially allowing deduction of mechanistic and clinically relevant insights. Multi-Omic Factor analysis (MOFA) enables the identification of major axes of variance in a complex dataset and hence we used it to identify functionally relevant, coordinated immune responses^16^ that are not necessarily captured by individual features alone. MOFA moreover allows data integration across multiple data types (views). Here, views represent: (1) cell-type specific gene expression profiles in each of the circulating immune cell-type clusters identified in the single-cell RNA sequencing of PBMCs and (2) prime-seq^18^ data of neutrophils, (3) circulating proteins as determined by plasma proteomics or (4) cytokine analysis as well as (5) clinical laboratory markers. The cell type-specific gene expression profiles for the cell-type clusters in scRNA-seq data were defined by calculating the mean expression across all cells per individual and cell-type cluster. Hence, MOFA extracts shared as well as view-specific combinations of features that describe variation of the circulating immune response from single-cell and -patient level data (Fig. 1a).

We trained MOFA with 20 factors balancing the tradeoff between a high fraction of explained variance and interpretable factors. The analysis of the percentage of variance explained by each factor in each view showed that the inferred factors capture patterns that are shared across different views (Fig. 1f). Further, we analyzed the factor values at sample-level to identify whether MOFA factors capture patterns that distinguish the different disease entities and timepoints (Suppl. Fig. 3b). MOFA revealed several factors with diagnostic and mechanistic implications as further described in the subsequent paragraphs.

### A distinct integrative Factor represents the superordinate immune signature during myocardial infarction

MOFA identified factors that capture clinically relevant patterns: Factor 2 captured a large extent of inter-patient variance across the different views and explained most inter-patient variance in the clinical view (Fig. 2a, Suppl. Table 5). We first focused on patients with ACS and a classical acute symptom onset, instant recanalization and no infectious complications during their disease course within the hospital in comparison to stable CCS patients without an acute event. Factor 2 correlated with the development and resolution of myocardial ischemia reflected in the time course of clinical markers of myocardial damage and accurately discriminated chronic from acute coronary syndromes (Fig.1c, 2b). We therefore termed Factor 2 *Integrative ACS*. Next, we sought to replicate the temporal pattern of the immune response captured by *Integrative ACS* in the scRNA-seq data of the independent Groningen cohort, which was measured at comparable timepoints (Fig. 1a). After matching the cell type annotations of the two cohorts (Suppl. Fig. 4) and applying the same pre-processing and normalization approach in the Groningen cohort we computed *Integrative ACS* by applying the feature factor weights identified in the Munich cohort on cell type specific expression data in the Groningen cohort (see methods, Suppl. Fig. 5,6). *Integrative ACS* in the Groningen cohort indeed showed the same pattern across time (Fig. 2c) as *Integrative ACS* from the Munich cohort (Fig. 2b).

**Fig. 2.**
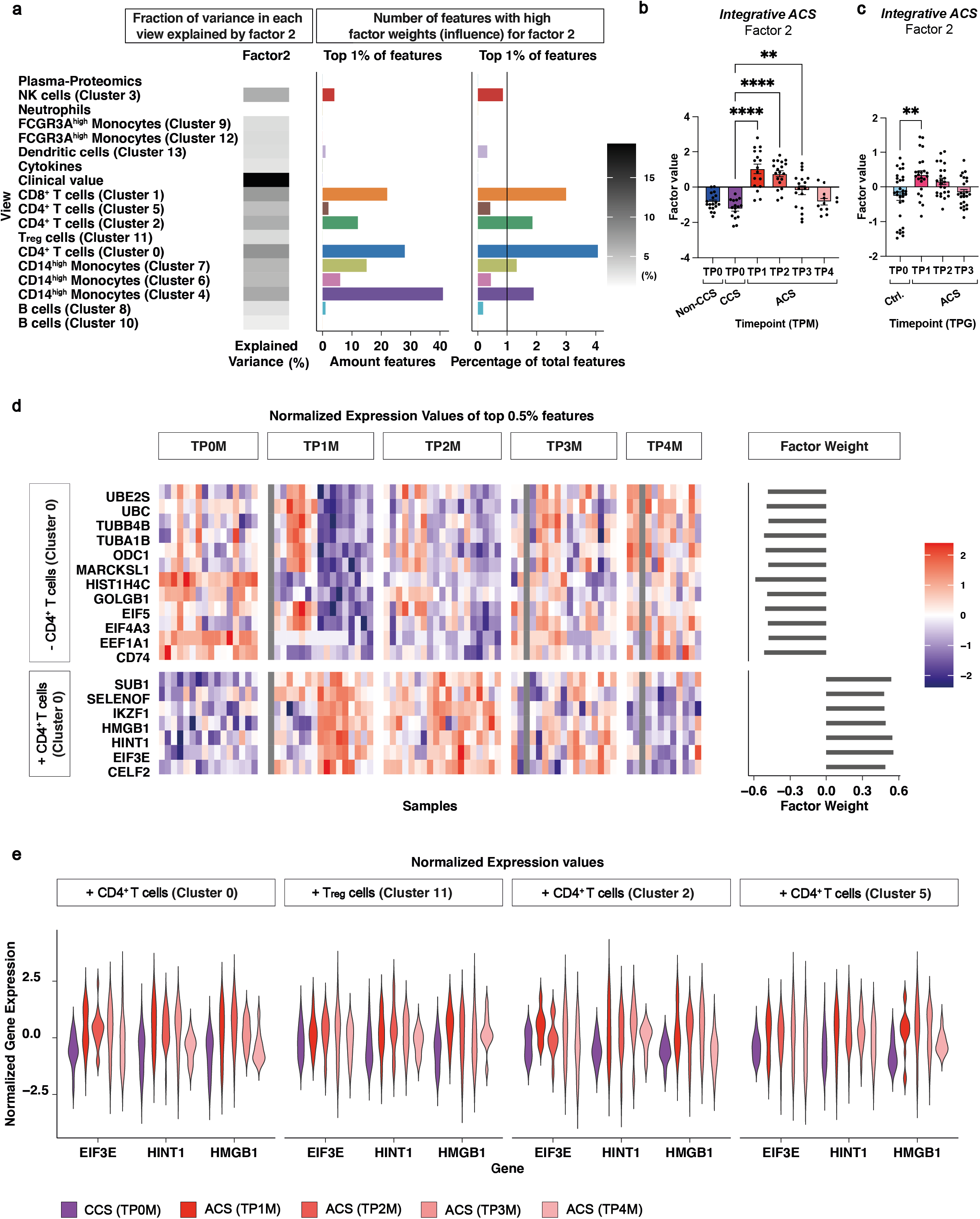
*Integrative ACS* defines the condensed immune signature of longitudinal myocardial infarction. **(a)** Overview of *Integrative ACS* (Factor 2): Heatmap shows for each view the percentage of the variance of that view that is explained by the factor. The barplots show the total amount (left) and the relative amount – in respect to the number of view specific features - (right) of features among the top 1% of highest-ranking features that influence the factor. **(b)** *Integrative ACS* (Factor 2). Comparison of the factor values of each timepoint of sterile ACS (TP1M n=17, TP2M n=19, TP3M n=17, TP4M n=12) with CCS patients (TP0M n=16) and non-CCS patients (TP0M n=18). Parametric distributed data were analyzed using the Ordinary One-Way ANOVA with correction for multiple comparisons by Dunnett’s test. *p≤0.05, **p≤0.01, ***p≤0.001. Illustration of the mean value with SEM. **(c)** Replication of *Integrative ACS* in Groningen cohort. Comparison of the factor values of each timepoint of ACS patients (TP1- 3G n=24) with control group (TP0G n=31) estimated based on top features of Integrative ACS. Non-parametric distributed data were analyzed using the Kruskal-Wallis test with correction for multiple comparisons by Dunn’s test. *p≤0.05, **p≤0.01. Illustration of the mean value with SEM. **(d)** *Integrative ACS* (Factor 2). Normalized expression values of top 0.5% features for CD4^+^ T cell Cluster 0 for sterile ACS (TP1M n=17, TP2M n=19, TP3M n=17, TP4M n=12) and CCS (TP0M n=16) patients in longitudinal comparison (heatmap) and weight of the features (barplot). (+ positive factor weight; - negative factor weight). **(e)** Normalized gene expression values of selected features for sterile ACS (TP1M n=16, TP2M n=19, TP3M n=16, TP4M n=11) and CCS (TP0M n=16) patients in longitudinal comparison. (+ positive factor weight; - negative factor weight).

Furthermore, we asked which specific views and single variables show strongest associations with *Integrative ACS*. Indeed, clinical markers of myocardial damage (Troponin T, CK and CK-MB) had high factor weights on *Integrative* ACS (Suppl. Table 6). However, the factor weight of the clinical variables was lower than that of many other Multi-Omics variables. Clinical markers were also not within the top 1% of variables with highest weights on Factor 2 (Fig. 2a, Suppl. Fig. 7). In addition, the temporal pattern of *Integrative ACS* could be reproduced after re-running MOFA factor analysis without any clinical variables (Suppl. Fig. 8, 9a,b), confirming that even though the factor captures a high amount of variance within the clinical markers and resembles the pattern of those, other Multi-Omic views are important to define *Integrative ACS*. We next asked what defines *Integrative ACS* to gain more profound insights into the nature of the immune response to myocardial ischemia.

We therefore focused on the molecular features contributing the highest factor weights to *Integrative ACS*. CD4^+^ act 1 (CCR7^low^ SOCS1^high^) cluster 0 T cell and CD4^+^ act 2 (CCR7^high^ SOCS1^high^) T cell cluster 2, as well as CD8^+^ T-cell cluster 1 and IL1B^high^ FCN1^high^ CD14^high^ monocyte cluster 4 had the largest relative amount of highly weighted features across views (Fig. 2a). Expression of EIF3E and HINT1 in CD4+ act 1 (CCR7^low^ SOCS1^high^) T-cells (Cluster 0) were among the highest-ranking features (Fig. 2d, Suppl. Table 6). In line, EIF3E is required for robust T-cell activation. Depending on CD28 coreceptor signaling, EIF3E regulates a burst in T-cell receptor signaling^22^. HINT1 complexed with HSP70 has been shown to hold strong immunomodulatory functions in NK cells^23^. HMGB1 across CD4^+^ and CD8^+^ T-cells similarly held a high factor weight in *Integrative ACS* (Fig. 2d, Suppl. Fig. 7a). HMGB1 promotes expansion and activation of T-cells and is a central alarmin for lymphoid cell fate and function^24^. As indicated by the strong association with the factor, the normalized expression of these genes showed a comparable development as *Integrative ACS* across the disease course (Fig. 2d,e).

Interestingly, T cell and monocyte clusters 0, 1 and 4, which contribute most features that constitute *Integrative ACS,* also showed significant alterations in circulating frequencies across disease evolution (Fig. 1e). CD4^+^ act 1 (CCR7^low^ SOCS1^high^) T cells (cluster 0) were defined as activated CD4^+^ T cells as described before^20^. Monocyte cluster 4 showed an IL1β^high^ CD14 ^high^ classical, pro-inflammatory phenotype (Fig. 1d, Suppl. Fig. 2a-b).

Hence, *Integrative ACS* forms a novel multilayered marker which largely explains inter-patient variance and defines the coordinative immune response to myocardial ischemia. *Integrative ACS* reflects the longitudinal pattern of myocardial ischemia and integrates clinical markers of myocardial damage with comprehensive Multi-Omic information.

### Distinct Interleukin signatures in monocytes and T cells characterize *Integrative ACS*

To allow for a more systematic understanding of characteristic immune signatures in *Integrative ACS* beyond the weight of individual features, we next sought to aggregate factor weights on the level of pathways. Across all our data views several immune system pathways of the REACTOME^25^ and KEGG^26^ database were enriched (Suppl. Table 7,8), including the Interleukin-6, -10, -12 & -27 signaling pathways which were mostly driven by high positive factor weights of genes of the CD4^+^ T cell and monocyte clusters as well as by some of the measured circulating cytokines (IL6, IL10, CXCL8) (Fig. 3a,b, Suppl. Fig. 9c,10). The enrichment of the IL6 cytokine signaling pathway was driven by high factor weights of *IL6ST*, *STAT3* and *JAK1* and *SOCS3* in cluster 2 and 5 CD4^+^ T cells and cluster 4 monocytes (Fig. 3b-d, Suppl. Fig. 11). In line, the circulating plasma cytokine IL6 itself also showed a considerable factor weight (Fig. 3c, Suppl. Table 6). Hence, the systemic immune response in ACS is dominated by a complex immune signature, characterized by an altered interleukin signaling in distinct monocyte and T cell clusters.

**Fig. 3.**
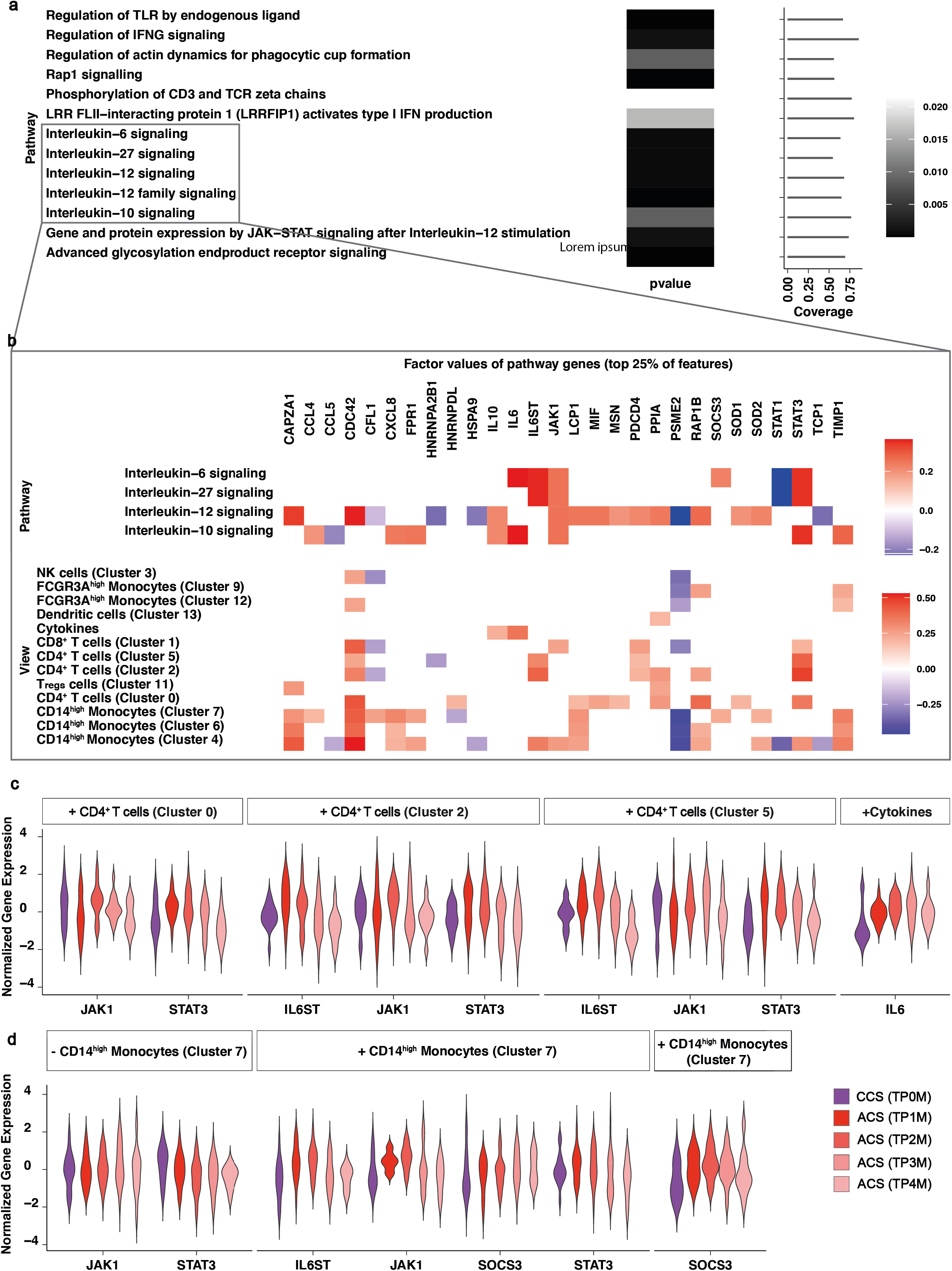
Integrative ACS is characterized by distinct interleukin signatures in monocytes and T cells. **(a)** Positively enriched REACTOME immune system pathways on *Integrative ACS* (Factor 2) across all data dimensions for which at least 50% of genes have been included within the feature set. FDR-adjusted p-value <0.05. **(b)** Factor weights of genes (top 25%) on *Integrative ACS* (Factor 2) belonging to enriched Interleukin pathways averaged across all dimensions and shown per dimension below. **(c)** Normalized gene expression values of genes belonging to Interleukin-6 signaling pathway for sterile ACS (Cluster 0,2,5: TP1M n=16, TP2M n=19, TP3M n=16, TP4M=11; Cytokines: TP1M n=17, TP2M n=19, TP3M n=16, TP4M=11) and CCS (TP0M n=16) patients in longitudinal comparison of CD4^+^ T cell clusters (cluster 0, 2 and 5) and Cytokine dimension. (+ positive factor weight). **(d)** Normalized gene expression values of genes belonging to Interleukin-6 signaling pathway for sterile ACS (TP1M n=16, TP2M n=19, TP3M n=16, TP4M n=11) and CCS (TP0M n=16) patients in longitudinal comparison of CD14^high^ monocytes clusters (cluster 4 and 7).

### Analysis of intercellular cross-talk unravels T cell-driven monocyte changes in ACS

Broad changes in cytokine profiles across different cell types in ACS may be coordinated by cell-cell communication. We therefore focused on identifying the underlying ligand-receptor pairs between immune cells and the downstream targets of cell surface receptors. For this we made use of the prior knowledge about potential ligand-receptor-target interactions provided by the NicheNet model^27^. To unravel cell-cell communication specific to *Integrative ACS*, we selected only features with the highest factor weights to define candidate target genes in the respective cell-types. We investigated ligands from other cell types, cytokines and proteins that show at the same time a high regulatory potential (given by the NicheNet model) and correlation of expression levels across samples with those target genes. Among the circulating cytokine features, this ligand-target analysis identified levels of circulating IL6, which also had a high factor weight on *Integrative ACS,* to associate with expression levels of Proto-oncogene serine/threonine-protein kinase Pim-1 in CD14^high^ cluster 4 monocytes (Fig. 4a-e). High IL6 levels correlated negatively with monocyte CD74 expression, involved in antigen presentation^28^, but were accompanied by increased VCAN expression in CD14^high^ cluster 7 monocytes (Fig. 4b-e). Summarized on a cellular level, most of the ligands whose expression showed high correlations with target genes with top factor weights of *Integrative ACS* were expressed in T-cell clusters. *Vice versa*, monocytes but also CD4^+^ and CD8^+^ T cells were the key receiver cells with expression levels of the largest number of targets correlating with T cell-derived ligands (Fig. 4a,b). The damage associated molecular pattern (DAMP) HMGB1, which also showed a high weight on *Integrative ACS*, was involved in a plethora of correlated ligand-target pairs. In detail, expression levels of the ligand HMGB1 in resting as well as activated CD4^+^ T cell clusters 0, 2, 5 CD4^+^ and cluster 1 CD8^+^ T cells resulted in high correlations with expression levels of the largest number of top-ranking target genes on *Integrative ACS* (Fig. 4a-c, Fig. 2d). HMGB1 was related to the reduction of proteasome activity within the receiver cell: expression of HMGB1 in Cluster 1 CD8^+^ T cells and cluster 0 CD4^+^ T cells showed a negative correlation with expression of UBC in cluster 0, 1 and 2 CD4^+^ T cells, suggesting a reduced ubiquitination potential^29^ (Fig. 4c-e, Fig. 2d). Ubiquitination in T cells is strongly linked to fine-tuning many immune responses, particularly positive or negative regulation of T cell activation^30^. Similarly, expression of HMGB1 in activated cluster 0 and cluster 2 CD4^+^ T cells correlated negatively with expression of Proteasome activator complex subunit 2 (PSME2) in cluster 4, 6, 7 CD14^high^ monocytes, emphasizing HMGB1-mediated inhibitory influences on proteasome activity (Fig. 4c-e, Fig. 2d). Besides that, TGFB1 expression in cluster 1 CD8^+^ T cells positively correlated with expression of ODC1 in cluster 4 CD14^high^ monocytes, known to inhibit inflammatory macrophage programs and macrophage apoptosis^31^, and therewith possible explaining the increased numbers of circulating cluster 4 monocytes during the disease course of ACS. ODC1 in these monocytes was downregulated early during ACS – however increased over time, suggesting an unleashed inflammatory monocyte state in early ACS and a gradual reduction of monocytic activation at later timepoints (Fig. 4 b-d). Hence, the communication between T cells and monocytes dominates the inflammatory response in ACS and is driven by the cytokine IL-6 and the alarmin HMGB1.

**Fig. 4.**
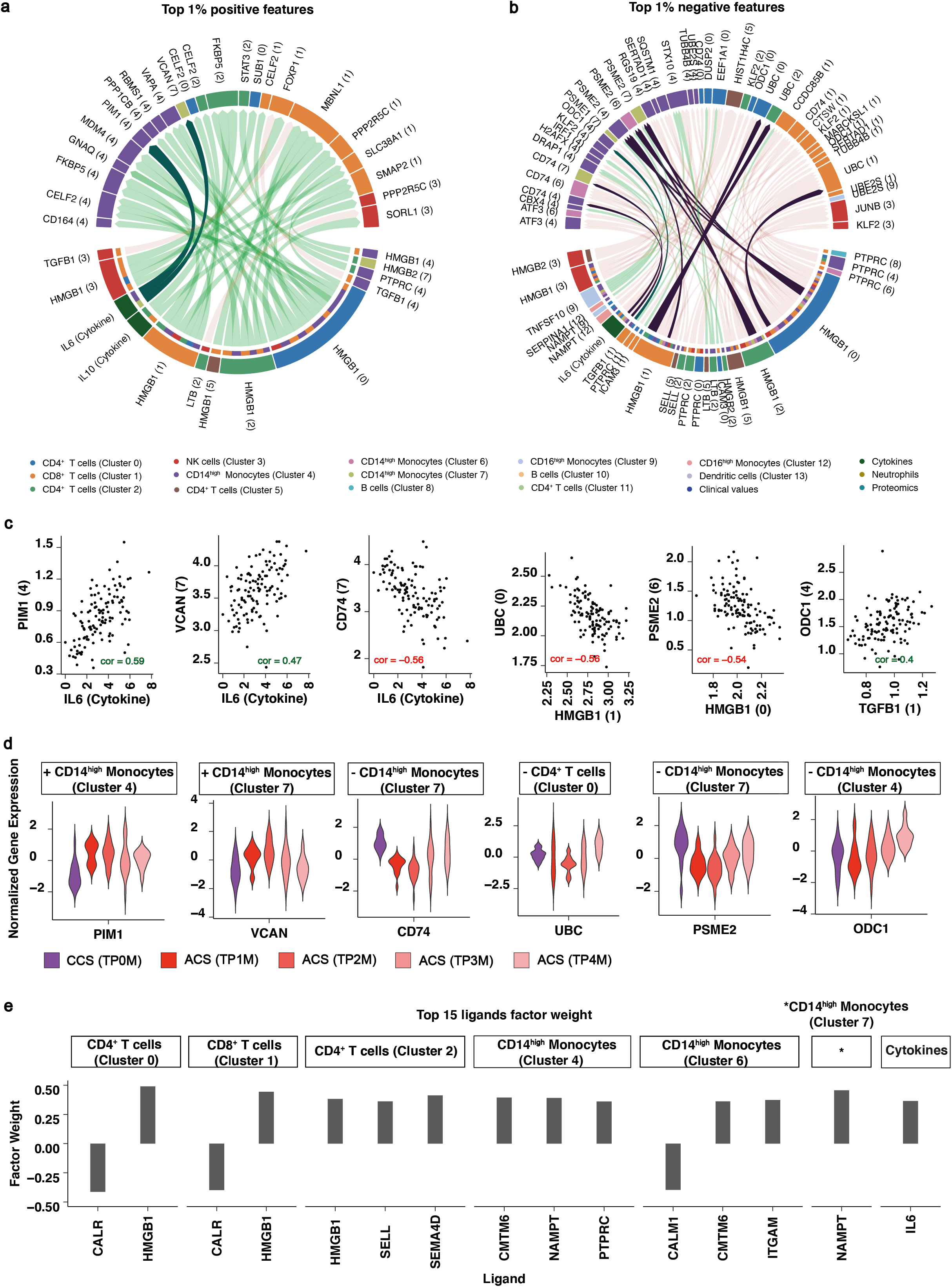
T cell mediated monocytic changes during ACS as unraveled by analysis of intercellular regulatory potential. **(a)** Spearman correlations (|cor| ≥0.4) between ligand and target genes across all samples (n=128). Target genes selected as top 1% of features with **positive** feature weight on *Integrative ACS.* Ligands selected based on minimum regulatory potential score of 0.0012 for those targets according to the NicheNet Model (corresponding to 97% quantile of regulatory potential score). **(b)** Spearman correlations (|cor| ≥0.4) between ligand and target genes across all samples (n=128). Target genes selected as top 1% of features with **negative** feature weight on *Integrative ACS*. Ligands selected based on minimum regulatory potential score of 0.0012 on those targets according to the NicheNet Model (corresponding to 97% quantile of regulatory potential score). **(c)** Spearman correlation scores of selected examples of circoplot (Fig. 4a,b). **(d)** Normalized gene expression values of selected features for sterile ACS (TP1M n=16, TP2M n=19, TP3M n=16, TP4M n=11) and CCS (TP0M n=16) patients in longitudinal comparison. **(e)** Factor weights of the top 15 ligands with the highest factor weight on *Integrative ACS* (Factor 2).

### Factor analysis distinguishes clinical ACS outcomes and predicts post-event development of cardiac dysfunction

We next hypothesized that differences in the time from symptom onset to presentation (i.e., time of myocardial ischemia) as well as different clinical courses of myocardial infarction (i.e., extent of myocardial damage) might be reflected in and influenced by variable immune responses in ACS. Therefore, we focused on ACS patients with different disease course: either with a symptom onset more than 24h ago (ACS with delayed recanalization after vessel occlusion), or patients who had experienced infectious complications (ACS acquiring hospital infection) and compared these to patients with uncomplicated ACS disease course with recent symptom onset (sterile ACS). Indeed, patients with an ACS with delayed recanalization after vessel occlusion and therefore a longer ischemia time showed a much stronger *Integrative ACS* response after recanalization than patients with direct presentation (Fig. 5a). In contrast, lower levels of cardiac biomarkers were observed in TP1M and TP2M in delayed coronary recanalization due to late patient presentation (Fig. 5b). Patients with an ACS that developed an infective complication during hospitalization showed increased *Integrative ACS* Factor values compared to patients without infective complications (Fig. 5a,c). Also, classical differential multiplex cytokine analyses revealed distinct differences in different clinical courses of ACS with Platelet-derived growth factor AB/BB levels being particularly elevated in patients with delayed recanalization and eminently low levels of circulating Interleukin-5 in patients with delayed recanalization or in ACS patients acquiring hospital infections (Suppl. Fig. 12,13a). Mechanistic and translational studies during the last decades suggested a possible impact of different immune states on a favorable or detrimental development of myocardial healing and myocardial function^3, 32, 33^. We therefore asked whether integrative MOFA factors might allow prediction of the extent of myocardial recovery at early sampling time points. This could serve to identify patients at risk for the development of ischemic heart failure and possibly trigger early initiation of heart failure treatment, more extensive revascularization, or prolonged monitoring on the ICU. To define patients with favorable or adverse developments of cardiac function, we measured baseline left ventricular ejection fraction (EF) during the acute phase of myocardial infarction in the intensive care unit and again before discharge by echocardiography and divided patients into two groups. Patients who showed a drop in left ventricular ejection fraction across the disease course were considered to have an adverse development of cardiac function (poor outcome) in contrast to a favorable development (good outcome) (Fig. 5d). We hypothesized that these variable disease courses might be associated and/or influenced by different immune signatures, either supporting cardiac healing or leading to detrimental inflammation causing adverse cardiac remodeling. We asked whether the MOFA factors could predict an adverse development of cardiac function. Therefore, we enrolled patients with ACS with acute symptom onset and compared a favorable outcome (improved or stable ejection fraction across the hospitalized disease course) with a non-favorable outcome (worsened ejection fraction across the hospitalized disease course). Factor 4 had particularly low levels already at the time of hospital admission in patients with a poor outcome compared to patients with improved cardiac function (Fig. 5e). In contrast, clinical laboratory markers of myocardial damage showed a non-significant trend towards increased CK levels across the disease course. However, no difference was observable at early timepoints, where predictive markers are needed (Fig. 5f,g). This stresses the utility of novel integrative factors for outcome prediction and emphasizes the relevance of the altered immune signature in early myocardial damage for the functional cardiac outcome. We evaluated the potential of Factor 4 values estimated at TP1M to predict good vs. poor outcome using the area under the curve (AUC) of the receiver operating characteristic curve (ROC). Prediction with Factor 4 outperformed the prediction based on established clinical markers at TP1M (Fig. 5g) even when taking into account the complete time course of those markers (Suppl. Fig. 13b, Suppl. Table 9). Due to the unsupervised training of the model, Factor 4 was estimated independently of outcome. Additionally, no information about the trajectory of specific patients (coupling of timepoints) was used during model estimation. Thus, these results are indicative of the potential translational value of Factor 4. We termed this Factor 4 *Integrative ACS Outcome*.

**Fig. 5.**
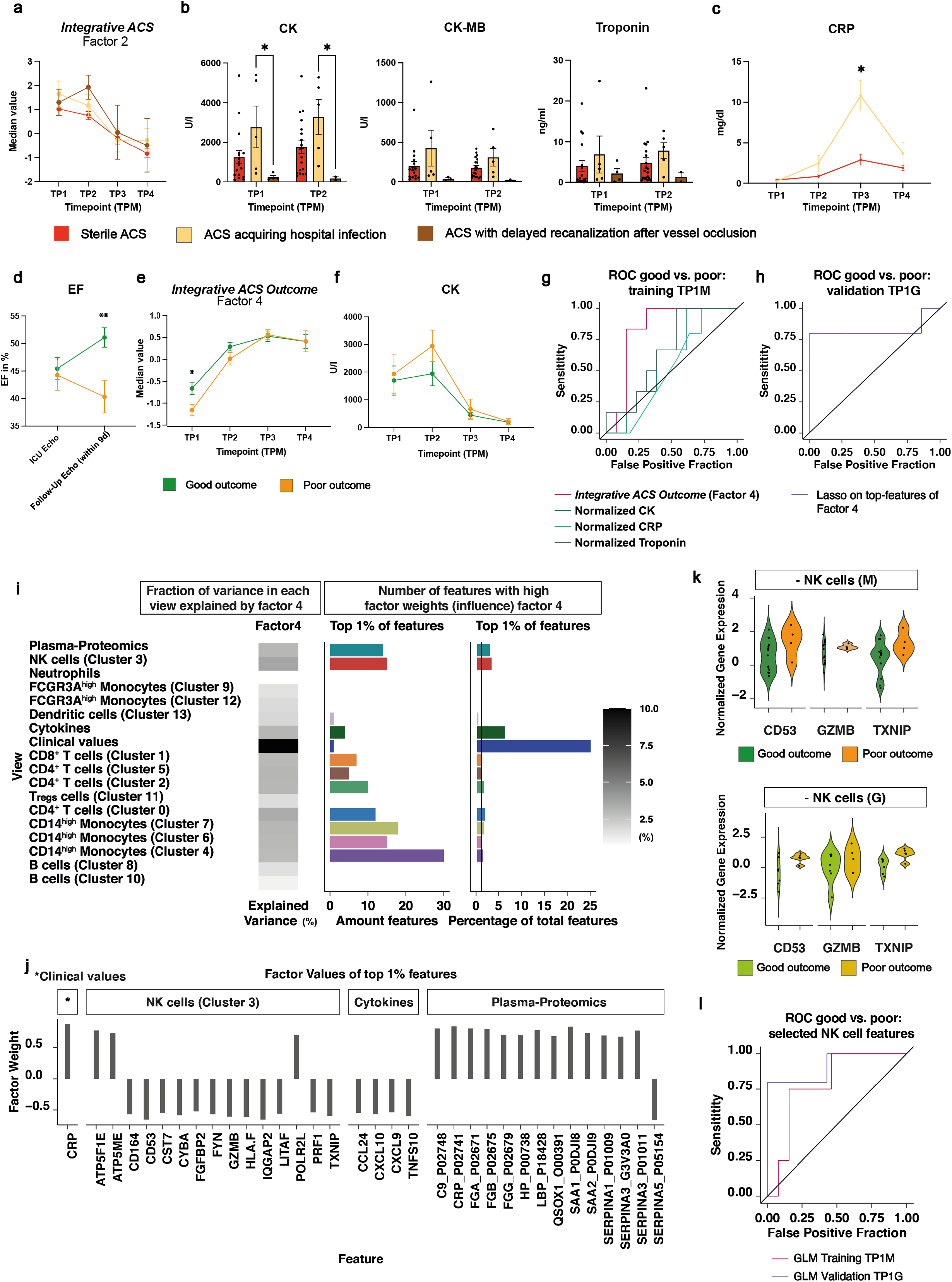
Factor analysis identifies distinct immune signatures in ACS subtypes. **(a)** *Integrative ACS* (Factor 2). Longitudinal comparison of factor values between ACS subtypes (sterile ACS TP1M n=17, TP2M n=19, TP3M n=17, TP4M n=12; ACS acquiring hospital infection TP1M n=5, TP2M n=5, TP3M n=5, TP4M n=4; ACS with delayed recanalization after vessel occlusion TP1M n=4, TP2M=2, TP3M n=2, TP4M n=2) **(b)** Comparison between the early timepoints (TP1-2M) of ACS subtypes (sterile ACS, ACS acquiring hospital infection and ACS with delayed recanalization after vessel occlusion). CK (sterile ACS TP1M n=17, TP2M n=19; infectious ACS TP1M n=5, TP2M n=5; ACS with delayed recanalization after vessel occlusion TP1M n=4, TP2M=2); CK-MB (sterile ACS TP1M n=16, TP2M n=19; ACS acquiring hospital infection TP1M n=5, TP2M n=5; ACS with delayed recanalization after vessel occlusion TP1M n=3, TP2M=1); Troponin (sterile ACS TP1M n=17, TP2M n=19; ACS acquiring hospital infection ACS TP1M n=5, TP2M n=5; ACS with delayed recanalization after vessel occlusion TP1M n=4, TP2M n=2). Dataset was analyzed using the Mixed-effects analysis with correction for multiple comparisons by Tukey’s test. *p≤0.05. **(c)** Longitudinal comparison of CRP between sterile ACS and ACS acquiring hospital infection patients (sterile ACS TP1M n=17, TP2M n=18, TP3M n=17, TP4M n=12; ACS acquiring hospital infection TP1M n=5, TP2M n=5, TP3M n=5, TP4M n=4). The dataset was analyzed using the Mixed-effects analysis with correction for multiple comparisons by Šidák’s test. *p≤0.05. Illustration of the mean value with SEM. **(d)** Ejection fraction (EF) comparing good (n=14) and poor (n=7) outcome during hospitalization. The parametric dataset was analyzed using an unpaired t-test. *p≤0.05, **p≤0.01. Illustration of the mean value with SEM. **(e)** *Integrative ACS Outcome* (Factor 4). Longitudinal comparison of factor values between good and poor outcome patients (good outcome: TP1M n=13, TP2M n=14, TP3M n=14, TP4M=11; poor outcome: TP1M n=6, TP2M n=7, TP3M n=7, TP4M n=4). The parametric dataset was analyzed using an unpaired t-test. *p≤0.05. Illustration of the mean value with SEM. **(f)** Longitudinal comparison of CK between patients with good and poor outcome (good outcome: TP1M n=13, TP2M n=14, TP3M n=14, TP4M=11; poor outcome: TP1M n=6, TP2M n=7, TP3M n=7, TP4M n=4). The dataset was analyzed using the Mixed-effects analysis with correction for multiple comparisons by Šidák’s test. *p≤0.05, **p≤0.01. Illustration of the mean value with SEM. **(g)** ROC AUC. Comparison of the predictive power of *Integrative ACS Outcome* (Factor 4) (n=19) and nominalized CK levels (n=19), normalized CRP levels (n=16) and normalized Troponin levels (n=19) collected at TP1M. **(h)** ROC AUC. Evaluation of prediction potential in Groningen cohort. ROC curve of lasso model trained on top features of *Integrative ACS Outcome* at TP1M and applied to Groningen cohort to predict the outcome (TP1G good outcome: n=7; poor outcome n = 5) **(i)** Overview of *Integrative ACS Outcome* (Factor4): Heatmap shows for each view the percentage of the variance of that view that is explained by the factor. The barplots show the total amount (left) and the relative amount – in respect to the number of view specific features - (right) of features among the top 1% of highest-ranking features that influence the factor. **(j)** Factor values of top 1% features showing only features belonging to NK cell cluster 3, Clinical, Plasma-Proteomics and Cytokine data dimension on *Integrative ACS Outcome* (Factor 4). **(k)** Normalized gene expression values of selected top features from NK cells in Munich cohort and Groningen cohort comparing patients with good (Munich: n=13, Groningen: n=7) and poor (Munich: n=4, Groningen: n=5) outcome at TP1. **(l)** ROC AUC. Prediction results of logistic regression model trained on selected NK features (CD53, GZMB, TXNIP) on the Munich dataset as training dataset and applied to the Groningen dataset as holdout validation dataset.

To corroborate the predictive value of the top-ranking features included in *Integrative ACS Outcome* already at the earliest timepoint, independent of their subsequent changes in expression at later timepoints, we trained a penalized logistic regression model that predicts outcome from these features at TP1M on the Munich cohort. We then applied this model to predict the outcome for the patients of the Groningen replication cohort at TP1G with available outcome data (see methods). Comparison of the predicted and observed outcomes resulted in ROC AUC of 0.83 on the replication cohort (Fig. 5h, Suppl. Table 9), thus demonstrating the ability of the model to generalize beyond the Munich cohort and differentiating good and poor outcome patients already at an early stage.

Next, we investigated which features mainly define this protective integrative factor. We assumed, that features with high positive factor weights on *Integrative ACS Outcome* define rather protective immune features, whereas features with high negative factor weights define rather detrimental immune features (Suppl. Fig.14). Interestingly, a high number of features indicating a distinct NK cell phenotype showed high negative weights on Factor 4 (Fig. 5i,j). This signature correlating with a negative development of cardiac function at later timepoints was characterized by NK cell activation and cytotoxicity as shown by high factor weights of *TXNIP*^34^, *PRF1*^35^, *LITAF*^36^, *GZMB*^35^, *FYN*^37^, *CST7*^38^ and *CD53*^39^ expression (Fig. 5j, Suppl. Fig. 14a). Also, anti-angiogenic mediators such as CXCL9 and CXCL10^40^ or TNFS10 (TRAIL)^41^ derived from the multiplex cytokine view had high negative factor weights on *Integrative ACS Outcome* (Fig. 5j, Suppl. Fig. 14-16). On the other hand, the plasma proteomics view revealed that circulating anti-trypsin enzymes of the SERPIN family (SERPINA1, SERPINA2, SERPINA3), known to protect cells from granzyme-mediated cytotoxicity^42^, showed high positive factor weights on *Integrative ACS Outcome* and hence associate with a more protective immune state. Interestingly, the degree of inflammation *per se* was not necessarily associated with an adverse outcome since circulating proinflammatory proteins such as CRP, SAA1, SAA2 and C9 showed high positive weights on the protective factor *Integrative ACS Outcome* (Fig. 5j).

Besides the analysis of high-ranking features characterizing *Integrative ACS Outcome,* we investigated their normalized expression values at TP1M and subsequent timepoints distinguishing good and poor outcome samples (Suppl. Fig. 17) focusing on their potential to differentiate outcome already at an early stage (TP1M). We observed differential expression of CD53, GZMB and TXNIP in NK cells between good and poor outcome groups at TP1M (Fig. 5k). We further evaluated whether these selected features at TP1M predict the outcome of those patients. A logistic regression model including only these features yielded a ROC AUC value of 0.79 (Fig. 5l, Suppl. Table 9) on the Munich cohort as training dataset. To evaluate its generalizability to other datasets, we applied it to the Groningen validation cohort, yielding a ROC AUC of 0.91 (Fig. 5l, Suppl. Table 9). In line with that we also find differential expression of those features between good and poor outcome at TP1G within the Groningen cohort (Fig. 5k). This highlights the potential of identifying a small subset of predictive features among top-ranking *Integrative Outcome* features that might serve as targeted clinical markers for outcome prediction without the need of collecting a Multi-Omics dataset.

### *Integrative CCS* robustly identifies the presence of coronary artery disease

Next, we tested whether MOFA factors can identify CCS patients with established CAD from patients with a CAD rule-out (non-CCS), which would provide a non-invasive diagnostic approach. The samples of patients with suspected coronary artery disease were sampled in a prospective manner without prior information on coronary artery status. The different groups (CCS, non-CCS) showed comparable cardiovascular risk factors and baseline characteristics (compare Suppl. Table 1). Factor 1 showed high positive values in patients with chronic coronary syndromes (Fig. 6a). In contrast, patients with healthy coronaries but also patients with acute coronary syndromes showed lower and mainly negative values of Factor 1 (Fig. 6a). We hence termed this factor *Integrative CCS*. In detail, 94% of patients with CAD showed an *Integrative CCS* value above 0, whereas 91% of patients with exclusion of CAD showed an *Integrative CCS* value below 0. Patients with coronary artery sclerosis (evidence of atherosclerosis with vessel narrowing <50%) showed intermediate values of *Integrative CCS* (Fig. 6a). We evaluated the potential of the *Integrative CCS* value to predict CCS patients against patients without CCS. When also including patients with non-occlusive coronary sclerosis in the non-CCS group, *Integrative CCS* outperformed the established “SCORE2”^43^ CVD risk score (ROC AUC 0.84 vs 0.63). Considering only patients without CCS or patients with established CCS (hence excluding coronary sclerosis), *Integrative CCS* again outperformed “SCORE2” (ROC AUC 0.99 vs 0.80, Fig. 6b, Suppl. Table 10)^43^.

**Fig. 6.**
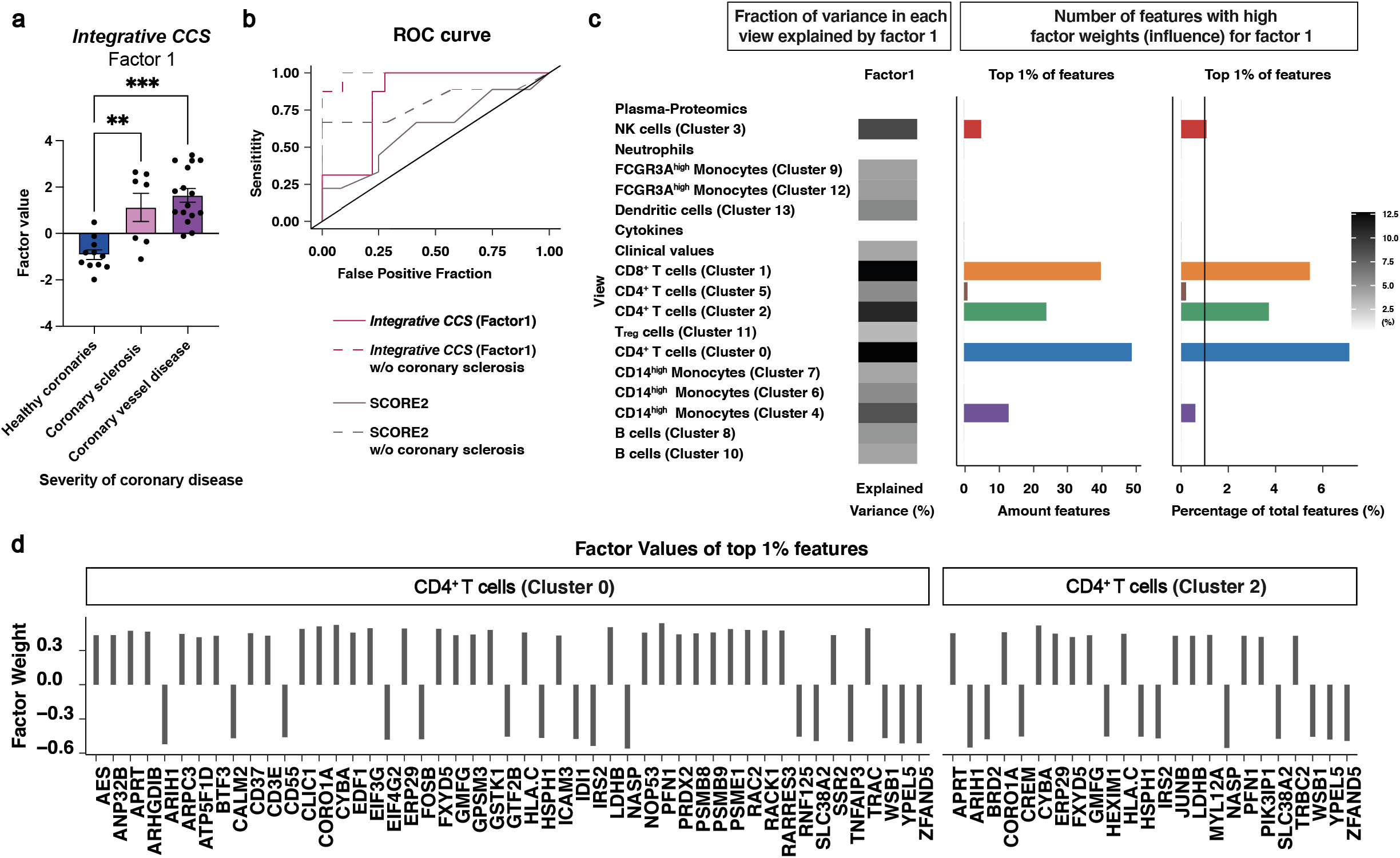
CCS patients can be robustly identified prior to invasive diagnostics using *Integrative CCS*. **(a)** *Integrative CCS* (Factor 1). Comparison of factor values for patients with CCS (TP0M n=16), coronary sclerosis (TP0M n=7) and healthy coronaries (TP0M n=11). Parametric distributed data were analyzed using the Ordinary One-Way ANOVA with correction for multiple comparisons by Tukey’s test. *p≤0.05, **p≤0.01, ***p≤0.001. Illustration of the mean value with SEM. **(b)** ROC AUC. Comparison of the predictive power of *Integrative CCS* (Factor 1) (n=34) and SCORE2 (n=21) on the risk of a cardiovascular event. The dotted line shows the ROC AUC of Integrative CCS (n=21) and SCORE2 (n=16) without including patients with coronary sclerosis. **(c)** Overview of *Integrative CCS* (Factor 1): heatmap shows for each view the percentage of the variance of that view that is explained by the factor and the barplots the total amount (left) and relative amount – in respect to the number of view specific features - (right) of features among the top 1% of highest-ranking features on the factor. **(d)** Factor values of top 1% features showing only features belonging to CD4^+^ T cells cluster 0 and 2 on *Integrative CCS Outcome* (Factor 1).

As the factor was identified in an unsupervised setting, without explicitly modeling the coronary artery status, this result indicates that integrative omics-based factors like *Integrative CCS* might potentially be clinically useful. The concept of MOFA analyses might provide a valuable tool to identify non-invasive diagnostic biomarkers for prospective validation in a large-scale independent clinical cohort of CCS patients.

Furthermore, we again analyzed which features dominate *Integrative CCS*. The factor was defined by many features of CD4^+^ and CD8^+^ T cells and captured mainly variance of those views (Fig. 6c). Expression of modulators of T cell antigen recognition, signal transduction and T cell activation such as CD3E, ICAM3 and TRAC^51, 52^ in activated cluster 0 CD4^+^ T cells had a high positive weight on *Integrative CCS*. In line, expression of PRDX2, known to be upregulated upon T cell receptor stimulation^53^, and CORO1A, JUNB as well as CD37, regulating T cell survival and homeostasis^54–56^, showed strong positive associations with *Integrative CCS* (Fig. 6d, Suppl. Fig. 18a,19a). In summary *Integrative CCS* was defined by a dysregulated activation pattern of the monocyte and T cell compartment (described in detail in the supplementary notes).

## Discussion

The systemic immune signatures associated with acute and chronic coronary syndromes remain incompletely understood, but are highly relevant for atherosclerosis, thrombosis, and myocardial remodeling. In particular, insights from Multi-Omics studies in patients might provide leads for developing new strategies for accessible biomarker signatures for diagnosis and prognosis in ACS and CCS.

Indeed, simultaneous Multi-Omics and single cell profiling in combination with unsupervised MOFA allowed us to obtain an unbiased description of the systemic immune signatures of coronary syndromes. *Integrative ACS* captured the time course of the immune response to ACS robustly across two independent cohorts. It was mainly defined by CD4^+^, CD8^+^ T cell and monocyte derived features, emphasizing the importance of these immune cells for the response to ischemic myocardial damage. EIF3E, pivotal for CD4^+^ T cell signaling^22^, as well as T cell HMGB1 were strongly associated with *Integrative ACS*. Multiple T cell derived ligands correlated with expression of downstream targets in monocytes. In line, several previous approaches focusing on whole blood or distinct immune cell subsets have provided essential

insights into systemic mechanisms that possibly drive atheroprogression or plaque rupture in coronary syndromes, unravelling a particular importance of CD4^+^ T cells phenotypes^44–47^. Moreover, in the OPTICO-ACS study, flow-cytometric analysis of local blood taken from the coronary arteries from patients with ACS by either plaque erosion or plaque rupture emphasized an important role for T cell-derived cytotoxic effector molecules in plaque-erosion ACS^46^.

Although all MOFA factors were estimated in an unsupervised manner, without using explicit information on outcome nor on the trajectories of specific patients, *Integrative ACS Outcome* predicted a favorable or poor treatment outcome of STEMI patients already at the time of hospital admission. This finding enabled the training of a supervised model that could then be applied in a second independent cohort, demonstrating the robustness and generalizability of the diagnostic performance of the identified integrative, multilayered biomarkers. Comparison to classical clinical laboratory markers (CK, CK-MB, Troponin T) showed that *Integrative ACS Outcome* outperformed established markers at early timepoints. *Integrative ACS Outcome* was defined by high-ranking features in NK cells such as *TXNIP*^34^*, GZMB*^35^ and *CD53*^39^. This could indicate NK cell cytotoxicity as a possible predictive marker of adverse outcome after myocardial infarction. Indeed, previous studies on lymphocyte-mediated cytotoxicity showed a deleterious role on post-ischemic cardiac remodeling^48^. Moreover, NK cells directly affect atherosclerosis and the regulation of pulmonary vascular permeability after myocardial infarction in experimental models^49, 50^.

*Integrative CCS*, again estimated in an unsupervised way, differentiated patients with CCS from patients without CAD, outperforming the established “SCORE2”^43^ CVD risk score. *Integrative CCS* should be considered a promising candidate integrative biomarker requiring independent replication in the future, as performed with *Integrative ACS* or *Integrative ACS Outcome*. High ranking features contributing to *Integrative CCS* indicated a dysfunctional T cell phenotype, possibly induced by a disrupted CD16^high^ monocyte-NAMPT-T cell signaling axis. NAMPT signaling has been associated with inflammatory monocyte/macrophage states and higher circulating NAMPT levels have been identified in ACS patients^51, 52^.

Our study holds multiple important implications for clinical, basic and translational cardiovascular research. First, we provide proof-of-concept that single cell Multi-Omic profiling of the circulating immune signature, paired with MOFA analyses allow prediction of disease state, phenotype, and outcome of remote, non-accessible injury sites. By reducing the very large number of potential predictors, in our case >13,000 variables of diverse datasets, including proteomics, single-cell transcriptomics and clinical data points to few factors, we identify overarching signatures of clinical significance. This implies that (1) integration and factor-based analysis of complex datasets can add crucial understanding of (cardiovascular) disease, and (2) Multi-Omic liquid biopsies without access to the tissue site of injury – in this case the infarcted cardiac muscle – with subsequent MOFA analysis offer a strategy for non-invasive diagnostics as well as disease course prediction also for non-accessible tissues.

In patients with chronic lymphatic leukemia, bulk RNA-seq with MOFA analysis identified a molecular signature predictive for treatment outcome that was mechanistically linked to the proliferation of tumor cells^53^. In this case, the blood-based data directly assessed the diseased tissue itself. Here, we demonstrate that this approach can be transferred to identify circulating biomarker signatures for risk stratification in solid organ disease by using routine blood drawings, and therefore elucidates novel tools for the concept of ‘liquid biopsies’ from circulating blood.

From a pathophysiological perspective, our data highlight that distinct, systemic immune states associate with coronary syndromes. In acute MI, these states follow distinct trajectories over the disease course from ischemia to reperfusion and healing/scar formation. Crucially, a specific signature also associates with favorable outcome – i.e. improvement of cardiac function. This underscores the possible importance of the identified axes to drive myocardial healing as well as targeting of pathological scar formation and supports the concept of therapeutic immune modulation to limit cardiac damage.

However, this study has limitations: The study is hypothesis generating but the power was not comparable to a clinical trial aiming to probe the predictive value of novel diagnostic markers for clinical use. An adequately powered, large-scale clinical trial is indispensable to evaluate the ultimate diagnostic performance of Multi-Omics factor analysis and to probe translation into standardized clinical settings. The methods used in this manuscript are highly cost-intensive, but selected candidate biomarkers such as expression of few genes in NK cells that are highly predictive of treatment outcome, could be assayed in a targeted way for such larger clinical cohorts. Finally, the hypotheses about the molecular underpinnings generated in these analyses require further mechanistic follow up studies.

In summary, this Multi-Omics study of coronary syndromes identifies systemic immune signatures representing multicellular gene programs that capture the time course of the immune response to ACS and enables early prediction of treatment outcome, which is of high clinical importance. Analysis of the components of the immune signatures reveal the underlying molecular and cell-cell communication pathways. These novel integrative analysis strategies set the stage for the identification of circulating multilayered immune-cell signatures that can be utilized as highly predictive stratification tools for diseased, but non-accessible tissues, using blood samples and subsequent MOFA analysis to outperform single-molecule biomarker approaches and clinically established laboratory tests.

## Data Availability

All data produced in the present study are available upon reasonable request to the authors

## Supplementary notes

### T cell function is the key driver of inter-patient variance in *Integrative CCS*

*Integrative CCS* was defined by many features of CD4^+^ and CD8^+^ T cells and captured mainly variance of those views (Fig. 6c). Expression of modulators of T cell antigen recognition, signal transduction and T cell activation such as CD3E, ICAM3 and TRAC^54, 55^ in activated cluster 0 CD4^+^ T cells had a high positive weight on *Integrative CCS*. In line, expression of PRDX2, known to be upregulated upon T cell receptor stimulation^56^, and CORO1A, JUNB as well as CD37, regulating T cell survival and homeostasis^57–59^, showed strong positive associations with *Integrative CCS* (Fig. 6d, Suppl. Fig. 18a<u>, 19a</u>). FOSB expression in activated cluster 0 CD4^+^ T cells, cluster 1 CD8^+^ T cells and cluster 3 NK cells had a negative weight on *Integrative CCS* (Fig. 6d, Suppl. Fig. 18a,19a, Suppl. Table 6). We further explored ligand-target correlations within *Integrative CCS* by again exploiting the ‘NicheNet’ database^27^ with ligands showing the highest weights on *Integrative CCS* (Suppl. Fig. 19b-e). Interestingly, ICAM3 expression in cluster 0 CD4^+^ T cells, cluster 1 CD8^+^ T cells and cluster 4 CD14^high^ monocytes was among the top 10 ligands with the highest weights on the factor (Suppl. Fig. 19b,c). Simultaneously, expression of the ligands CALM1 and CALM2 in similar clusters, involved in intracellular calcium homeostasis, showed high negative factor weights on *Integrative CCS* (Fig. Suppl. Fig. 19b,c)). PTMA (encoding Thymosin alpha-1) is highly expressed by T cells and has been shown to hold broad disease-protective effects^60–62^. ICAM3 or CALM1 from cluster 11 and 0 CD4^+^ T cells negatively correlated with PTMA expression in cluster 1 CD8^+^ T cells (Suppl. Fig. 19d,f,g). Moreover, CALM1 in multiple CD4^+^ T cell as well as B cell and monocyte clusters was accompanied with increased PTMA expression in cluster 1 CD8^+^ T cells (Suppl. Fig. 19d,f,g. In summary *Integrative CCS* can be characterized by a negative regulation of PTMA in CD8^+^ T cells by multiple cell types.

However, what sustains CD4^+^ T cell phenotype in *Integrative CCS*? NAMPT (encoding Nicotinamide phosphoribosyltransferase) in cluster 9 and 12 CD16^high^ monocytes showed a strong negative correlation with JUNB expression in CD4^+^ T cell clusters (Suppl. Fig. 19g). Expression of NAMPT in Cluster 9 and 12 also showed strong negative associations with *Integrative CCS* (Suppl. Fig. 19b, Suppl. Table 6). This ascribes unleashed CD4^+^ T cell JUNB levels in *Integrative CCS* to a reduced NAMPT expression in CD16^high^ monocytes (Suppl. Fig. 19e-g, Suppl. Fig.). NAMPT has been described to be a key regulator of monocyte differentiation^63^ particularly during inflammatory states^64^ and has been described to be increased in patients with ACS as well as in M1 inflammatory macrophages^52^. Ultimately, lower levels of NAMPT signaling trigger a dysfunctional T cell phenotype^65^. Multiplex cytokine analyses also revealed distinct differences between CCS and non-CCS patients, including GM-CSF, EGF, LIF, CCL4, IL-10 and IL-25 (Suppl. Fig. 20,21). In summary, integrative factor analysis suggests a dysregulated monocyte and T cell activation pattern in patients with CAD to define the disease compared to patients without CAD.

## Methods

### Munich Cohort: Ethics & patient cohort

Informed consent was obtained from the patients in accordance with the Declaration of Helsinki and with the approval of the Ethics Committee of the Ludwig Maximilian University of Munich (No.: 19-274). We collected blood from n=62 patients employing repetitive serial sampling and separately analysed the different immune cell constituents. For blood collection we used heparin-anticoagulated blood (i.e. Sarstedt AG & Co. KG, cat# 02.1065.001). A total of n=125 whole blood test, n=122 PBMC samples, n=246 plasma samples and n=121 PMNs samples were used for analyses. In the acute coronary syndrome (ACS) group, patients with ST segment elevation myocardial infarction (STEMI) were included and blood was analysed longitudinally. Blood sampling was done periinterventionally (TPM1) – during catheterization to avoid time loss, 14 (± 8) h after intervention (TPM2), 60 (± 12) h after acute event (TPM3) and before discharge, about 5-8 d after acute event (TPM4). A further subdivision was made into patients without direct reperfusion within 24h after symptom onset (delayed myocardial reperfusion, n=4) and patients with direct reperfusion within 24h due to coronary intervention (acute myocardial infarction, n=24). A subgroup of patients with evidence of infection in laboratory testing who were treated with antibiotics in the clinical setting defined a subgroup with hospital acquired infection (n=5) which was differentiated from the sterile group with STEMI without hospital acquired infections (n=19). The latter was used for comparison with the chronic coronary syndrome group (CCS). Patients were also subdivided based on clinical outcome. For this purpose, the ejection fraction (EF) measurement was determined according to Simpson’s method in echocardiography. A comparison was made between the findings on admission and during the hospital stay or before discharge (ΔEF). Based on these, a classification was made according to positive (good outcome) and negative (poor outcome) ΔEF in acute setting. The chronic cardiac event group included patients with an initial diagnosis of chronic coronary syndrome based on a cardiac catheterisation (lumen reduction of >50%) or coronary CT scan (>75 percentile) (CCS, TP0M n=16). Coronary healthy patients, with a catheter or CT based rule out of CAD, were included as a comparison group for the chronic coronary syndrome group (non-CCS, TP0M n=18). Coronary sclerosis was defined as coronary irregularities without significant lumen obstruction (<50%). The cardiovascular risk of CCS and non-CCS patients was calculated by using SCORE2^66^. The following parameters were used: Age, total cholesterol, HDL, systolic blood pressure, smoking history and gender.

Exclusion criteria for the Munich cohort were cardiogenic shock, age >85 and <30 years, severe systemic diseases (chronic liver disease, active haemato-oncologic diseases, active cancer, autoimmune diseases, acute inflammatory event with a CRP >2 mg/dl or fever at admission) and the use of immunosuppressants at inclusion. For the CCS cohort, patients with significant elevation of Troponin T levels were also excluded.

### Clinical blood tests

The clinical blood tests were performed as part of the treatment during hospitalisation. We involved the following clinical biomarkers and blood cells: CK, CK-MB, Troponin T, CRP, Leukocytes and Neutrophils. The statistical analysis and the graphical illustration were performed with Prism 9.

### Cytokine und Chemokine assays

For the isolation of plasma, 2x 1 ml whole blood was centrifuged at 2000 x g (rcf) for 20 min at 4°C (Centrifuge 5424 R, Eppendorf AG). Afterwards, the supernatant was carefully removed and pooled in a common Eppendorf reaction vessel for cryoasservation at -80°C.

For detection and quantitation of cytokines and chemokines, samples were sent on dry ice to EveTechnologies® to perform a Human Cytokine/Chemokine 71-Plex Discovery Assay® Array (HD71). Within the assay the following biomarkers were determined: 6CKine, BCA-1, CTACK, EGF, ENA-78, Eotaxin, Eotaxin-2, Eotaxin-3, FGF-2, Flt3L, Fractalkine, G-CSF, GM-CSF, GROα, I-309, IFNα2, IFNγ, IL-1α, IL-1β, IL-1RA, IL-2, IL-3, IL-4, IL-5, IL-6, IL-7, IL-8, IL- 9, IL-10, IL-12p40, IL-12p70, IL-13, IL-15, IL-16, IL-17A, IL-17E/IL-25, IL-17F, IL-18, IL-20, IL- 21, IL-22, IL-23, IL-27, IL-28, IL-33, IP-10, LIF, MCP-1, MCP-2, MCP-3, MCP-4, M-CSF, MDC, MIG, MIP-1α, MIP-1β, MIP-1δ, PDGF-AA, PDGF-AB/BB, RANTES, sCD40L, SCF, SDF- 1α+β, TARC, TGFα, TNFα, TNFβ, TPO, TRAIL, TSLP, VEGF-A.

The statistical analysis and the graphical illustration were performed with Prism 9.

### Plasma proteome analysis

The isolation and storage of the plasma was mentioned above. The plasma vials were slowly thawed at +4°C and mixed at a ratio of 1:5 with a proteomic buffer (2% SDS (Thermo Scientific, cat#J22638.AE), 2.5 mM DTT (Invitrogen, cat#P2325) in 50 mM Tris (Invitrogen, cat#AM9820)). Afterwards the samples were immediately boiled at 95°C for 10 minutes and cryoconserved at -80°C. Plasma samples were prepared by SDS-lysis, automated SP3 cleanup and tryptic digest essentially as described^67, 68^. Samples were measured on an orbitrap Exploris 480 instrument (Thermo Fisher Scientific) in label-free data-independent acquisition (DIA) mode whilst separating peptides on a 44 min gradient on a nanoEASY 1200 system (Thermo Fisher Scientific) coupled to the mass spectrometer. Raw files were analysed in Spectronaut 14 (Biognosys^69^) against a spectral library that was generated from 52 fractions measured in the same manner as described^67^. An FDR cutoff of 0.01 was applied and spectra were searched against a human Uniprot database from 2018 including isoforms. For data filtering, the option Qvalue percentile with a fraction of 0.2 was used and global normalisation by median was applied. Further downstream analysis was performed in R. Normalised intensities were filtered for at least 80% valid values per row and column, remaining missing values were median-centred and imputed using a randomised Gaussian distribution with a downshift of 1.8. For significance calling, the limma package was consulted to calculate moderated t statistics^70^. Nominal p-values were corrected using the Benjamini-Hochberg method.

### Isolation of polymorphonuclear neutrophils (PMNs)

Initially, 400µl of whole blood was added to a tube and 20μl each of the Isolation Cocktail and RapidSpheres (EasySep Direct Human Neutrophil Isolation kit, STEMCELL Technologies Inc., cat#19666) were added. After 5 min incubation at RT, the reagent was filled up to 4ml with PBS (Dulbecco’s Phophate Buffered Saline (1x), ThermoFisher Scientific, cat#14190-094) + 1mM EDTA (Ethylenediaminetetraacetic acid, Sigma-Aldrich Chemie GmbH, cat#03690). Subsequently, the tube was placed in a magnet (EasySepMagnet, STEMCELL Technologies Inc.) for 5 min. Then, without being removed from the magnet, the contents of the tube were transferred to a new tube in a continuous motion. After adding 20μl RapidSpheres again, the incubation steps were repeated without adding PBS + 1mM EDTA. After decanting again, the new tube was placed in the magnet for 5 min without addition of RapidSpheres. The newly decanted tube was centrifuged at 320 x g for 7 min at 4°C (Centrifuge 5810 R, Eppendorf AG). The pellet was then resuspended in 100μl PBS. 10µl of the suspension was used to adjust the concentration to 5 million cells/ml. For this, the dead cells were stained with trypan blue (Sigma-Aldrich Chemie GmbH, cat#T8.154) and the concentration was calculated using a Neubauer counting chamber (LO-Laboroptik Ltd, Lancing). For cryoasservation at -80°C, cell suspension was added to RLT plus buffer (Qiagen GmBH, cat#1053393) containing 1% 2-mercapathoethanol (Sigma-Aldrich Chemie GmbH, cat#M3148) at a ratio of 1:10.

### prime-seq

For the analysis of the transcriptome of PMNs, prime-seq, an early barcoding bulk RNA-seq method, was used. Samples were pre-treated with proteinase K (Life Technologies, cat#AM2548) followed by isolation with cleanup beads (Sigma-Aldrich, cat#GE65152105050250) (ratio 2:1 beads per sample). DNase I (Thermo Fisher, cat#EN0521) was used to digest the cells to make the transcriptome accessible for the process of reserve transcription. This was done by adding 30 units of Maxima H enzyme (Thermo Fisher, cat#EP0753) and 1x Maxima H buffer (Thermo Fisher, cat#EP0753), 1 mM dNTPs (Thermo Fisher, cat# R0186), 1 µM template-switching oligo (IDT) and 1 µM barcoded oligo-dT primers (IDT) and incubating for 90 minutes at 42°C (reaction volume: 10µl). After pooling of all samples, they were purified in a 1:1 ratio with cleanup beads. For elimination of the leftover primers, exonuclease I (NEB, cat#M0293L) was added (Incubation setup: 37°C for 20 min, then 80°C for 10 min), followed by another purification with cleanup bead s. The synthesis of the second strand of cDNA was prepared by adding 1X KAPA HS Ready Mix (Roche, cat#07958935001) and 0.6 µM SINGV6 primer (IDT) (reaction volume: 50µL). For amplification, subsequent PCR cycles were performed: Start: 98 °C for 3 min; 15 cycles: 98 °C for 15 s, 65 °C for 30 s, 72 °C for 4 min; End: 72 °C for 10 min. To re-purify the sample, cleanup beads were added at a ratio of 0.8:1 beads per sample and dissolved out in 10 µL DNase/RNase-free distilled water (ThermoFisher, cat#10977-049). Quantification and size selection of the purified cDNA was then performed using the Quant-iT PicoGreen dsDNA Assay Kit (ThermoFisher, cat#P7581) and the High-Sensitivity DNA Kit (Agilent, cat#5067- 4627). For library preparation a fivefold lower reaction volume of the NEBNext Ultra II FS Library Preparation Kit (NEB, cat#E6177S) than recommended by the manufacturer was used. Fragmentation of cDNA was performed using the enzyme mix and the reaction buffer (reaction volumes 6µl) and ligation was performed using Ligation Enhancer, Ligation Master Mix and a custom prime-seq adapter (1.5 µM, IDT) (reaction volume: 12.7 µL). SPRI-select beads (Beckman Coulter, cat#B23317) were then used for a double size selection (ratio of 0.5 and 0.7). For amplification, Q5 Master Mix (M0544L, NEB), 1 µL i7 Index Primer (Sigma-Aldrich) and 1µL i5 Index Primer (IDT) were used followed by PCR (start: 98 °C for 30 s; 13 cycles: 98 °C for 10 s, 65 °C for 75 s, 65 °C for 5 min; end: 65 °C for 4 min). After successful size selection with SPRI-select beads and a quality check, the libraries were sequenced by using a NextSeq (Ilumina).

The sequencing reads were processed using zUMIs pipeline using the Gencode human release version (https://www.gencodegenes.org/human/release_35.html). Only barcodes matching the expected samples were considered and exported as count matrices, both raw counts and library-size normalized ones. First, the data was checked using fastqc (version 0.11.8^71^). Regions on the 3’ end of the fragment reading into the poly-A tail were removed by Cutadapt (version 1.12^72^). The zUMIs pipeline (version 2.9.4d)^73^ was applied, filtering the data, with a phred threshold of 20 for 4 bases the UMI and BC, mapping the reads to the human genome with the Gencode annotation (v35) using STAR (version 2.7.3a), reads were counted using RSubread (version 1.32.4)^74, 75^.

### PBMC isolation

For isolation of peripheral blood mononuclear cells (PBMCs), 8 ml of whole blood was transferred to a BD Vacutainer® CPTTM (Becton, Dickinson and Company, cat#362780), swivelled twice and centrifuged at 1650 x g for 20 min at room temperature (RT) (Centrifuge 5810 R, Eppendorf AG). After swivelling twice, the supernatant was transferred into a 15 ml tube and a further centrifugation step with 350 x g for 7 min at 4°C was performed. The resulting cell pellet was resuspended in 4 ml freezing medium and alliqouted. The freezing medium consisted of 45% RPMI (VLE-RPMI 1640, Bio&SELL GmbH, cat#BS.52551528.5) with 1% glutamine (GibcoTM L-Glutamine (200 mM), Thermo Fisher Scientific, cat# BS.K0283), 45% FBS (FBS SUPERIOR stabil®, Bio&SELL GmbH, cat#FBS.S0615) and 10% DMSO (Dimethyl sulphoxide, Sigma-Aldrich Chemie GmbH, cat#D2438). For cryoasservation, samples were slowly frozen in a Mr. Frosty freezing container (Thermo Fisher Scientific, cat#5100-0036) at -80°C for 24h and then transferred to -80°C freezers.

### FACS and scRNA-seq preparation

For scRNA-seq analysis of the frozen PBMCs, an adapted thawing protocol of 10X was used (*Fresh Frozen Human Peripheral Blood Mononuclear Cells for Single Cell RNA Sequencing*, Document Number CG00039 Rev E, 10x Genomics, (2023, May 2^nd^): https://assets.ctfassets.net/an68im79xiti/71r5PbRPB1LeqRkuPltBzr/64cfaa099d0a7fd41f79a4aecd643926/CG00039_Demonstrated_Protocol_FreshFrozenHumanPBMCs_RevE.pdf.

Samples were thawed at 37°C for 3 min. This was followed by stepwise dilution (5x 1:1) with dropwise addition of complete growth medium. The complete growth medium consistit (10% FBS + 90% RPMI). The sample was then filtered using a 50 µm strainer and centrifuged at 300g for 5 min at RT. Supernatant was removed to the last millilitre and the cell pellet was resuspended in it by using a wide bore pipet. After slowly adding another 9 ml of complete growth medium, the sample was split into two. A further centrifugation step at 300 rcf for 5 min at 4°C was performed. One half of the sample was used for further processing for scSeq analysis. The cell pellet was resuspended in 100µl Fc block (BD Pharmingen, cat#564200) (1:50) and incubated on ice for 10 min. To label the cells, TotalSeqB™ anti-human hashtag antibodies (1:500) (BioLegend) were added to the sample and then incubated at 4°C for 30 min. To maximize the performance, TotalSeqB™ anti-human hashtag antibodies were pre-centrifuged at 14000g at 4°C for 10 min. Following this, the sample was washed 3 times by adding 5ml of FACS buffer (PBS with 0.5% BSA (Albumin Fraktion V, Carl Roth GmbH & Co. KG, cat#8076.4)) and centrifugation at 250g for 10 min at 4°C each time. After the last centrifugation step, the cell pellet was resuspended in 0.04% BSA in PBS and the concentration was adjusted to 200 cells/µl using a Neubauer counting chamber. Lastly, marked samples were pooled. The other half of the sample was used to prepare the FACS analysis. The sample was incubated with 200µl Fc block (1:50) at 4°C for 10 min. Staining of the cells was done by a 20 min incubation with an antibody mastermix (1:400). After centrifugation at 300g for 7 min at 4°C, the cell pellet was resupended in 300µl FACS buffer. The dead cells were stained immediately before flow cytometry with LSRFortessa Flow Cytometer (BD Biosciences). The flow cytometry data were analysed with FlowJo (BD). The statistical analysis and the graphical illustration were performed with Prism 9.

### Antibody panel

| Color | Antigen | Company | Cat. # |
| --- | --- | --- | --- |
| BV650 | CD45 | BioLegend | 304044 |
| FITC | CD14 | BioLegend | 557153 |
| PE | CD16 | BioLegend | 302008 |
| PERCP-Cy5.5 | CD8 | BioLegend | 344710 |
| APC-Cy7 | CD19 | BioLegend | 363010 |
| BV 605 | HLA-DR | BioLegend | 307640 |
| BV786 | CD123 | BioLegend | 306032 |
| BV510 | CD3 | BioLegend | 317332 |
| BV 711 | CD56 | BioLegend | 362542 |
| AF 700 | CD34 | BioLegend | 343622 |
| BV421 | CD4 | BioLegend | 357424 |
| SytoxRed | Dead cell stain | Invitrogen | S34859 |

### ScRNA Library preparation

For single cell sequencing, libraries were prepared using the Chromium Next GEM Single Cell 3’ Reagent Kit v3.1 (CG000206 Rev D) from the 10X Genomics® protocol. Barcode-based multiplexing with TotalSeqB™ anti-human hashtag antibodies was performed to reduce artefacts associated with batch variation. According to the manufacturer’s instructions, the Gel Beads-in-emulsion (GEMs) were first prepared to obtain cDNA with reserve transcription. After purification of the cDNA, an amplification and size selection were performed. After final quantification and quality control, the gene expression and cell surface libraries were constructed for sequencing, which was performed by using an Illumina NovaSeq.

### Bioinformatics analysis of the scRNA-seq dataset

#### Pre-processing

##### SC Data Preparation (cellranger)

After sequencing, the FASTQ files for the gene and cell surface libraries were processed using the cellranger count pipeline (chemistry: Single Cell 3’ v3; pipeline version: 3.1.0). Each sample was mapped to the human reference genome (GRCh38; version: 3.1.0). The library and reference files were created according to the 10X Genomics instructions and example files for Antibody Capture with TotalSeq™-B (https://support.10xgenomics.com/single-cell-gene-expression/software/pipelines/latest/using/feature-bc-analysis#feature-ref (17^th^ Jan 2023)).

The pipeline quantifies each feature (genes + antibodies) in each cell and generated quality control summaries and feature barcode matrices for each of the 14 libraries (Suppl. Table 3). For further analysis we took the ‘filtered_feature_bc_matrix.h5’ of each library and split it up into two separate anndata objects: one containing the gene expression and one the antibody capture counts.

##### Demultiplexing and Doublet Identification

For the demultiplexing of the data we take the antibody capture counts anndata objects of each library convert them to Seurat objects, normalize the counts with ‘CLR’ (centered log-ration transformation) and apply the ‘HTODemux’ function of the Seurat package (version: 4.1.1) as described in the vignette with the default 0.99 quantile threshold in order to classify the cells as positive or negative for each HTO (hashtag oligo). Cells that have been classified as positive for more than one HTO have been annotated as doublets.

##### Cell quality control and filtering (QC)

In a next step, we transferred the cell annotation results from the demultiplexing to the gene expression anndata files and applied some cell quality control criteria and filtering based on the gene expression counts to remove low quality cells. These steps were done for each library separately.

In a first basic filtering step we kept only cells that have counts on at least 200 genes and genes that have counts in at least 3 cells. Furthermore, we combined the percentage of mitochondrial gene counts (pct_counts_mt), number of genes by counts (n_genes_by_counts) and total counts (total_counts) criteria to filter out further cells. We only kept cells that have:

- n_genes_by_counts < 5000 ∩ total_counts < 20000
- n_genes_by_counts > 500 ∩ pct_counts_mt < 15

Subsequently the data was normalized (10.000 counts per cell) and log transformed (log1p) using the scanpy toolkit^76^ 1.8.1 in Python v.3.9.6. Furthermore, we excluded mitochondrial and ribosomal genes as they were not of interest for the analysis.

##### Data Integration, Clustering and Cell-Type Annotation

In order to get a joint embedding of the complete dataset and correct for potential batch effects between the libraries we took the processed data from the QC and applied the Scanorama method (scanorama.correct_scanpy) as explained in the package description using 2000 highly variable genes, batch-size parameter of 2000 and default parameters. This returned a Scanorama corrected count-matrix and a joint-embedding which we then use as input for the computation of a neighbourhood graph (scanpy.pp.neighbors, n_neighbors = 10, n_pcs = 50) and the subsequent clustering of the cells using the Leiden algorithm (scanpy.tl.leiden; default parameters).

We found 18 different clusters which we annotated manually by looking at the expression patterns of PBMC marker genes selected based on literature research and calculating differentially expressed genes (DEGs) between the clusters using a Wilcoxon rank sum test with Bonferroni-Hochberg adjustment as implemented in the scanpy framework (scanpy.tl.rank_genes_groups). With this strategy we could annotate all 18 clusters to all common major peripheral blood mononuclear immune cell-types (Suppl. Fig. 2a,b).

#### Data Analysis

##### Compositional Analysis

In order to investigate compositional changes of the cell-type clusters between the different patient groups and timepoints we determined the percentage of cells that have been assigned to the different cell-type clusters for each patient and timepoint separately (for each patient and timepoint: amount of cells belonging to the cluster/ total amount of cells) and adjust them with centered log-ratio (CLR) transformation. Adjusted values were then analyzed using the Ordinary One-Way ANOVA with correction for multiple comparisons by Dunnett’s test (*p≤0.05, **p≤0.01). The statistical analysis and the graphical illustration were performed with Prism 9.

### Multi-Omics Data Integration (MOFA)

#### Data Harmonization and Model Training

##### Data Harmonization and Integration

For the integrated and combined analysis of all the different data sources (single-cell data, cytokines, neutrophils, proteomics and clinical data) we applied several pre-processing and normalization steps separately on the features of the different types of data to make them comparable and adjust the distributions.

###### ScRNA-seq Data

We applied the pseudo-bulk approach to summarize single-cell data on the level of celltype (cluster) specific gene expression per sample because all other omics data was measured on the bulk-level. To this end, we calculated for each of the identified 18 cell-type clusters for each sample (= patient and timepoint) the mean counts across all cells. Afterwards we adjusted the gene counts of each sample in each cluster with a scaling factor so that each sample has the same amount of counts across all genes to account for technical differences in sequencing depth between the samples.

In order to ensure that we only consider reliably expressed genes we applied additional filtering steps on the genes and clusters:

We excluded clusters 14-18, which had only very low numbers of cells per patient and timepoint (mostly less than 10 cells).

We filtered out genes based on the total number and percentage of cells that expressed those genes in the corresponding cluster keeping only genes that fulfill one of the criteria below:

- percentage of cells expressing gene > 50 ∩ total number of cells expressing gene > 1200
- percentage of cells expressing gene > 40 ∩ total number of cells expressing gene > 3000

The thresholds were chosen considering that gene should be detectable in a high number of cells and in several samples but at the same time a considerable number of genes for each cluster should be kept.

After filtering we log transform the count values and apply quantile normalization as further normalization steps to align the distributions of gene expression levels between the samples. This results in 11.831 features (which correspond to genes) across all the different clusters (ranging from 315 for cluster 13 and 2159 for cluster 4) which we used as input for the different cell-type cluster dimensions from the single-cell data for the MOFA analysis (Suppl. Fig. 3a).

###### Cytokines

In order to prepare the cytokine data for integration with the other datasets we set ‘OOR’ values to 0 and log transform the values to adjust the distributions after adding a pseudocount of 1 to all values. Furthermore, we exclude cytokines which have valid measured values in less than 20% of the samples. In total this results in 65 different cytokines which have been used as input features for the integrated analysis (Suppl. Fig. 3a).

###### Neutrophils

As input features from the neutrophil dimension, we take the umi exon counts from the prime-seq and apply the processing steps below to align the reads with the single-cell sequencing data, adjust for potential technical effects and strictly remove samples and genes with low quality reads.

In a first step we adjusted the gene names and map them from ‘ENSEMBL’ gene-ids to ‘SYMBOL’ gene-ids. Then we filtered out ribosomal and mitochondrial genes as we also excluded them in the single-cell pre-processing and are not relevant for the analysis. Furthermore, we excluded genes that are not expressed in at least 80% of the samples and removed samples that do not have reads in at least 90% of the remaining genes. In a next step we adjusted for differences in sequencing depth between the samples and normalized the counts with a scaling factor so that the sum of reads for each sample are the same. Then we logarithmized the resulting counts. Finally, we decided to keep only highly variable genes, so we removed all genes which variance lies below the 25% quantile of the variance distribution. This resulted in a total of 892 genes measured on 92 samples which are considered as input features for the neutrophil dimension. As for the single-cell data we applied quantile normalization to the counts in a final normalization step (Suppl. Fig. 3a).

###### Proteomics

For proteomics, we used the same pre-processing and normalization steps as in the described in the previous Plasma Proteome analysis and took the resulting normalized and median-centered intensities measured for 490 different proteins as input features of this dimension (Suppl. Fig. 3a).

###### Clinical Data

As input features of the clinical data dimension, we used the measured CK, CRP, CK-MB and Troponin values and log transformed then (Suppl. Fig. 3a).

##### Model Training

After these individual pre-processing steps, we had in total 13.382 features across the 18 different dimensions (cell-type cluster 1-14, neutrophils, cytokines, proteomics and clinical data) and applied feature-wise quantile normalization onto the quantiles of the standard normal distribution for all data types. Then we train the MOFA model using the R/Bioconductor package MOFA2 (version: 1.2.2) with maxiter parameter 50.000 to ensure convergence and 20 factors and the remaining default parameters. The number of estimated factors was chosen to balance the trade-off between explained variance and low number of factors after various tests with different parameters (Suppl. Fig. 3a).

To evaluate the effect of the clinical features we train a second MOFA model excluding the 4 clinical features with in total 13.378 features and compare the resulting factor values and feature weights to the original model.

#### Downstream Analyses

##### Gene sent enrichment analysis (Pathways)

On the feature weight matrix resulting from our trained MOFA model we conduct pathway enrichment analysis for the first 5 inferred factors of the MOFA model using the gene set annotations from the REACTOME^25^ and KEGG^77^ databases. We test all pathways belonging to the ‘Immune System’ category in REACTOME (n=191) and pathways that are classified as ‘Immune system’ or ‘Signal transduction’ pathways in KEGG (n=52).

To test the enrichment of the pathways across all our data input dimensions we generate an extended pathway gene annotation set for those pathways in which a feature (consisting of data dimension and gene/protein code) is considered to belong to the pathway if the gene/protein code maps to the genes annotated to the pathway. To map the gene/protein codes to the gene-set annotations in KEGG and REACTOME we use the bitr function from the clusterProfiler package (version 4.0.5) to convert them to ENTREZID.

We remove all the pathways for which we have included less than 20% of the total amount of genes annotated to the pathway in our feature set and run the enrichment analysis using the ‘run_enrichment’ method implemented in the R/Bioconductor package MOFA2 (version: 1.2.2) with set.statistic parameter ‘rank.sum’ and default parameters otherwise. We run the enrichment separately for features with only positive or negative weights and jointly across all features.

Pathways with an adjusted p-value < 0.05 (Benjamini-Hochberg adjustment) have been considered to be significantly enriched.

##### Cell-cell communication

To analyze the potential axes of cell-cell communication between different cell-types we use the prior knowledge about potential ligand-receptor-target interactions of the NicheNet^27^ resource collected in the nichenetr package (version: 1.1.0) and load the provided ligand-receptor network and ligand-target matrix^78^. Based on the classifications in those networks we identify ligands, receptors and potential targets among the 13.382 features included in our integrated dataset resulting from the ‘Data Harmonization and Integration step’. We calculated spearman correlation between all identified ligand-target pairs within this dataset.

For the further analysis of the calculated ligand-target correlations in combination with the corresponding regulatory potential score we only consider ligand-target pairs:

- between ligand and targets of different cell-types (e.g., between monocytes and T cells) and different views (e.g., between cytokines and the different cell-type clusters).
- where we have reliably measured a receptor in the target cell-type-cluster to which the ligand might potentially bind to as specified by ligand-receptor network provided by the NicheNet^27^ resource in order to affect the target gene (in case the target belongs to one of the cell-type cluster views from the scRNA-seq data). We consider a receptor gene to be reliably measured in case it fulfills one of the thresholds below:

`o` percentage of cells expressing receptor gene in cell-type cluster > 30 ∩ total amount of cells expressing receptor gene in cell-type cluster > 600
`o` percentage of cells expressing receptor gene in cell-type cluster > 10 ∩ total amount of cells expressing receptor gene in cell-type cluster > 1200

Subsequently we focus on pairs with high correlation and regulatory potential scores where the target gene has a high feature weight on the analyzed MOFA factor.

##### Predictions

To evaluate the prediction potential of our MOFA factors to distinguish our different patient groups we calculate ROC curves contrasting the prediction power of the inferred factors to established clinical markers.

For Factor 4 predictions we only consider factor values from samples measured at TP1 that could be classified to have a ‘good’ or ‘poor’ outcome. We compare the prediction potential of the factor values to the value of the clinical markers (CK, Troponin, CRP) for those samples at TP1. For the benchmarking against the prediction power across the complete time-course of the clinical values we take the maximum and mean values of those markers across all measured timepoints. In both cases we scale the clinical values as well as the factor values to be in a range between 0 and 1(￼) and use them as input for the ROC curve calculation giving the probability of a sample being classified as ‘good’ vs. ‘poor’ outcome.

For Factor 1 predictions we only consider factor values from samples classified as ‘CCS’ or ‘non-CCS’ and contrast those to the prediction power of calculated ‘SCORE2’^43^. for those samples. As for Factor 4 we scale these values to be in a range between 0 and 1 and use them as input for the ROC curve calculation giving the probability of a sample being classified as ‘CCS’ vs. ‘non-CCS’ sample.

### Replication in validation cohort (Groningen dataset)

To validate our approach and findings we used an independent second dataset including n=24 patients measured across three different timepoints (TP1G: at the heart catheterisation, TP2G: 24 hours after admission, TP3G: after 6-8 weeks) and a control group (TP0G, n=31) contained within the Groningen study^17. 79,80,81^t1: at the heart catheterisation, TP2Gt2: 24 hours after admission, TP3Gt3: after 6-8 weeks) and a control group (TP0G, n=31) generated within the Groningen study^17^. The Groningen study was part of the CardioLines biobank ^79^. From all patients informed consent was obtained. As a control group age and sex-matched participants from the the LifeLines^80, 81^.: at the heart catheterisation, TP2Gt2: 24 hours after admission, t3: after 6-8 weeks) and a control group (n=31) generated within the Groningen study^17^. The Groningen study was part of the CardioLines biobank^17^. From all patients informed consent was obtained. As a control group age and sex-matched participants from the LifeLines^17^.

Further specifications of the dataset and the processing can be found within the corresponding manuscript^17^. The data in the Groningen study^17^ was measured with two different chemistries: v2 10x chemistry and v3 10x chemistry which showed strong technical differences in gene expression profiles between the samples that were prepared with different chemistries. Therefore, a separate processing of both datasets was necessary. In our replication we focused on samples measured with the v2 10X chemistry as this cohort included a higher number of samples (v2: n=55; v3: n=21). The V3 10X chemistry cohort did not include a sufficient number of samples which could be divided into poor or good outcome groups and would have therefore been underpowered. Classification into good and poor outcome groups in the Groningen cohort was performed similarly as in the Munich cohort. Based on the ΔEF from echocardiography results (during the hospital stay and follow up), a classification was made according to positive (good outcome, n=7) and negative or stable (poor outcome, n=5) values.

For the replication, in a first step, we evaluated the alignment of the different strategies for cell-type annotations that were applied in the two different studies. Subsequently we harmonized annotations between both datasets and developed strategies to replicate findings on the MOFA factors that were presented within the current study. MOFA Factor 1 (*Integrative CCS*) could not be evaluated in the Groningen cohort as this dataset did not include the differentiation of the control group into ‘CCS’ and ‘non-CCS’ patients.

#### Alignment of cell-type annotations

In the Groningen study cell-type annotation was done using the automated Azimuth method (for more details see manuscript^17^). As our data was processed and annotated in a different way in a first step, we compared clusters and annotations resulting from our study to the Groningen study. For this we run the pre-processing and automated annotation strategy as described in the Groningen study^17^ on our data and compared the resulting annotations of the single cells to the annotations resulting from our initial clustering and manual annotation strategy (Suppl. Fig. 4).In the Groningen study cell-type annotation was done using the automated Azimuth method (for more details see manuscript^17, 82^). As our data was processed and annotated in a different way in a first step, we compared clusters and annotations resulting from our study to the Groningen study. For this we run the pre-processing and automated annotation strategy as described in the Groningen study^17^ [insert reference] on our data and compared the resulting annotations of the single cells to the annotations resulting from our initial clustering and manual annotation strategy (Suppl. Fig. 4). The results from this comparison are outlined in Supplementary Fig. 4. In general, our clustering and the automated azimuth annotation resulted for some cell-types in more granular (e.g. B cell cluster 10 would be distributed across ‘B naive’, ‘B memory’ and ‘B intermediate’ azimuth annotations) or more aggregated annotations (e.g. CD14^high^ monocyte clusters 4, 6 and 7 would all be aggregated as CD14^high^ monocytes) but on a more aggregated level annotations aligned well except for some T cell clusters (namely cluster 1 CD8^+^ T cells, cluster 11 CD4^+^ T cells and cluster 5 CD4^+^ T cells).

#### Replication of MOFA analysis with harmonized cell-type annotations

To evaluate the effect of the different granularity levels of annotation on the MOFA factor results in a subsequent step we run the same MOFA analysis as outlined above on the Munich data this time using instead of our manually annotated cell-type clusters the annotations resulting from the pre-processing and automated azimuth annotation as described in the Groningen manuscript (Suppl. Fig. 5a). We evaluated whether the resulting factors were able to capture the same patterns as found with our original strategy and whether factor and feature weights of the newly inferred factors and the factors presented in the manuscript aligned well by correlating them (Suppl. Fig. 5b, 6a,b). Overall, we could also reproduce the patterns presented previously with the alternative annotations (Suppl. Fig. 5b) and the inferred factor and feature weights of the presented factors were highly correlated (|cor| > 0.8) (Suppl. Fig. 6a,b).

In order to map the different input features based on the different cell-type annotation levels we mapped features as given in the table below:

| **Munich approach: clusters and annotations** | **Groningen approach: automated azimuth** |
| --- | --- |
| CD14<sup>high</sup> monocytes clusters (cluster 4, 6, 7) | CD14 Mono |
| B cells (cluster 8, 10) | B cell (aggregating 'B naive', 'B memory', 'B intermediate') |
| CD16<sup>high</sup> monocytes (cluster 9, 12) | CD16 Mono |
| CD4<sup>+</sup> T cells (cluster 0, 2) | CD4TCM |
| Dendritic cells (cluster 13) | cdC2 |
| NK cells (cluster 3) | NK |
| CD4<sup>+</sup> T cells (cluster 5, 11) | / |
| CD8<sup>+</sup> T cells (cluster 1) | / |

As an example: the weight of a specific gene from cluster 4 CD14^high^ monocytes (e.g. HMGB1) would then be matched to the same gene (e.g. HMGB1) of the aggregated CD14 Mono dimension given by the automated azimuth annotation.

Features from cluster 1, 5 and cluster 11 were not mapped due to the low alignment between the clusters and corresponding unique azimuth annotations.

#### Processing of Groningen scRNA seq data

In the next step we applied the pre-processing steps as described within the *ScRNA-seq Data* paragraph for the MOFA analysis also on the Groningen scRNA-seq dataset resulting in normalized pseudobulk-aggregated features per annotated azimuth cell-type in the Groningen data.

As the expression values of the genes where notably lower in the Groningen dataset we made some minor adjustments to the requirements with regards to the percentage of cells expressing a gene and the total amount of cells expressing a gene to get a comparable set of features as in our data:

- percentage of cells expressing gene > 30 ∩ total amount of cells expressing gene > 1000
- percentage of cells expressing gene > 20 ∩ total amount of cells expressing gene > 2500

After applying these pre-processing steps on the Groningen data, we had in total 6.353 features across the 13 different dimensions (azimuth cell-types 1-13).

#### Factor 2 (time pattern replication)

To replicate the Factor 2 on the Groningen dataset in a first step we mapped input features from the Munich dataset to the Groningen dataset as outlined in the table above and kept only features available in both datasets after the pre-processing (therefore cytokine, clinical, proteomics and neutrophil features as well as some genes from the scRNA-seq dataset not within both datasets were removed). We used the resulting feature-weight matrix from the Munich azimuth MOFA estimation (W^MU^) and calculated the right inverse to then apply it on the normalized input data (Y^GR^) from the Groningen cohort to infer the corresponding sample factor matrix (Z^GR^) for the Groningen cohort as shown below. The pattern across timepoints of the resulting sample factor matrix (Z^GR^) was then compared to the pattern given within the Munich dataset.

*Z*^*GR*^ = *Y*^*GR*^ ⋅ (*W*^*MU*^)^−1^ with: (*W*^*MU*^) ^−1^ = *W*^*T*^(*WW*^*T*^)^−1^

(GR = Groningen cohort, MU = Munich cohort)

#### Factor 4 (prediction replication)

For Factor 4 the main goal of our replication was to evaluate the potential of top-ranking features on Factor 4 to predict the outcome already at an early stage (TP1). As Factor 4 is derived based on the pattern across all four timepoints measured in our data and top-ranking Factor 4 features are not only characterized by the variation at TP1 between ‘good’ and ‘poor’ outcome samples but also by the variation across the different timepoints we chose to add another step to our analysis to identify those features within the top-ranking features that have high prediction potential only looking at their TP1 values. For this we chose to select the intersection of the top 280 features of the *Integrative ACS Outcome* Factor (corresponding to roughly 20% of the total amount of features) based on the MOFA model estimated with our manual annotation strategy and the MOFA model with the automated azimuth annotation. Subsequently, we trained a lasso model (logistic regression) with cross validation using the cv.glmnet function of the R package glmnet (version: 4.1.7; alpha=1; family = ‘binomial’; other: default parameters) taking as input only the value of those features at TP1M and predicting the outcome of the samples.

Then we applied this trained model on the same set of features with their values at TP1G on the Groningen dataset considering this dataset our holdout test dataset and evaluated prediction performance for those samples calculating AUC values.

Additionally, to evaluating the broad selected top-ranking feature set of Factor 4 in this way we also evaluated the prediction potential of features that we highlighted in the paper (namely NK cell features CD74, TXNIP, GZMB) based on potential biological mechanisms and trained a logistic regression model for these features on the Munich data which we then applied to the Groningen cohort.

### Echocardiographic assessment of LV function

Echocardiographic assessment of left ventricular ejection fraction (LVEF) as a proxy for systolic function was performed according to current guidelines. In brief, B-mode echocardiography was performed by cardiac intensive care unit (C-ICU) fellows and, following discharge from the ICU, by cardiology residents. LVEF was then measured using the biplane summation-of-disks method in apical four chamber (A4CH) and apical two chamber (A2CH) views by an experienced cardiology fellow that was unaware of patient outcomes at the time of measurement.

## Figure alignment, data and code availability

Figures were aligned by Adobe Illustrator.

All mapped data will be made available from the German Human Genome Archive (GHGA) and the European Genome-Phenome Archive (accession number pending).

All code will be made available on github at https://github.com/heiniglab/stemi_mofa

## Acknowledgments

We thank the patients and the families for participating in the study.

## Authorship contributions

Initiation and experimental design: K.P., L.N. K.S; Project administration and supervision: K.P., M.H., L.N. K.S.; Conceptualization: K.P., C.L., V.K., M.H., L.N. K.S.; Writing original draft: K.P.; Patient identification, patient information and informed consent acquisition, sample acquisition: K.P., T.P., L.N. K.S.; Methodology, investigation and formal analysis: K.P., V.K., C.L., L.N., M.J., S.B., V.K., L.E., R.K., A.J., V.P., R.E., O.P., M.H.; Writing & editing, all authors; Visualization: C.L., V.K.; Funding acquisition: K.P., N.H., P.M., W.E., R.Z., S.M., M.H., L.N., K.S.

## Tables

TABLE 1: Clinical characteristics of Munich patient cohort

TABLE 2: Clinical characteristics of Groningen patient cohort (V2) TABLE 3: Cellranger QC statistics

TABLE 4: Sample Meta Data and Cell Amounts Overview TABLE 5: Variance Decomposition

TABLE 6: Feature Factor Weights

TABLE 7: Pathway Enrichment Results REACTOME TABLE 8: Pathway Enrichment Results KEGG TABLE 9: AUC Values TP1 good vs. poor

TABLE 10: AUC Values CCS vs. non-CCS

**Suppl. Fig. 1**

**(a)** Clinical blood tests. Individual timepoints of sterile ACS (TPM1-4) compared to CCS (TP0M). CRP (ACS: TP1M n=17, TP2M n=18, TP3M n=17, TP4M n=12; CCS: TP0M n=15); Neutrophils (ACS: TP1M n=6, TP2M n=5, TP3M n=5, TP4M n=3; CCS: TP0M n=5). Parametric distributed data were analyzed using the Ordinary One-Way ANOVA with correction for multiple comparisons by Dunnett’s test; non-parametric distributed data were analyzed using the Kruskal-Wallis test with correction for multiple comparisons by Dunn’s test. *p≤0.05, **p≤0.01, ***p≤0.001. Illustration of the mean value with SEM. **(b)** Gating strategy used for flow-cytometric quantification of PBMCs in blood. **(c)** Analysis of the relative proportion of phenotypically defined immune cells to CD45^+^ leukocytes in PBMCs based on flow cytometry. Individual timepoints of sterile ACS (TP1M n=24, TP2M n=25, TP3M n=23, TP4M n=17) compared to CCS (TP0M n=16). Parametric distributed data were analyzed using the Ordinary One-Way ANOVA with correction for multiple comparisons by Dunnett’s test; non-parametric distributed data were analyzed using the Kruskal-Wallis test with correction for multiple comparisons by Dunn’s test. *p≤0.05, **p≤0.01, ***p≤0.001. Illustration of the mean value with SEM.

**Suppl. Fig. 2**

**(a)** Normalized and log-transformed mean expression of differentially expressed genes (bottom x-axis) shown across all clusters (y-axis). Differentially expressed genes calculated using a Wilcoxon rank sum test with Bonferroni-Hochberg correction contrasting expression of one cluster (top x-axis) to expression of the other clusters. **(b)** Scanorama transformed mean expression of PBMC (left) and T cell marker (right) genes per cluster (y-axis) and annotated cell-types (top x-axis). **(c)** Analysis of centered log ratio (CLR) transformed cell type abundance based on scRNA-seq dataset. Individual timepoints of sterile ACS (TP1M n=16, TP2M n=19, TP3M n=16, TP4M n=11) compared to CCS patients (TP0M n=16). Parametric distributed data were analyzed using the Ordinary One-Way ANOVA with correction for multiple comparisons by Dunnett’s test; non-parametric distributed data were analyzed using the Kruskal-Wallis test with correction for multiple comparisons by Dunn’s test. *p≤0.05. Illustration as a Box-Whiskers plot (minimum to maximum).

**Suppl. Fig. 3**

**(a)** Overview data input MOFA: amount of features for each input data dimension (D), amount of samples and missing data highlighted by grey colors (specifying missing data for a specific dimension and sample). **(b)** Factor values of all samples (n=128) in longitudinal comparison for all 20 MOFA factors.

**Suppl. Fig. 4**

**(a)** Comparison of the cell-type annotation strategies on Munich data: x-axis showing clusters and assigned cell-types based on marker genes approach; y-axis showing automated cell-type annotations using the Groningen Azimuth annotation pipeline. **(b)** Comparison of the cell-type annotation strategies on Munich data as umap: left umap showing clusters and assigned cell-types based on marker genes approach; right umap showing automated cell-type annotations using the Groningen Azimuth annotation pipeline on the same umap.

**Suppl. Fig. 5**

**(a)** Overview data input MOFA with automated azimuth annotations: amount of features for each input data dimension (D), amount of samples and missing data highlighted by grey colors (specifying missing data for a specific dimension and sample). **(b)** Factor values of all samples (n=128) in longitudinal comparison for all 20 MOFA factors estimated with azimuth annotations.

**Suppl. Fig. 6**

**(a)** Pearson correlation between **sample factor values** inferred by two different MOFA models: (1) model with manual cell-type cluster annotation (Suppl. Fig. 3) (y-axis) and (2) model with automated azimuth annotations (Suppl. Fig. 5) (x-axis). **(b)** Pearson correlation between **feature factor weights** inferred by two different MOFA models; (1) model with automated azimuth annotations (Suppl. Fig. 5) (y-axis) and (2) model with manual cell-type cluster annotation (Suppl. Fig. 3).

**Suppl. Fig. 7**

**(a)** *Integrative ACS* (Factor 2). Normalized expression values of top 1% features for sterile ACS (TP1M n=17, TP2M n=19, TP3M n=17, TP4M n=12) and CCS (TP0M n=16) patients in longitudinal comparison visualized within heatmap and weight of the features visualized as barplot (+ positive factor weight; - negative factor weight).

**Suppl. Fig. 8**

**(a)** Pearson correlation between **sample factor values** inferred by two different MOFA models: (1) model including clinical features (y-axis) and (2) model without clinical features (x-axis). **(b)** Pearson correlation between **feature factor weights** inferred by two different MOFA models; (1) model including clinical features (y-axis) and (2) model without clinical features (x-axis) per factor.

**Suppl. Fig. 9**

**(a)** Comparison overlap of top 1% of features (D=132) between MOFA model inferred with clinical variables as features and without clinical variables. **(b)** Factor values of all samples (n=128) in longitudinal comparison for all 20 MOFA factors inferred by the MOFA model trained without clinical features. **(c)** Circulating plasma cytokines are shown in comparison of the individual timepoints of sterile ACS (TP1M n=17, TP2M n=19, TP3M n=16, TP4M n=11) with CCS (TP0M n=16). Parametric distributed data were analyzed using the Ordinary One-Way ANOVA with correction for multiple comparisons by Dunnett’s test; non-parametric distributed data were analyzed using the Kruskal-Wallis test with correction for multiple comparisons by Dunn’s test. *p≤0.05. Illustration of the median with interquartile range. In case only the column factor was significant, graphs are marked with a vertical bar on top.

**Suppl. Fig. 10**

**(a)** Circulating plasma cytokines are shown in comparison of the individual timepoints of sterile ACS (TP1M n=17, TP2M n=19, TP3M n=16, TP4M n=11) with CCS (TP0M n=16). Parametric distributed data were analyzed using the Ordinary One-Way ANOVA with correction for multiple comparisons by Dunnett’s test; non-parametric distributed data were analyzed using the Kruskal-Wallis test with correction for multiple comparisons by Dunn’s test. *p≤0.05, **p≤0.01, ***p≤0.001. Graphs in which the column factor was not significant but individual timepoints in the multiple comparison were significant are marked in red. Illustration of the median with interquartile range. In case only the column factor was significant, graphs are marked with a vertical bar on top.

**Suppl. Fig. 11**

**(a)** The components of the cellular interleukin-6 cascade illustrated by REACTOME. The coloring of the components is based on the average factor values of the genes belonging to the pathway and the top 25% of genes on *Integrative ACS*. Adapted from URL: https://reactome.org/content/detail/R-HSA-1059683

**Suppl. Fig. 12**

**(a)** Circulating plasma cytokines are shown in comparison of the individual timepoints of ACS subtypes (sterile ACS TP1M n=17, TP2M n=19, TP3M n=16, TP4M n=11; ACS acquiring hospital infection TP1M n=5, TP2M n=5, TP3M n=5, TP4M n=4; ACS with delayed recanalization after vessel occlusion TP1M n=4, TP2M n=2, TP3M n=2, TP4M n=2). Dataset was analyzed using the Mixed-effects analysis with correction for multiple comparisons by Tukey’s test. *p≤0.05, **p≤0.01, ***p≤0.001. Graphs in which the column factor was not significant but individual timepoints in the multiple comparison were significant are marked in red. Illustration of the median with interquartile range.In case only the column factor was significant, graphs are marked with a vertical bar on top.

**Suppl. Fig. 13**

**(a)** Circulating plasma cytokines are shown in comparison of the individual timepoints of ACS subtypes (sterile ACS TP1M n=17, TP2M n=19, TP3M n=16, TP4M n=11; ACS acquiring hospital infection TP1M n=5, TP2M n=5, TP3M n=5, TP4M n=4; ACS with delayed recanalization after vessel occlusion TP1M n=4, TP2M n=2, TP3M n=2, TP4M n=2). Dataset was analyzed using the Mixed-effects analysis with correction for multiple comparisons by Tukey’s test. *p≤0.05, **p≤0.01, ***p≤0.001. Graphs in which the column factor was not significant but individual timepoints in the multiple comparison were significant are marked in red. Illustration of the median with interquartile range. **(b)** ROC AUC. Comparison of the predictive power of *Integrative ACS Outcome* (Factor 4) (n=19) and mean and max CK levels (n=19) and mean and max CRP troponin levels (n=19) based on the complete longitudinal values.

**Suppl. Fig. 14**

**(a)** *Integrative ACS Outcome* (Factor 4) Normalized expression values of top 1% features in longitudinal comparison visualized within heatmap and weight of the features visualized as barplot, all samples included (n=128). Divided by outcome; ’NA’ in case no EF value has been available for the ACS samples (n=7) and for CCS and non-CCS samples (+ positive factor weight; - negative factor weight).

**Suppl. Fig. 15**

**(a)** Circulating plasma cytokines are shown in comparison of good and poor outcome (good outcome: TP1M n=13, TP2M n=14, TP3M n=13, TP4M n=10; poor Outcome: TP1M n=6, TP2M n=7, TP3M n=7, TP4M n=2). The dataset was analyzed using the Mixed-effects analysis with correction for multiple comparisons by Šidák test. *p≤0.05. Graphs in which the column factor was not significant but individual timepoints in the multiple comparison were significant are marked in red. In case only the column factor was significant, graphs are marked with a vertical bar on top. Illustration of the median with interquartile range. Illustration of the median with interquartile range.

**Suppl. Fig. 16**

**(a)** Circulating plasma cytokines are shown in comparison of good and poor outcome (good outcome: TP1M n=13, TP2M n=14, TP3M n=13, TP4M=10; poor Outcome: TP1M n=6, TP2M n=7, TP3M n=7, TP4M n=2). The dataset was analyzed using the Mixed-effects analysis with correction for multiple comparisons by Šidák test. *p≤0.05, **p≤0.05. Graphs in which the column factor was not significant but individual timepoints in the multiple comparison were significant are marked in red. Illustration of the median with interquartile range. Illustration of the median with interquartile range. In case only the column factor was significant, graphs are marked with a vertical bar on top.

**Suppl. Fig.17**

**(a)** Normalized expression values of selected top features of *Integrative ACS Outcome* (Factor 4) in longitudinal comparison for samples classified with good or poor outcome (NK cells (cluster 3): good outcome: TP1M n=13, TP2M n=13, TP3M n=14, TP4M=11; poor Outcome: TP1M n=4, TP2M n=6, TP3M n=6, TP4M n=3 Cytokines: good outcome: TP1M n=13, TP2M n=14, TP3M n=13, TP4M=10; poor Outcome: TP1M n=6, TP2M n=7, TP3M n=7, TP4M n=4 Plasma Proteomics: good outcome: TP1M n=10, TP2M n=13, TP3M n=13, TP4M=10; poor Outcome: TP1M n=6, TP2M n=7, TP3M n=7, TP4M n=4).

**Suppl. Fig. 18**

**(a)** *Integrative CCS* (Factor 1). Normalized expression values of top 1% features in longitudinal comparison visualized within heatmap and weight of the features visualized as barplot, all samples included (n=128) (+ positive factor weight; - negative factor weight).

**Suppl. Fig. 19**

**(a)** Normalized expression values of selected top features of *Integrative CCS* (Factor 1) in comparison for samples classified CCS (TP0M n=16) and non CCS (TP0M n=17)**. (b)** Factor weights of the top 10 ligands with the highest factor weight on *Integrative CCS* (Factor 1). **(c)** Normalized gene expression values of selected top features comparing patients with CCS (TP0M n=16) and non-CCS (TP0M n=17). **(d)** Spearman correlations (|cor| ≥0.4) between ligand and target genes across all samples (n=128). Target genes selected as top 1% of features with **negative** feature weight on *Integrative CCS* (Factor 1). Ligands selected based on minimum regulatory potential score of 0.0012 on those targets according to the NicheNet Model (corresponding to 97% quantile of regulatory potential score). **(e)** Spearman correlations (|cor| ≥0.4) between ligand and target genes across all samples (n=128). Target genes selected as top 1% of features with **positive** feature weight on *Integrative CCS* (Factor 1). Ligands selected based on minimum regulatory potential score of 0.0012 on those targets according to the NicheNet Model (corresponding to 97% quantile of regulatory potential score). **(f)** Normalized expression values of selected top features of *Integrative CCS* (Factor 1) in comparison for samples classified CCS (TP0M n=16) and non-CCS (TP0M n=17)**. (g)** Correlation score of selected examples of circoplots (d,e).

**Suppl. Fig. 20**

**(a)** Circulating plasma cytokines are shown in comparison of CCS (TP0M n=16) with non-CCS (TP0M n=18) patients. Parametric data were analyzed using an unpaired t-test, non-parametric data were tested using the Mann-Whitney test. *p≤0.05, **p≤0.01. Illustration of the median with interquartile range.

**Suppl. Fig. 21**

**(a)** Circulating plasma cytokines are shown are shown in comparison of CCS (TP0M n=16) with non-CCS (TP0M n=18) patients. Parametric data were analyzed using an unpaired t-test, non-parametric data were tested using the Mann-Whitney test. *p≤0.05. Illustration of the median with interquartile range.

