## Supplementary Figures for "Multi-Omic Factor Analysis uncovers immunological signatures with pathophysiologic and clinical implications in coronary syndromes"

Suppl. Fig. 1

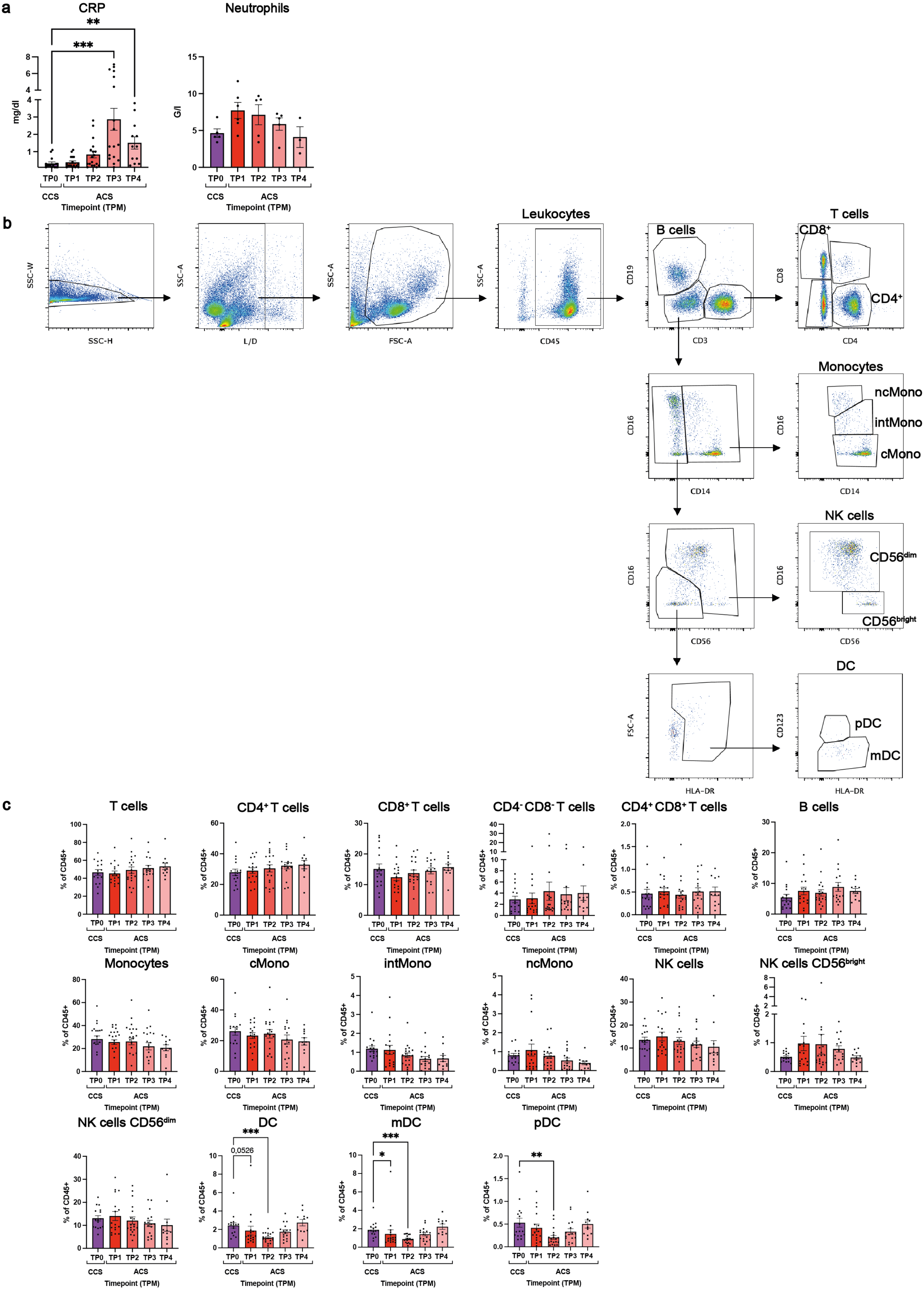
a
CRP
Neutrophils
mg/dl
G/l
TP0 TP1 TP2 TP3 TP4
CCS
ACS
Timepoint (TPM)
b
Leukocytes
B cells
T cells
CD8+
CD4+
Monocytes
ncMono
intMono
cMono
NK cells
CD56dim
CD56bright
DC
pDC
mDC
SSC-W
SSC-H
SSC-A
L/D
FSC-A
CD45
CD19
CD3
CD8
CD4
CD16
CD14
CD56
FSC-A
HLA-DR
CD123
c
T cells
CD4+ T cells
CD8+ T cells
CD4- CD8- T cells
CD4+ CD8+ T cells
B cells
Monocytes
cMono
intMono
ncMono
NK cells
NK cells CD56bright
NK cells CD56dim
DC
mDC
pDC
% of CD45+
0,0526

Suppl. Fig. 2

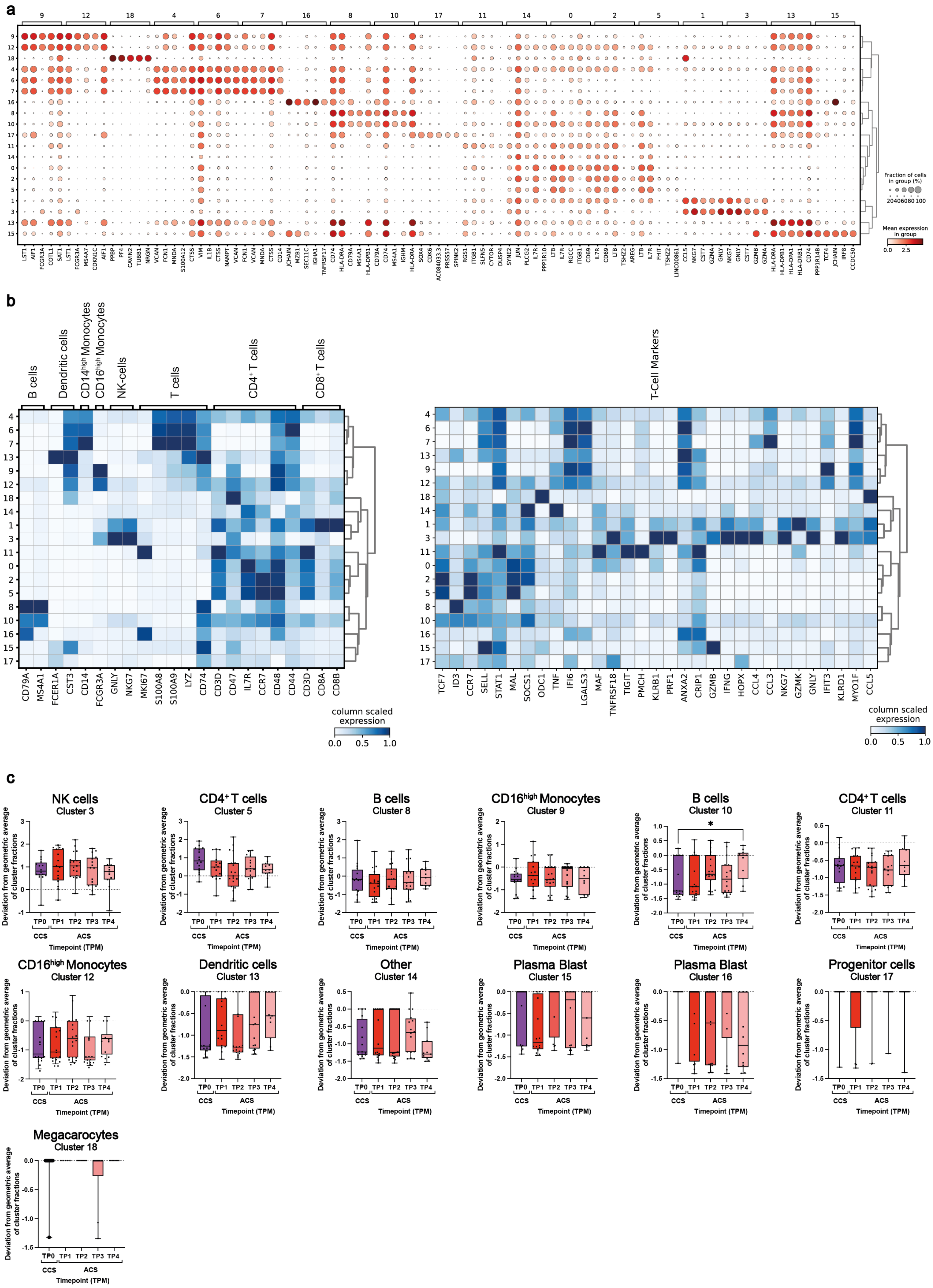
a
Fraction of cells in group (%)
20 40 60 80 100
Mean expression in group
0.0 2.5
b
B cells
Dendritic cells
CD14^high Monocytes
CD16^high Monocytes
NK-cells
T cells
CD4⁺ T cells
CD8⁺ T cells
CD79A MS4A1 FCER1A CST3 CD14 FCGR3A GNLY NKG7 MKI67 S100A8 S100A9 LYZ CD74 CD3D CD47 IL7R CCR7 CD48 CD44 CD3D CD8A CD8B
column scaled expression
0.0 0.5 1.0
T-Cell Markers
TCF7 ID3 CCR7 SELL STAT1 MAL SOCS1 ODC1 TNF IFI6 LGALS3 MAF TNFRSF18 TIGIT PMCH KLRB1 PRF1 ANXA2 CRIP1 GZMB IFNG HOPX CCL4 CCL3 NKG7 GZMK GNLY IFIT3 KLRD1 MYO1F CCL5
column scaled expression
0.0 0.5 1.0
c
NK cells
Cluster 3
CD4⁺ T cells
Cluster 5
B cells
Cluster 8
CD16^high Monocytes
Cluster 9
B cells
Cluster 10
CD4⁺ T cells
Cluster 11
CD16^high Monocytes
Cluster 12
Dendritic cells
Cluster 13
Other
Cluster 14
Plasma Blast
Cluster 15
Plasma Blast
Cluster 16
Progenitor cells
Cluster 17
Megacarocytes
Cluster 18
Deviation from geometric average of cluster fractions
TP0 TP1 TP2 TP3 TP4
CCS ACS
Timepoint (TPM)

Suppl. Fig. 3

a

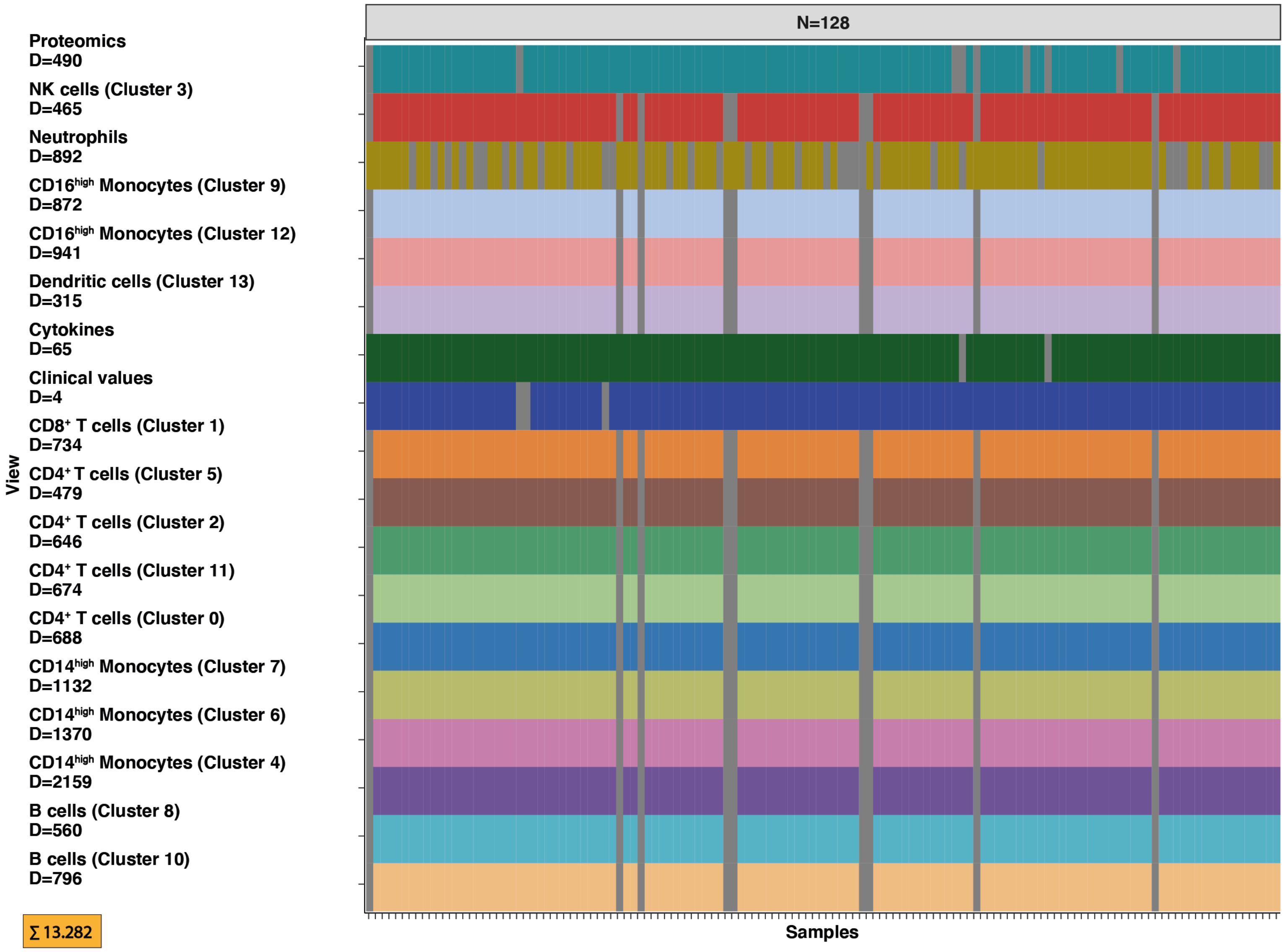


b

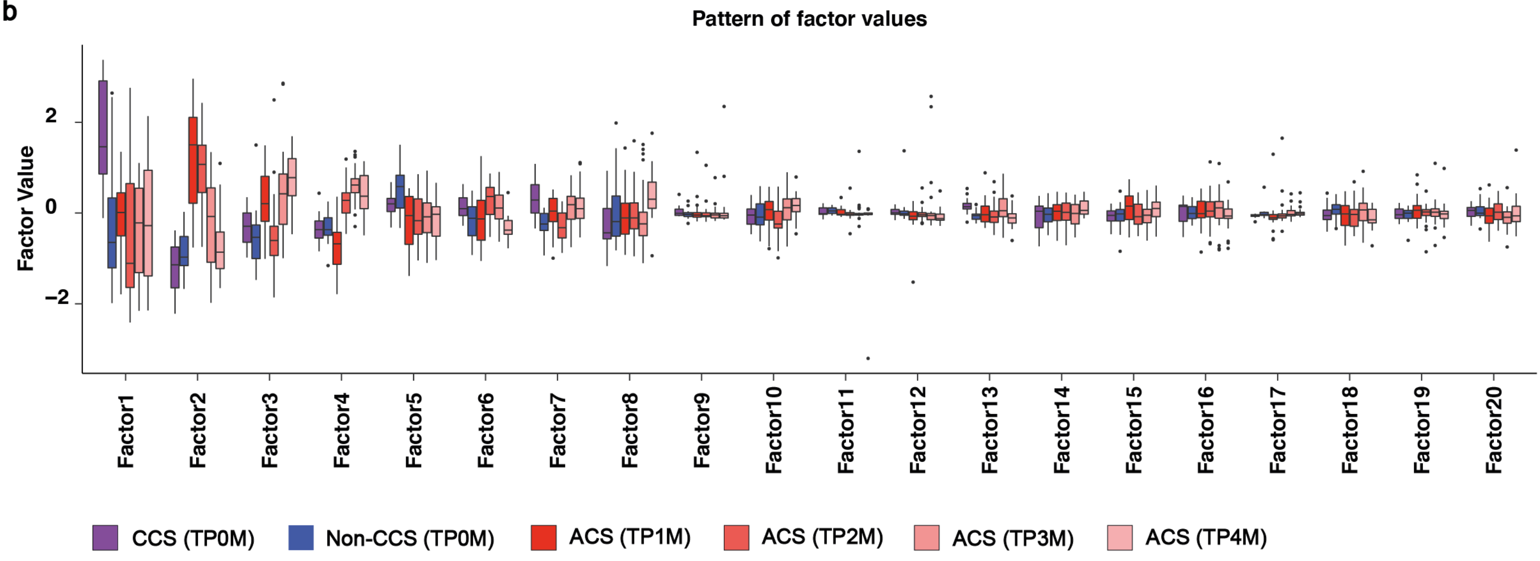

Suppl. Fig. 4

**a**

predicted.celltype.l2 (rows) × Cluster cell type Scanorama (columns)

| predicted.celltype.l2 | CD4$^+$ T cells (Cluster 0) | CD8$^+$ T cells (Cluster 1) | B cells (Cluster 10) | T$_{regs}$ cells (Cluster 11) | CD16$^{high}$ Monocytes (Cluster 12) | Dendritic cells (Cluster 13) | Other cells (Cluster 14) | CD4$^+$ T cells (Cluster 2) | NK cells (Cluster 3) | CD14$^{high}$ Monocytes (Cluster 4) | CD4$^+$ T cells (Cluster 5) | CD14$^{high}$ Monocytes (Cluster 6) | CD14$^{high}$ Monocytes (Cluster 7) | B cells (Cluster 8) | CD16$^{high}$ Monocytes (Cluster 9) | Total |
|---|---|---|---|---|---|---|---|---|---|---|---|---|---|---|---|---|
| Total | 24806 | 19754 | 2897 | 2769 | 1934 | 1365 | 1345 | 19071 | 15670 | 14625 | 12514 | 11202 | 7836 | 7399 | 3114 | |
| Treg | 437 | 34 | 24 | 785 | 1 | | 84 | 701 | | 18 | 275 | | | 1 | | 2362 |
| NK | | 1379 | 53 | 1 | | 3 | 41 | | 14185 | 35 | | | | | | 15699 |
| MAIT | 32 | 1085 | 11 | | 4 | | 8 | 1 | 15 | 20 | 1 | | | | | 1177 |
| gdT | 5 | 1652 | 7 | 7 | 2 | | 8 | 30 | 176 | 9 | 5 | | | | | 1901 |
| cDC2 | | | | | | 1171 | | | | 80 | | 58 | 1 | | | 1314 |
| CD8 TEM | 87 | 9476 | 118 | 71 | 19 | | 174 | 21 | 502 | 126 | 9 | | | | | 10611 |
| CD8 Naive | 40 | 5 | 8 | 13 | 2 | | 8 | 1444 | | 9 | 1083 | | | | | 2612 |
| CD4 TEM | 2638 | 2318 | 62 | 101 | 13 | | 143 | 826 | 4 | 56 | 131 | | 1 | 1 | | 6296 |
| CD4 TCM | 21149 | 476 | 286 | 1571 | 90 | 1 | 626 | 14323 | 2 | 363 | 6201 | | 2 | 1 | | 45102 |
| CD4 Naive | 36 | 15 | 17 | 2 | 2 | | 133 | 1524 | | 9 | 4752 | | | 2 | | 6495 |
| CD4 CTL | | 2262 | 19 | 5 | 2 | | 6 | | 43 | 9 | | | | | | 2346 |
| CD16 Mono | | | | | 1557 | | | | | 54 | | 123 | 23 | | 2920 | 4680 |
| CD14 Mono | | 1 | | | 129 | 89 | 19 | 1 | | 13466 | | 11012 | 7789 | | 156 | 32727 |
| B naive | | | 597 | | 13 | | 2 | | | 11 | | | | 2935 | | 3558 |
| B memory | | | 744 | | 17 | 1 | 2 | | | 49 | | | | 1175 | | 2010 |
| B intermediate | | | 889 | | 28 | 2 | 5 | | | 64 | | | | 3270 | | 4261 |

Colour scale: 10000, 20000, 30000, 40000

**b**

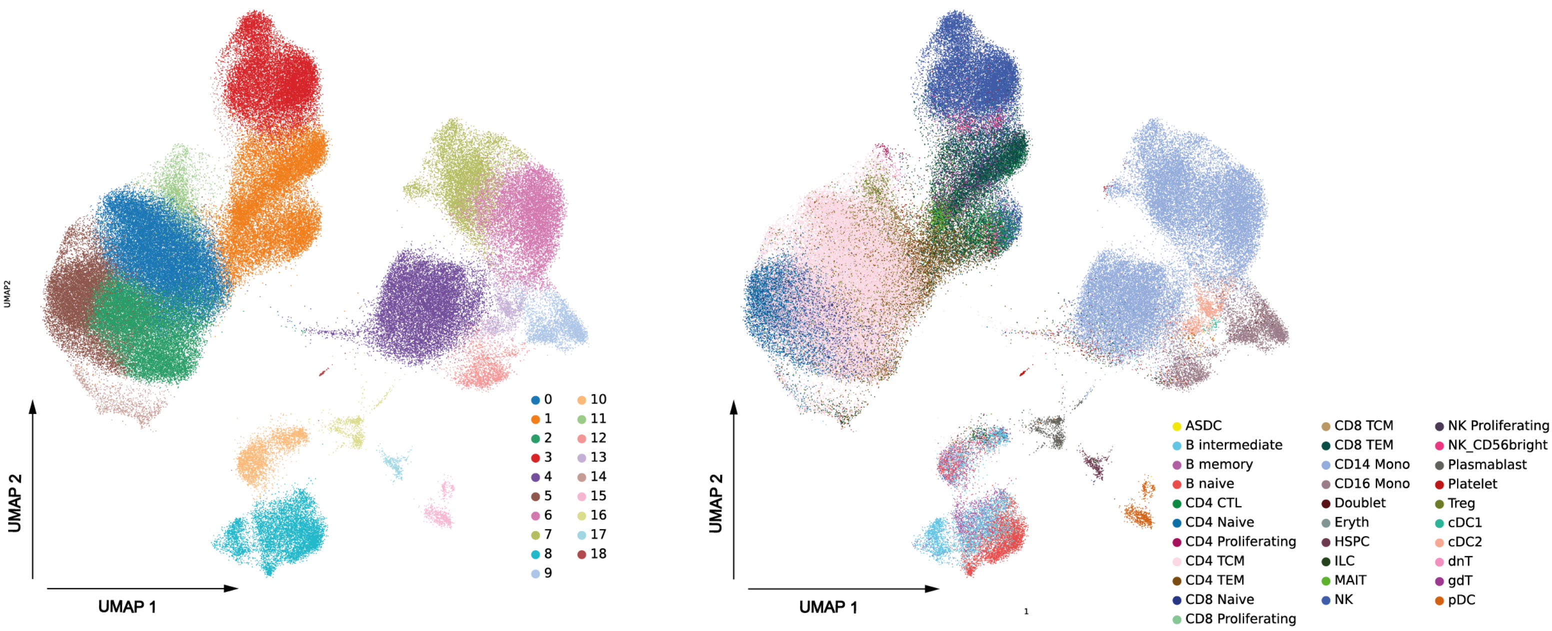

Suppl. Fig. 5

a

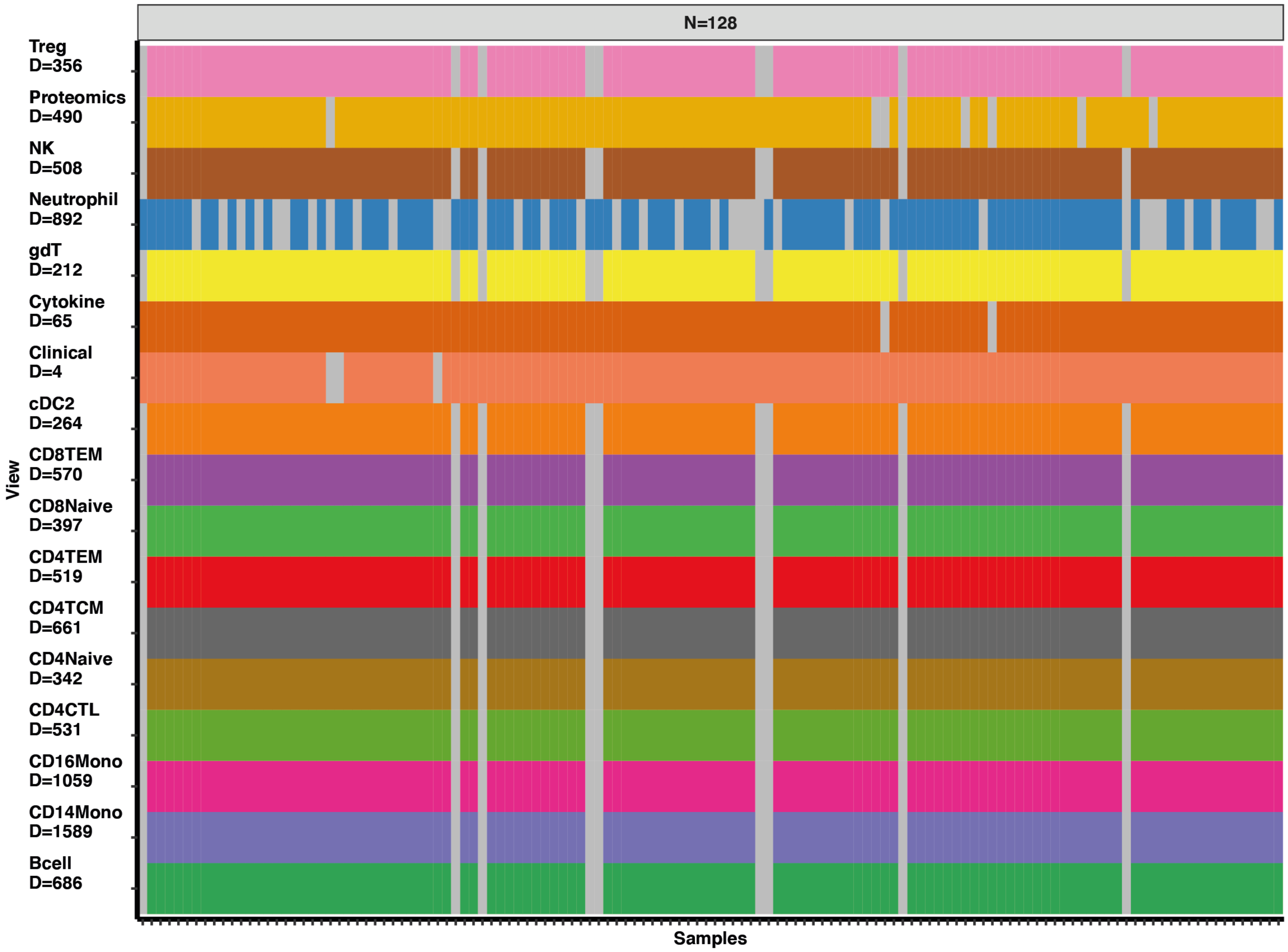


b

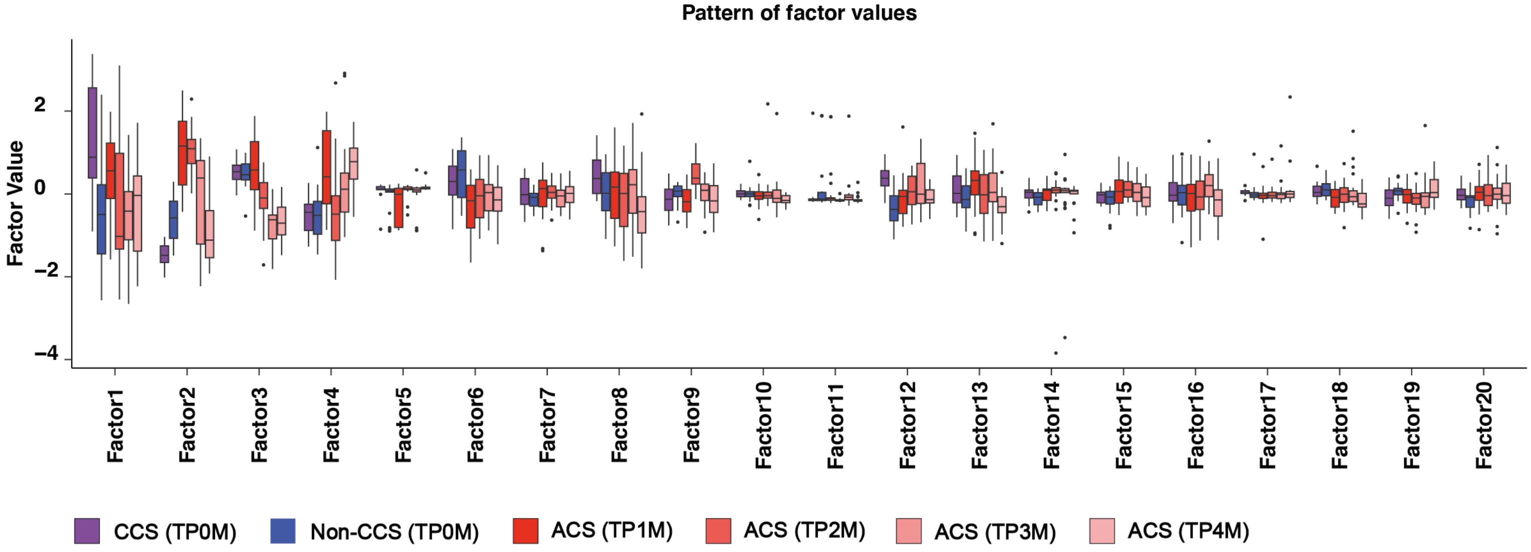

**Suppl. Fig. 6**

**a** **Sample factor values**

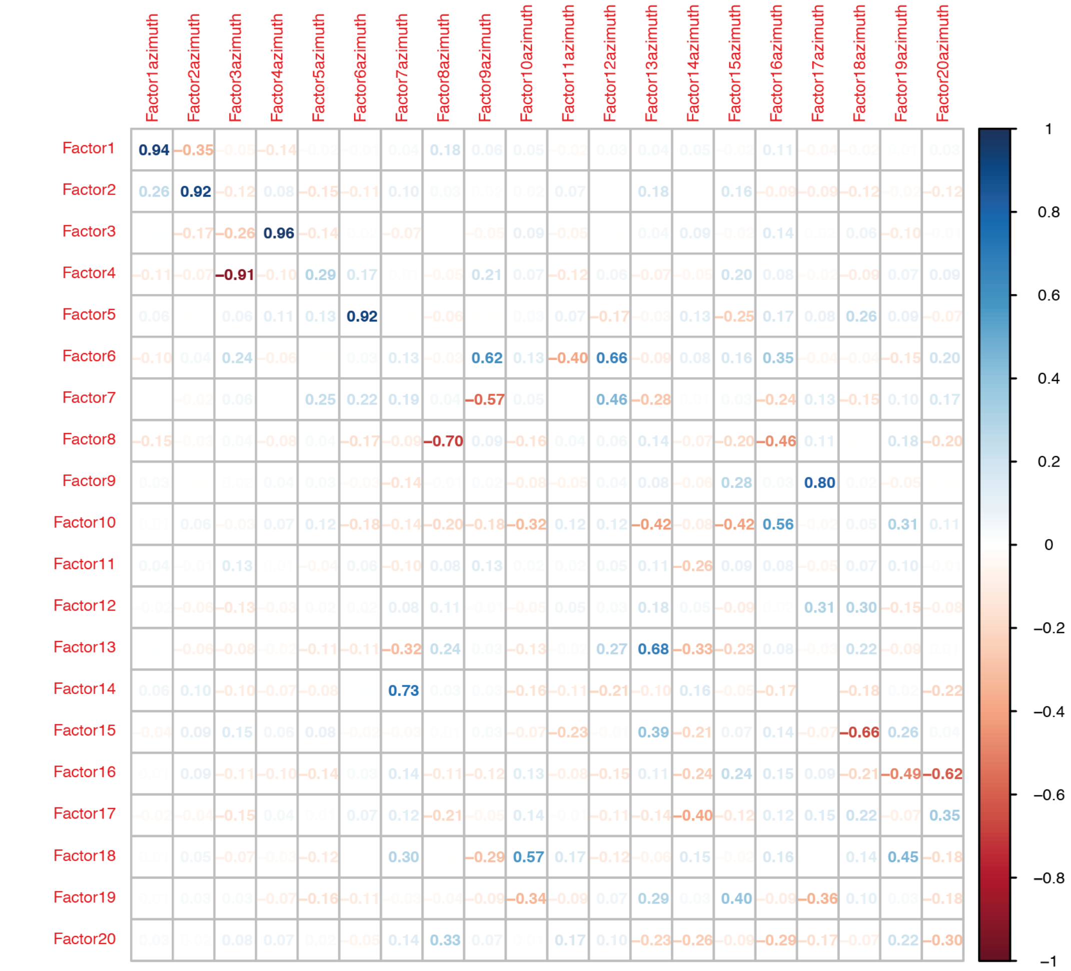


**b** **Feature factor weights**

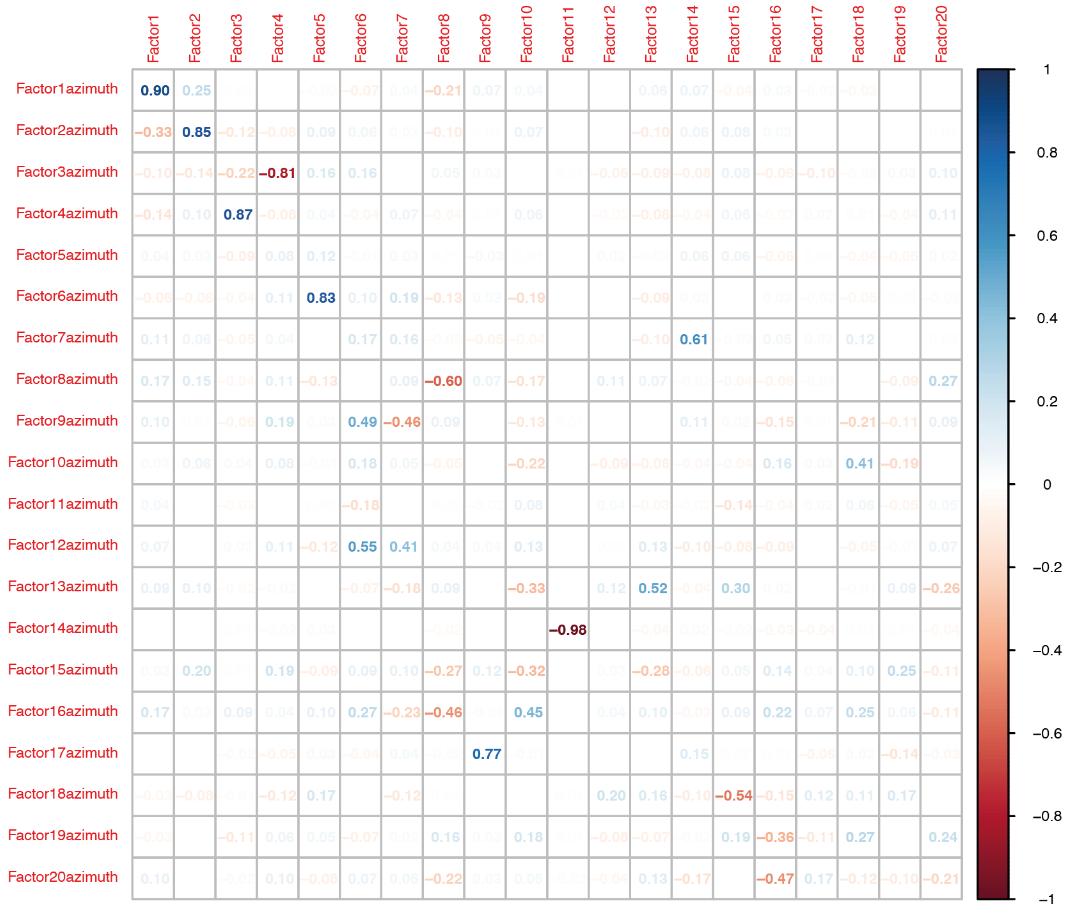

Suppl. Fig. 7

a

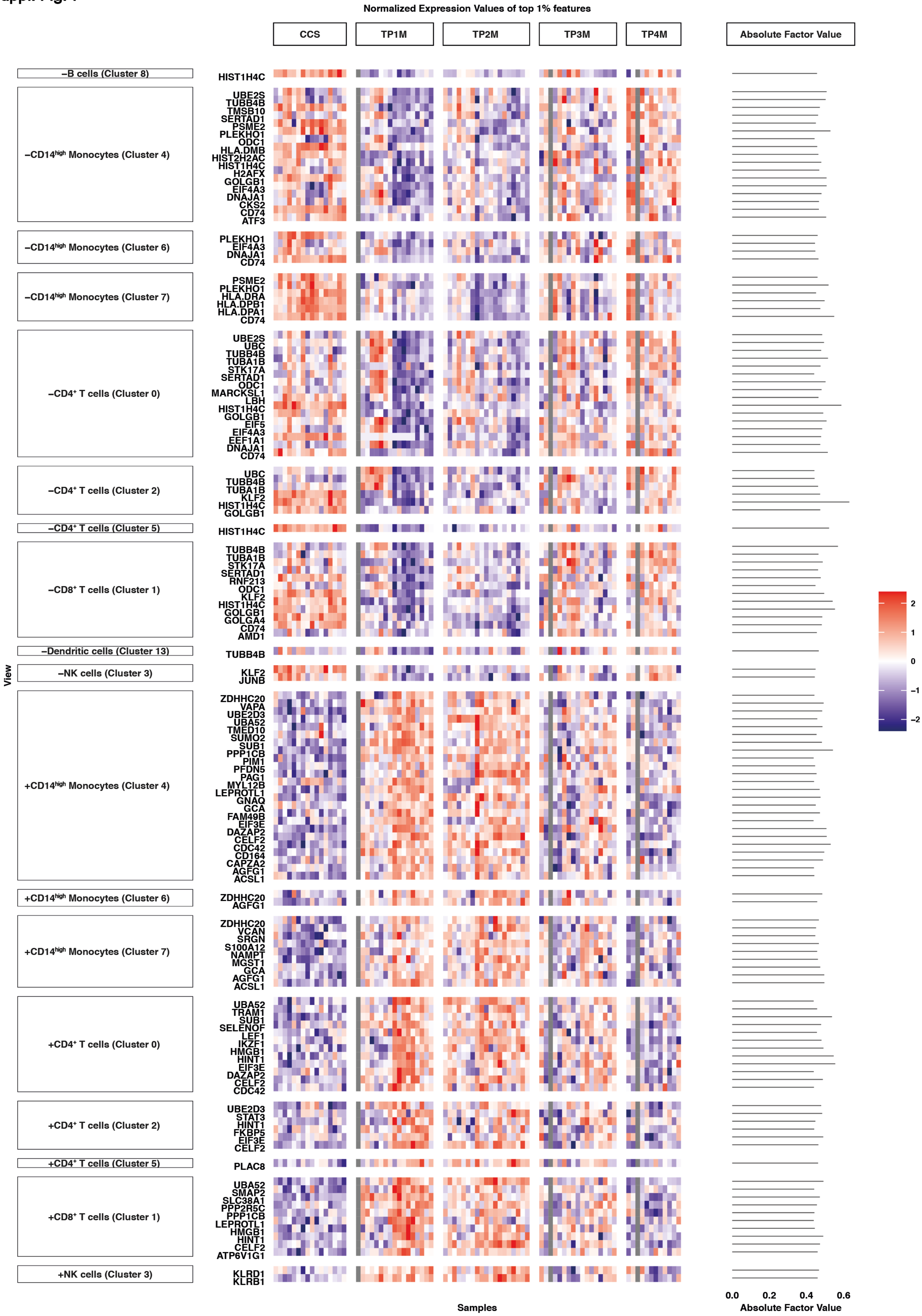
Normalized Expression Values of top 1% features
CCS
TP1M
TP2M
TP3M
TP4M
Absolute Factor Value
−B cells (Cluster 8)
HIST1H4C
−CD14high Monocytes (Cluster 4)
UBE2S
TUBB4B
TMSB10
SERTAD1
PSME2
PLEKHO1
ODC1
HLA.DMB
HIST2H2AC
HIST1H4C
H2AFX
GOLGB1
EIF4A3
DNAJA1
CKS2
CD74
ATF3
−CD14high Monocytes (Cluster 6)
PLEKHO1
EIF4A3
DNAJA1
CD74
−CD14high Monocytes (Cluster 7)
PSME2
PLEKHO1
HLA.DRA
HLA.DPB1
HLA.DPA1
CD74
−CD4+ T cells (Cluster 0)
UBE2S
UBC
TUBB4B
TUBA1B
STK17A
SERTAD1
ODC1
MARCKSL1
LBH
HIST1H4C
GOLGB1
EIF5
EIF4A3
EEF1A1
DNAJA1
CD74
−CD4+ T cells (Cluster 2)
UBC
TUBB4B
TUBA1B
KLF2
HIST1H4C
GOLGB1
−CD4+ T cells (Cluster 5)
HIST1H4C
−CD8+ T cells (Cluster 1)
TUBB4B
TUBA1B
STK17A
SERTAD1
RNF213
ODC1
KLF2
HIST1H4C
GOLGB1
GOLGA4
CD74
AMD1
−Dendritic cells (Cluster 13)
TUBB4B
−NK cells (Cluster 3)
KLF2
JUNB
+CD14high Monocytes (Cluster 4)
ZDHHC20
VAPA
UBE2D3
UBA52
TMED10
SUMO2
SUB1
PPP1CB
PIM1
PFDN5
PAG1
MYL12B
LEPROTL1
GNAQ
GCA
FAM49B
EIF3E
DAZAP2
CELF2
CDC42
CD164
CAPZA2
AGFG1
ACSL1
+CD14high Monocytes (Cluster 6)
ZDHHC20
AGFG1
+CD14high Monocytes (Cluster 7)
ZDHHC20
VCAN
SRGN
S100A12
NAMPT
MGST1
GCA
AGFG1
ACSL1
+CD4+ T cells (Cluster 0)
UBA52
TRAM1
SUB1
SELENOF
LEF1
IKZF1
HMGB1
HINT1
EIF3E
DAZAP2
CELF2
CDC42
+CD4+ T cells (Cluster 2)
UBE2D3
STAT3
HINT1
FKBP5
EIF3E
CELF2
+CD4+ T cells (Cluster 5)
PLAC8
+CD8+ T cells (Cluster 1)
UBA52
SMAP2
SLC38A1
PPP2R5C
PPP1CB
LEPROTL1
HMGB1
HINT1
CELF2
ATP6V1G1
+NK cells (Cluster 3)
KLRD1
KLRB1
View
Samples
0.0
0.2
0.4
0.6
Absolute Factor Value
2
1
0
−1
−2

**Suppl. Fig. 8**

**a** **Sample factor values**

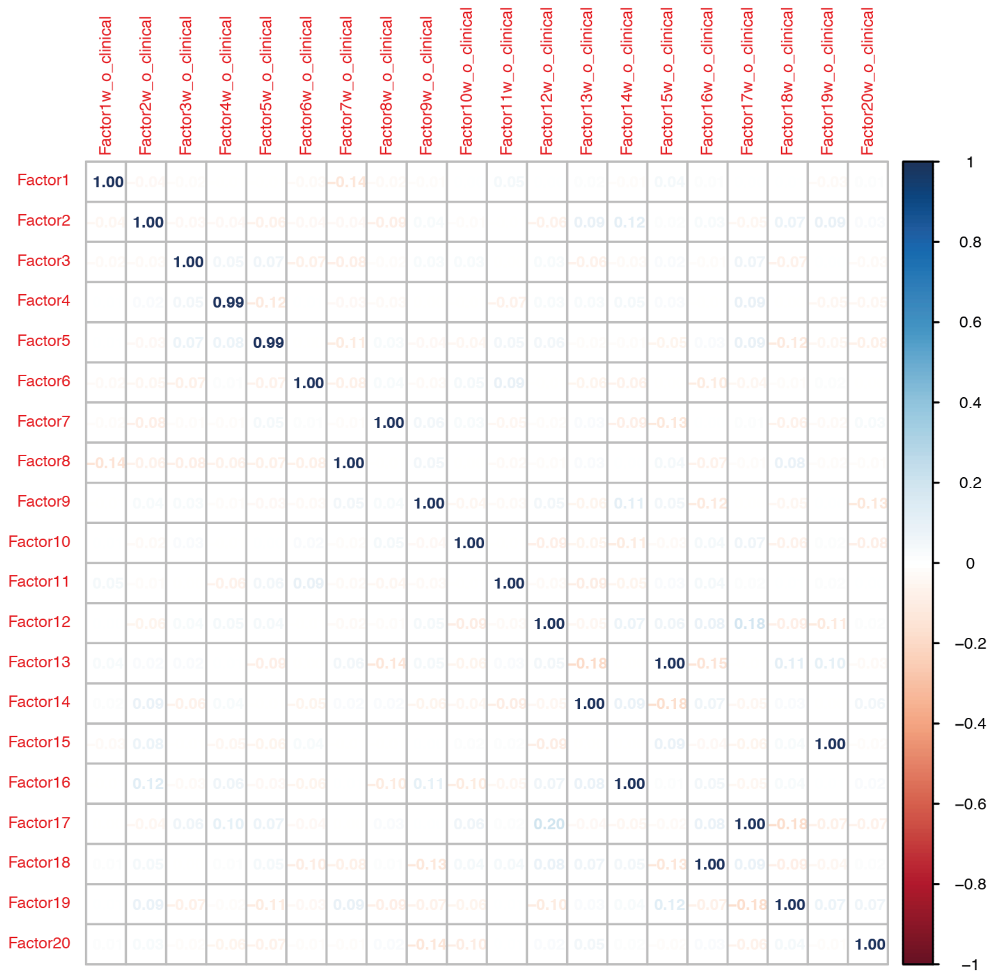


**b** **Feature factor weights**

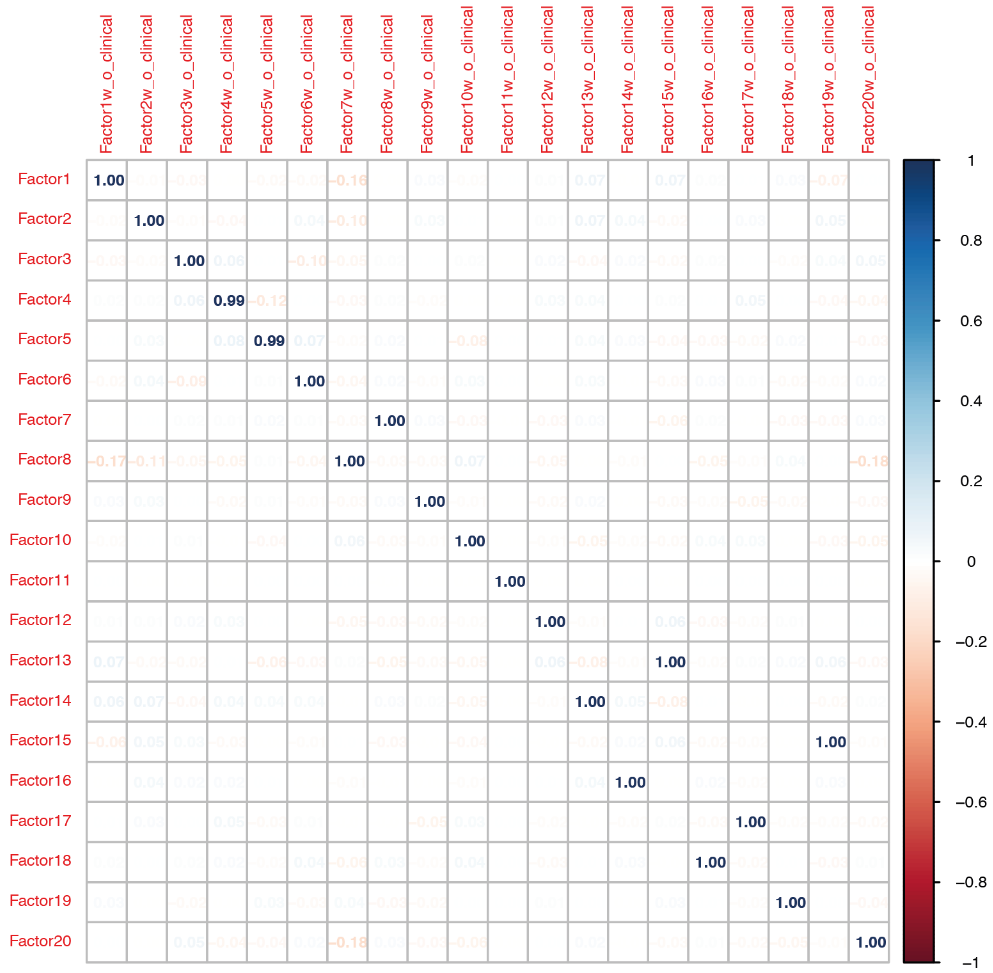

Suppl. Fig. 9

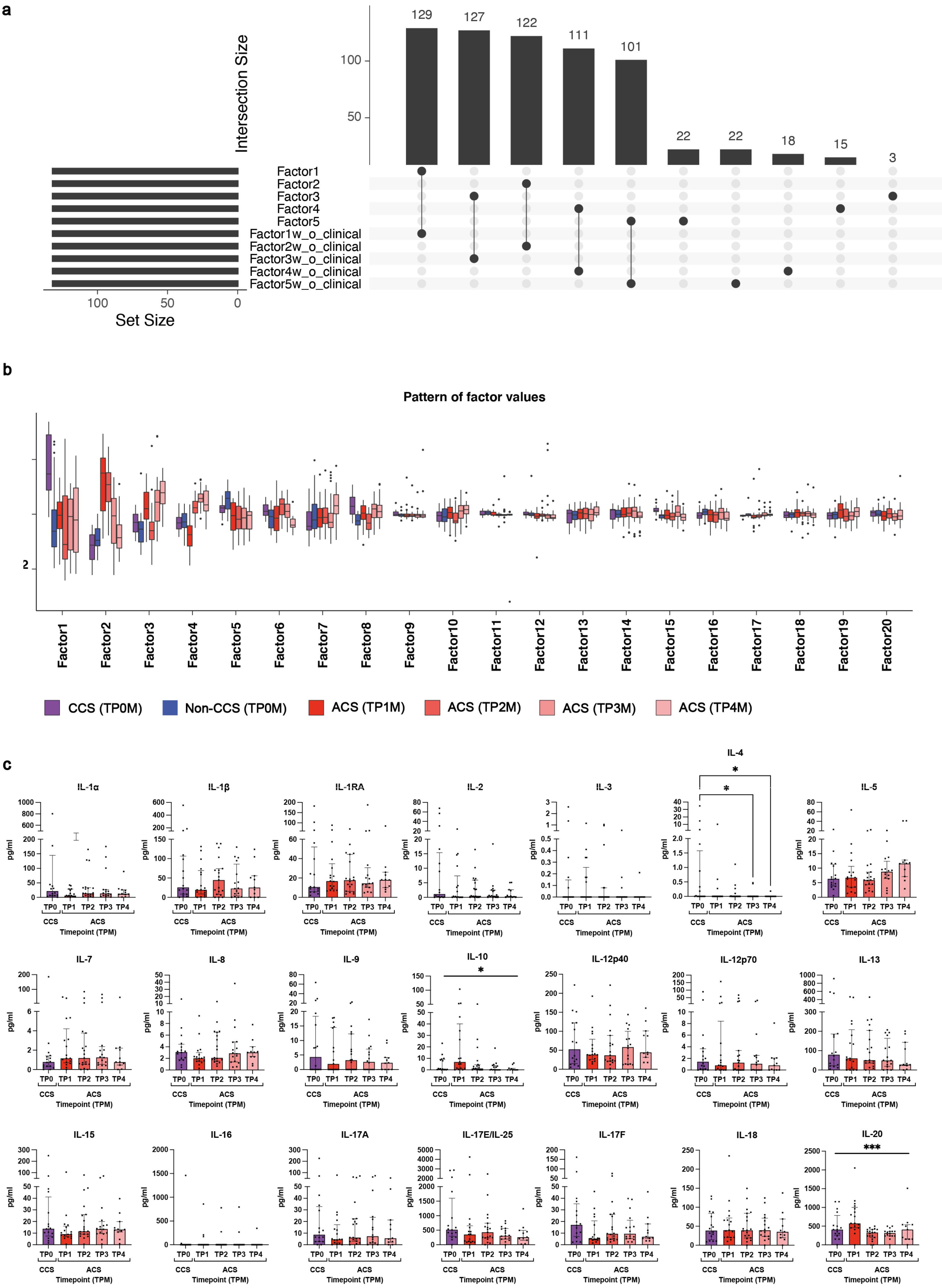

a
Intersection Size
129
127
122
111
101
22
22
18
15
3
100
50
Factor1
Factor2
Factor3
Factor4
Factor5
Factor1w_o_clinical
Factor2w_o_clinical
Factor3w_o_clinical
Factor4w_o_clinical
Factor5w_o_clinical
100
50
0
Set Size
b
Pattern of factor values
2
Factor1
Factor2
Factor3
Factor4
Factor5
Factor6
Factor7
Factor8
Factor9
Factor10
Factor11
Factor12
Factor13
Factor14
Factor15
Factor16
Factor17
Factor18
Factor19
Factor20
CCS (TP0M)
Non-CCS (TP0M)
ACS (TP1M)
ACS (TP2M)
ACS (TP3M)
ACS (TP4M)
c
IL-1α
IL-1β
IL-1RA
IL-2
IL-3
IL-4
IL-5
IL-7
IL-8
IL-9
IL-10
IL-12p40
IL-12p70
IL-13
IL-15
IL-16
IL-17A
IL-17E/IL-25
IL-17F
IL-18
IL-20
pg/ml
TP0
TP1
TP2
TP3
TP4
CCS
ACS
Timepoint (TPM)
*
***

### Suppl. Fig. 10

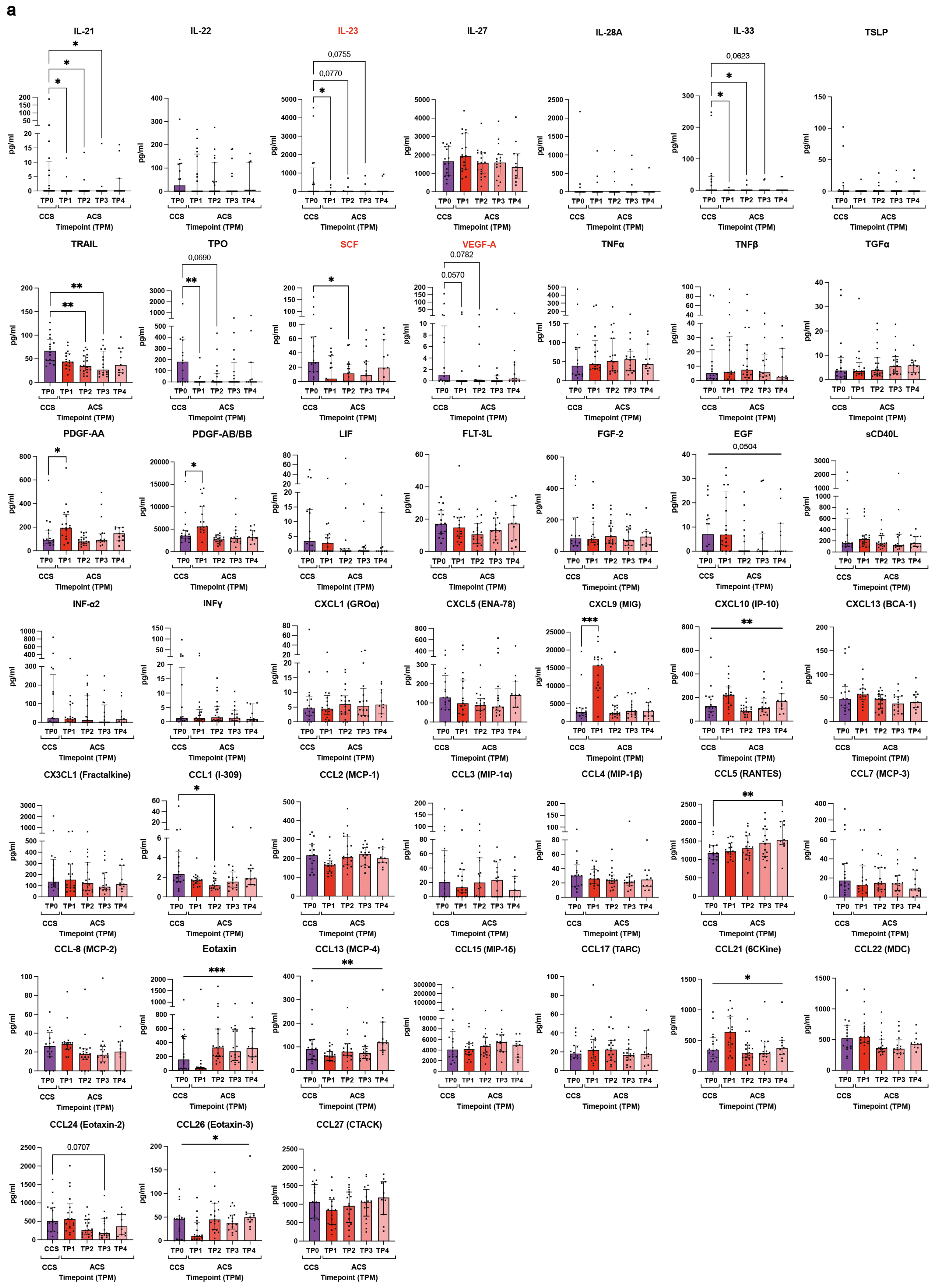
a
IL-21
IL-22
IL-23
0,0755
0,0770
IL-27
IL-28A
IL-33
0,0623
TSLP
TRAIL
TPO
0,0690
SCF
VEGF-A
0.0782
0.0570
TNFα
TNFβ
TGFα
PDGF-AA
PDGF-AB/BB
LIF
FLT-3L
FGF-2
EGF
0,0504
sCD40L
INF-α2
INFγ
CXCL1 (GROα)
CXCL5 (ENA-78)
CXCL9 (MIG)
CXCL10 (IP-10)
CXCL13 (BCA-1)
CX3CL1 (Fractalkine)
CCL1 (I-309)
CCL2 (MCP-1)
CCL3 (MIP-1α)
CCL4 (MIP-1β)
CCL5 (RANTES)
CCL7 (MCP-3)
CCL-8 (MCP-2)
Eotaxin
CCL13 (MCP-4)
CCL15 (MIP-1δ)
CCL17 (TARC)
CCL21 (6CKine)
CCL22 (MDC)
CCL24 (Eotaxin-2)
0.0707
CCL26 (Eotaxin-3)
CCL27 (CTACK)
pg/ml
TP0
TP1
TP2
TP3
TP4
CCS
ACS
Timepoint (TPM)

**Suppl. Fig. 11**

**a**

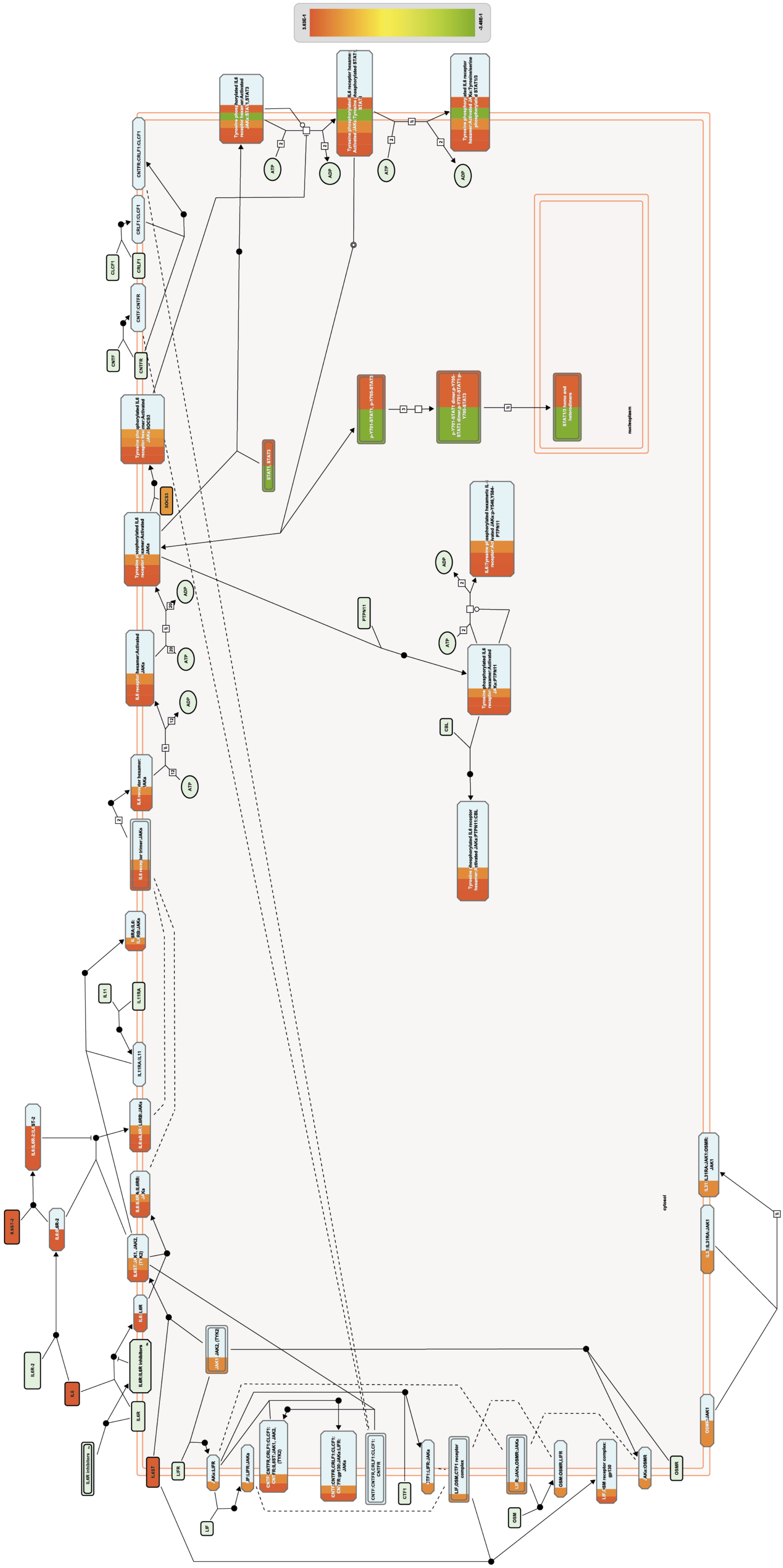

### Suppl. Fig. 12

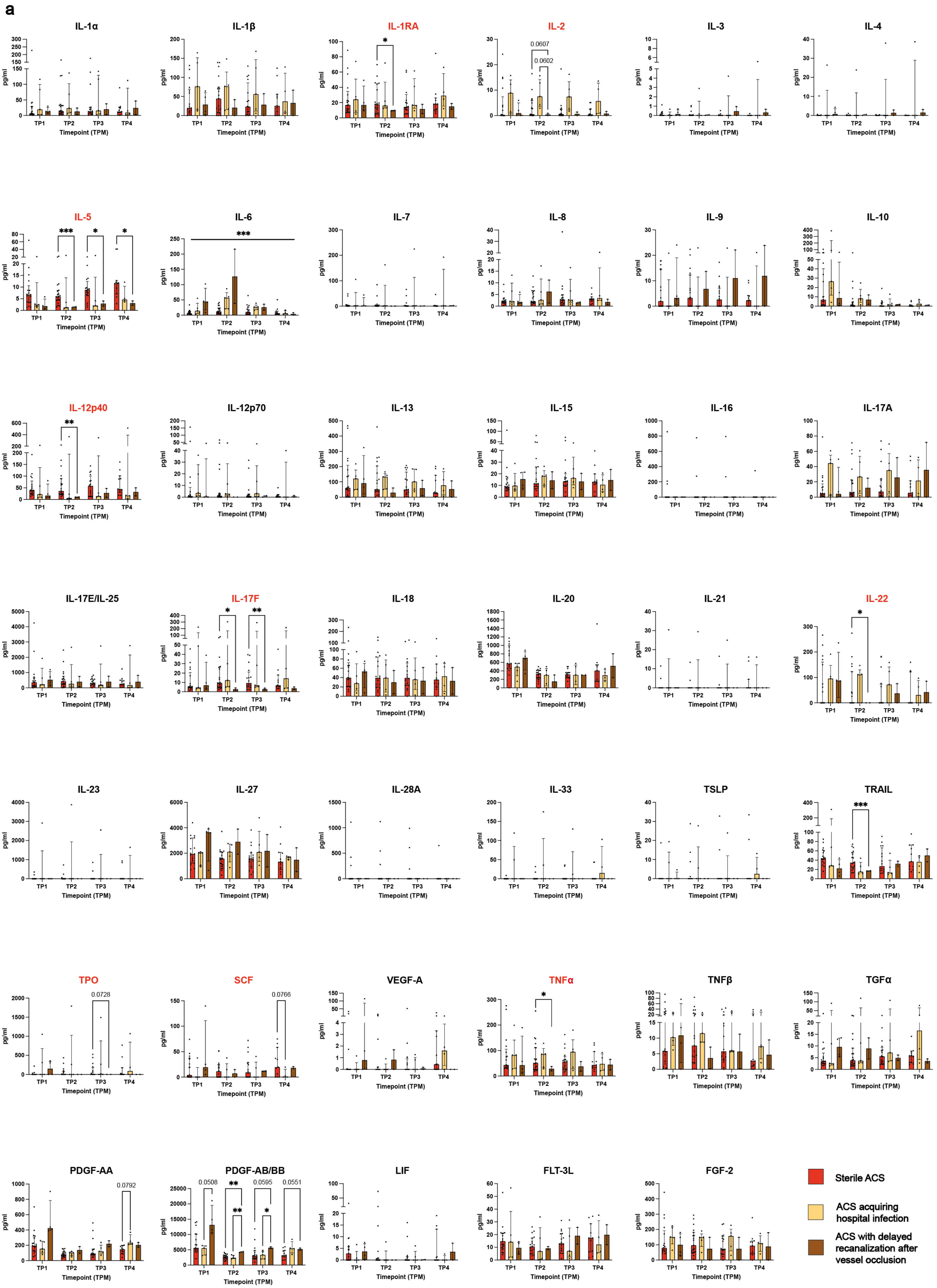
a
IL-1α
IL-1β
IL-1RA
IL-2
IL-3
IL-4
IL-5
IL-6
IL-7
IL-8
IL-9
IL-10
IL-12p40
IL-12p70
IL-13
IL-15
IL-16
IL-17A
IL-17E/IL-25
IL-17F
IL-18
IL-20
IL-21
IL-22
IL-23
IL-27
IL-28A
IL-33
TSLP
TRAIL
TPO
SCF
VEGF-A
TNFα
TNFβ
TGFα
PDGF-AA
PDGF-AB/BB
LIF
FLT-3L
FGF-2
pg/ml
Timepoint (TPM)
TP1
TP2
TP3
TP4
Sterile ACS
ACS acquiring hospital infection
ACS with delayed recanalization after vessel occlusion

Suppl. Fig. 13

a

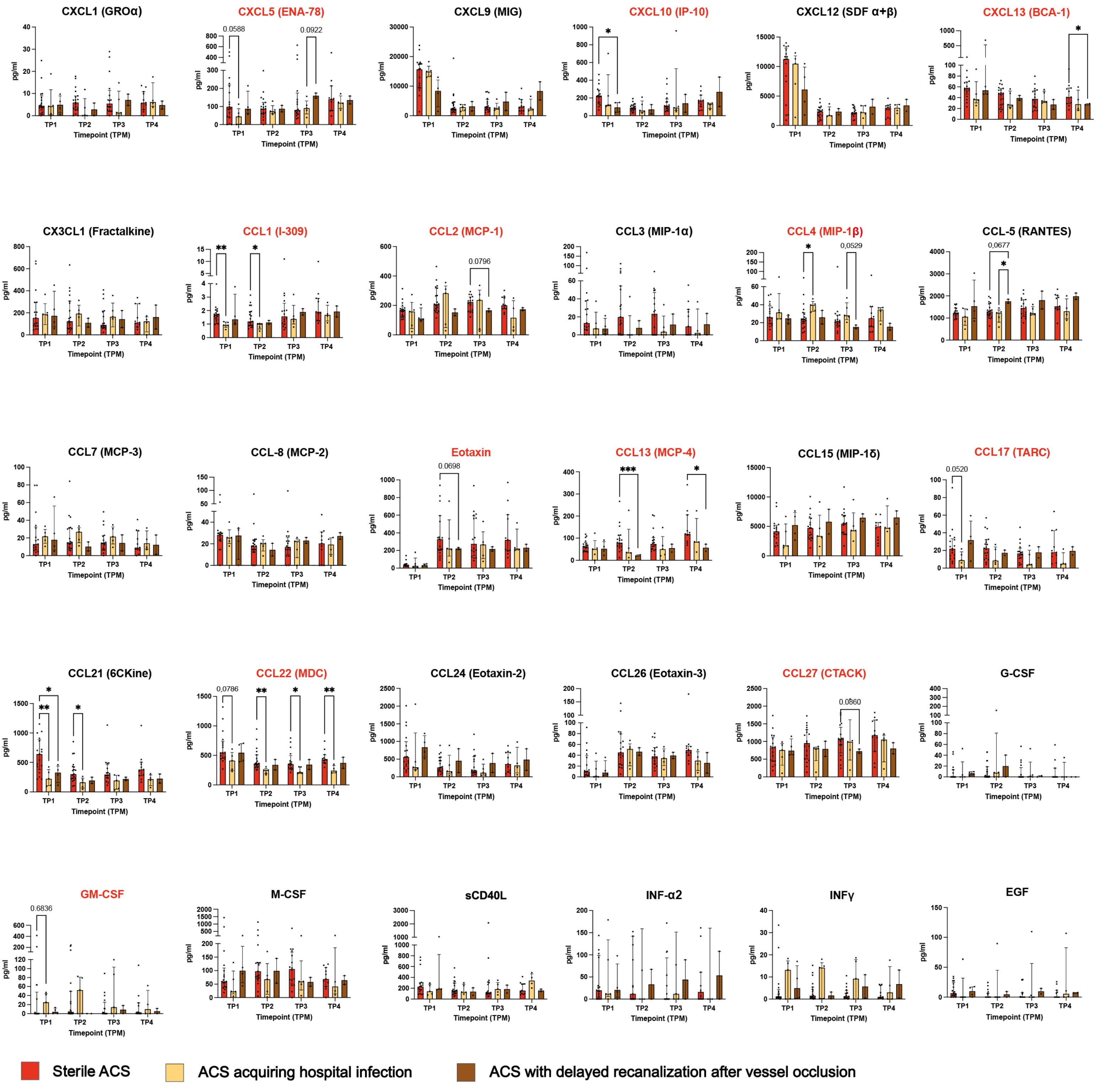


b

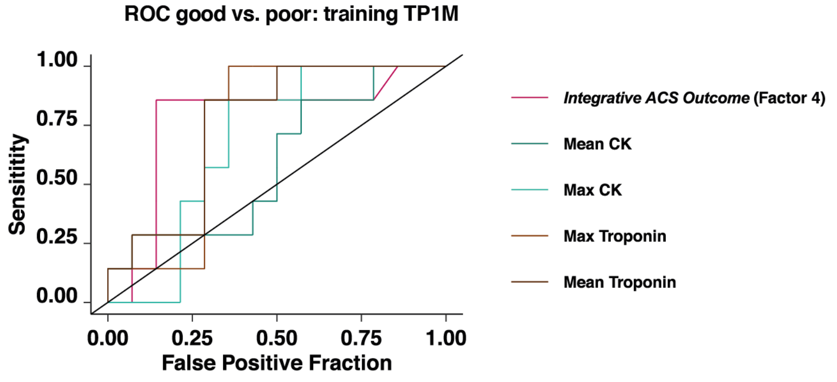

Suppl. Fig. 14

a

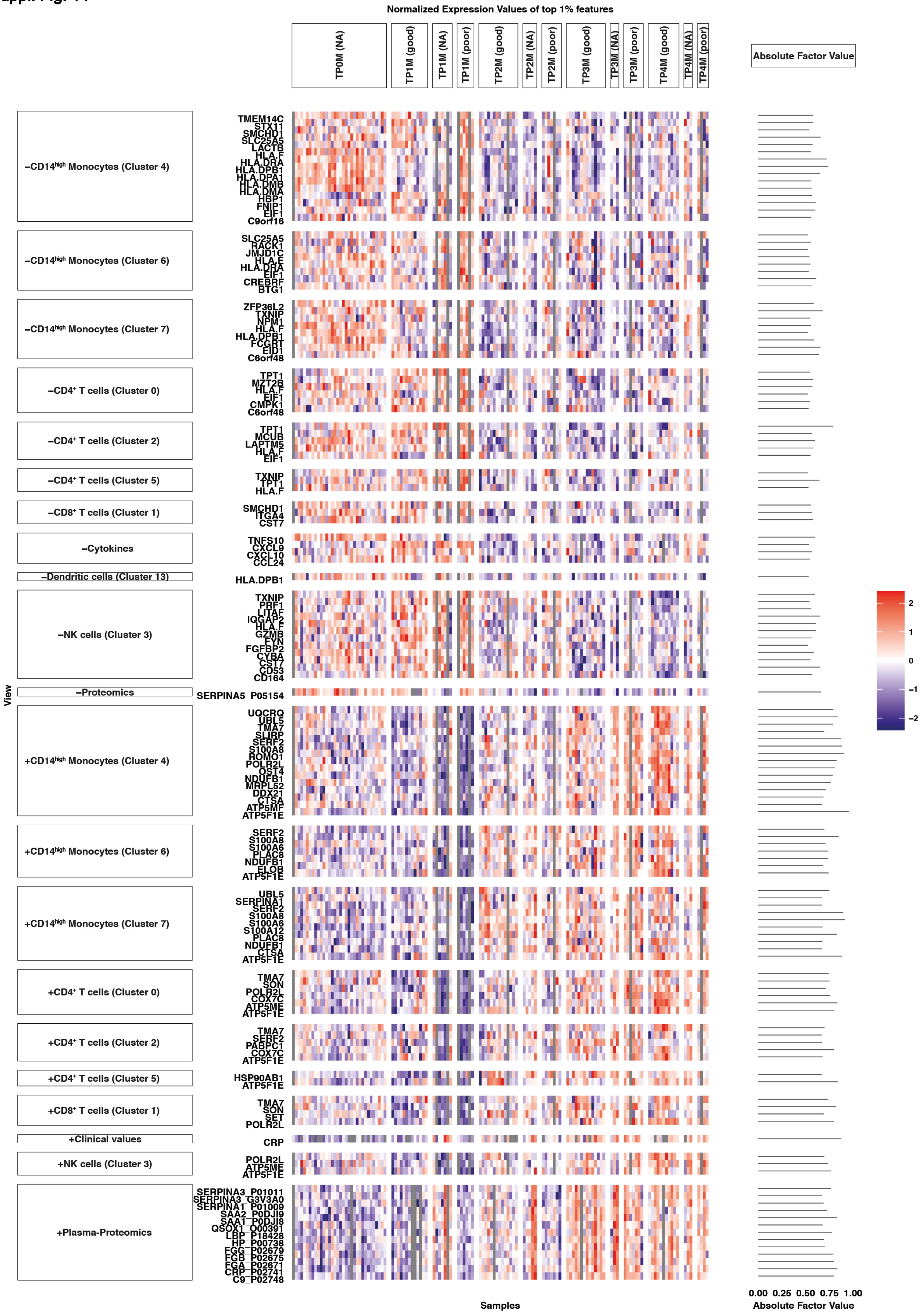

#### Suppl. Fig. 15

**a**

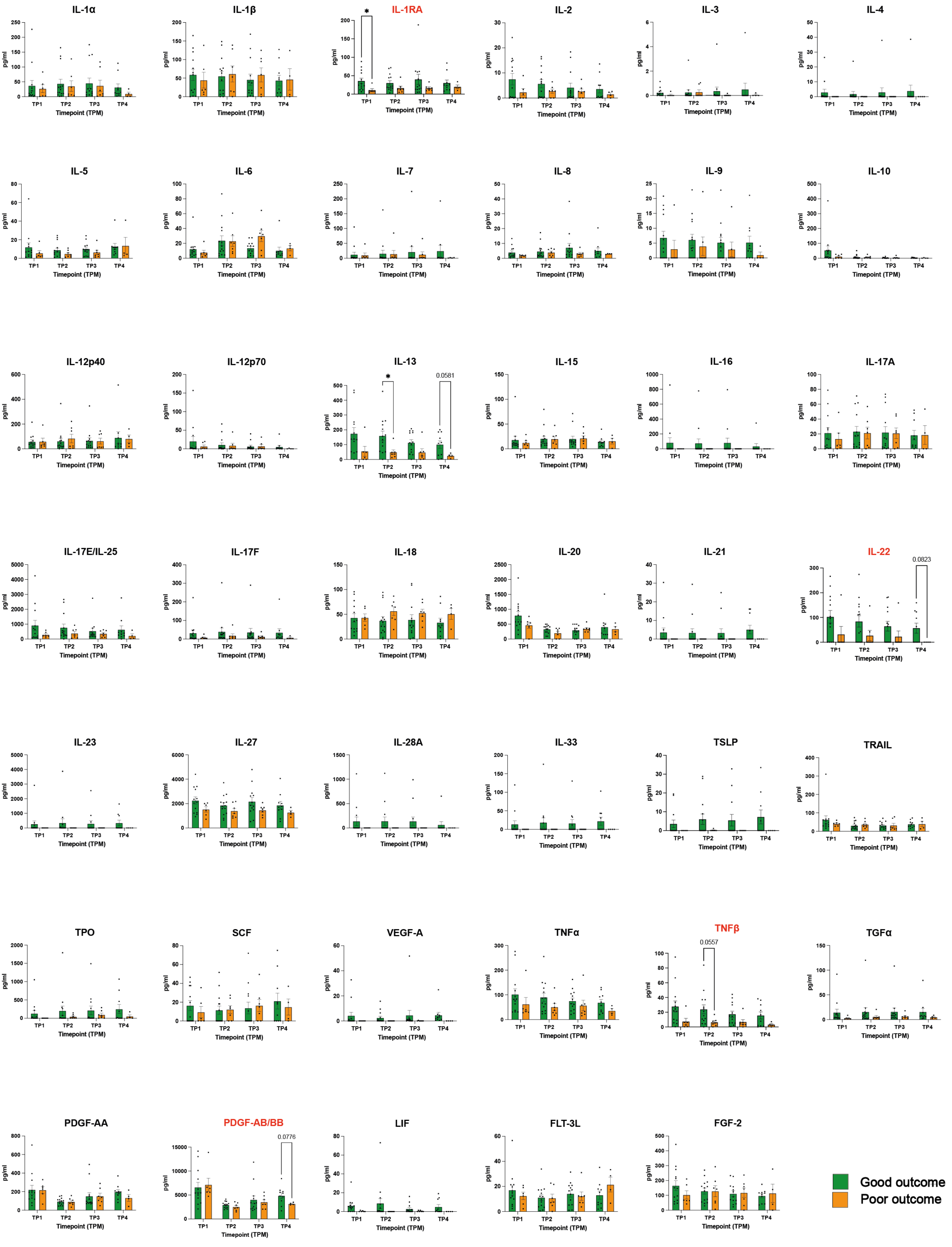

**Suppl. Fig. 16**

**a**

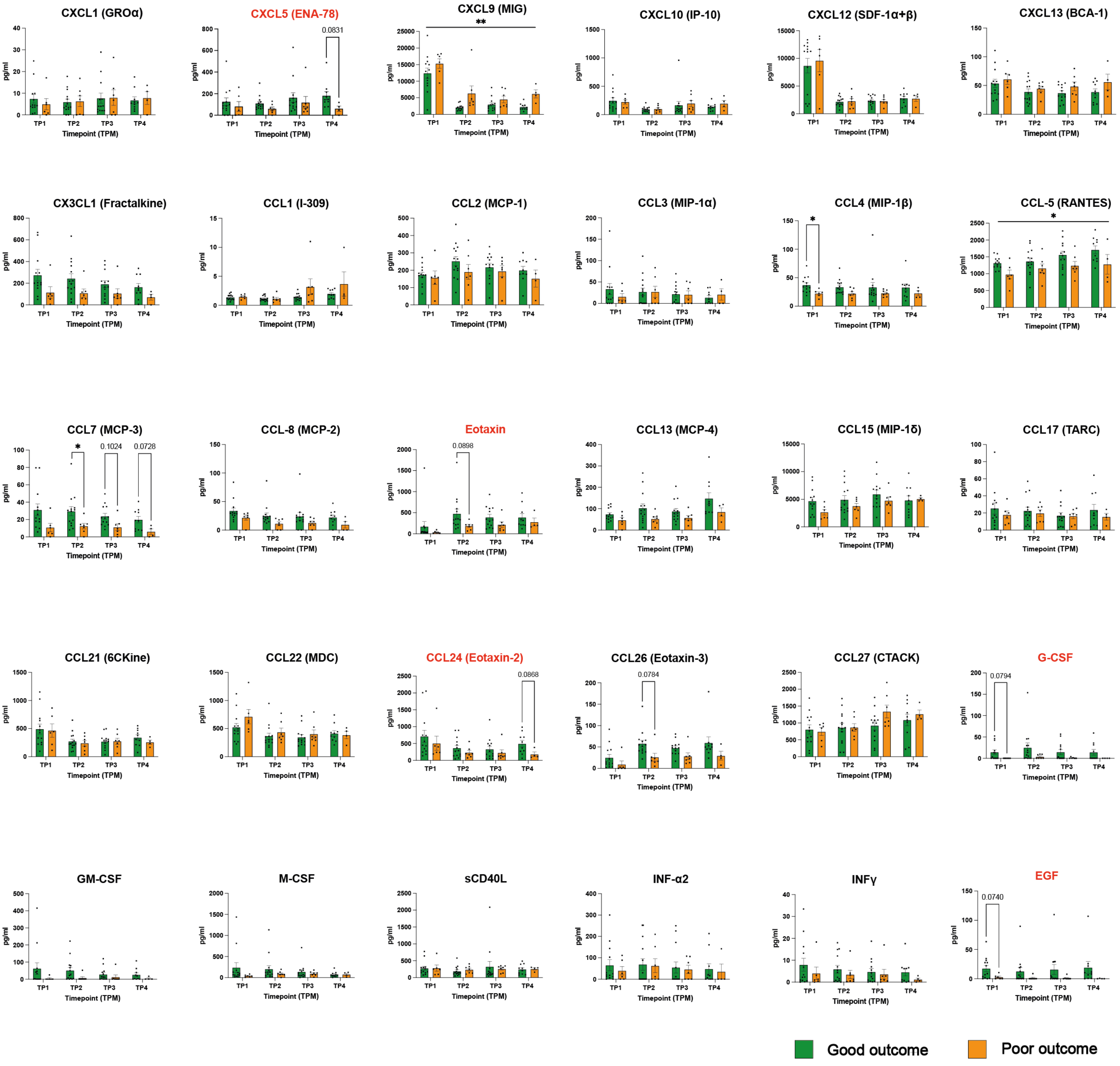

Suppl. Fig. 17

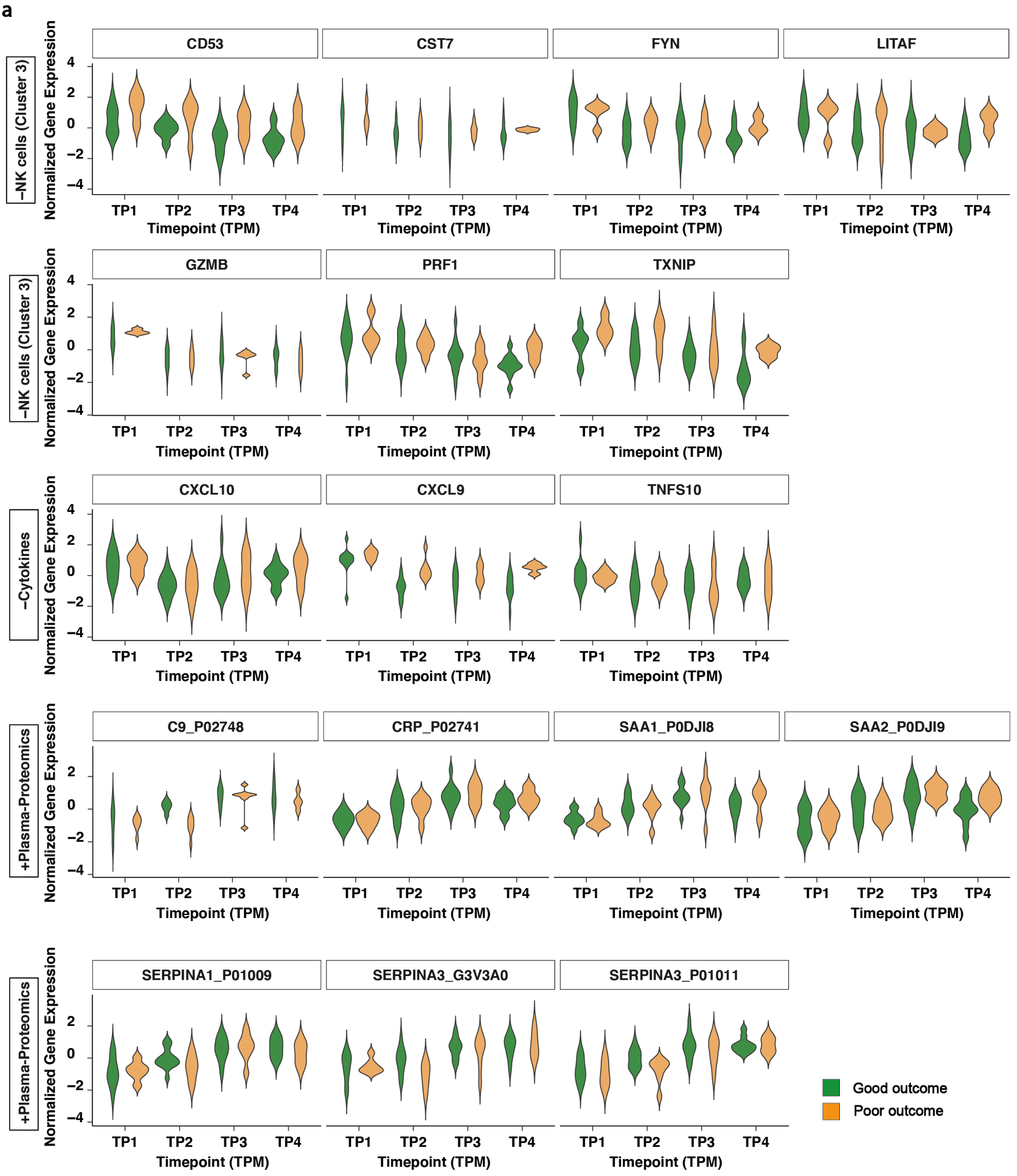
a
–NK cells (Cluster 3)
Normalized Gene Expression
CD53
CST7
FYN
LITAF
GZMB
PRF1
TXNIP
–Cytokines
CXCL10
CXCL9
TNFS10
+Plasma-Proteomics
C9_P02748
CRP_P02741
SAA1_P0DJI8
SAA2_P0DJI9
SERPINA1_P01009
SERPINA3_G3V3A0
SERPINA3_P01011
4
2
0
–2
–4
TP1
TP2
TP3
TP4
Timepoint (TPM)
Good outcome
Poor outcome

Suppl. Fig. 18

a

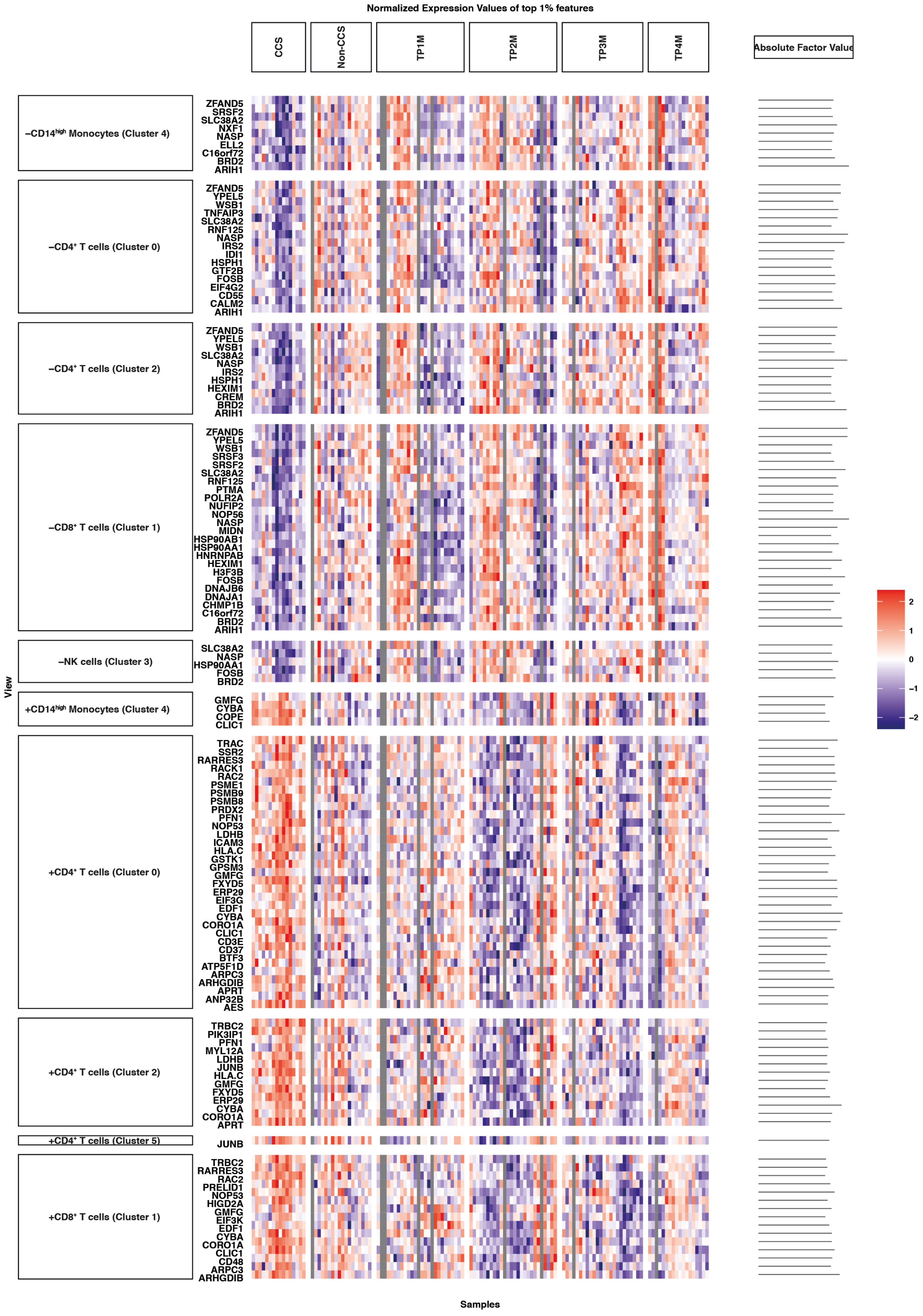

Suppl. Fig. 19

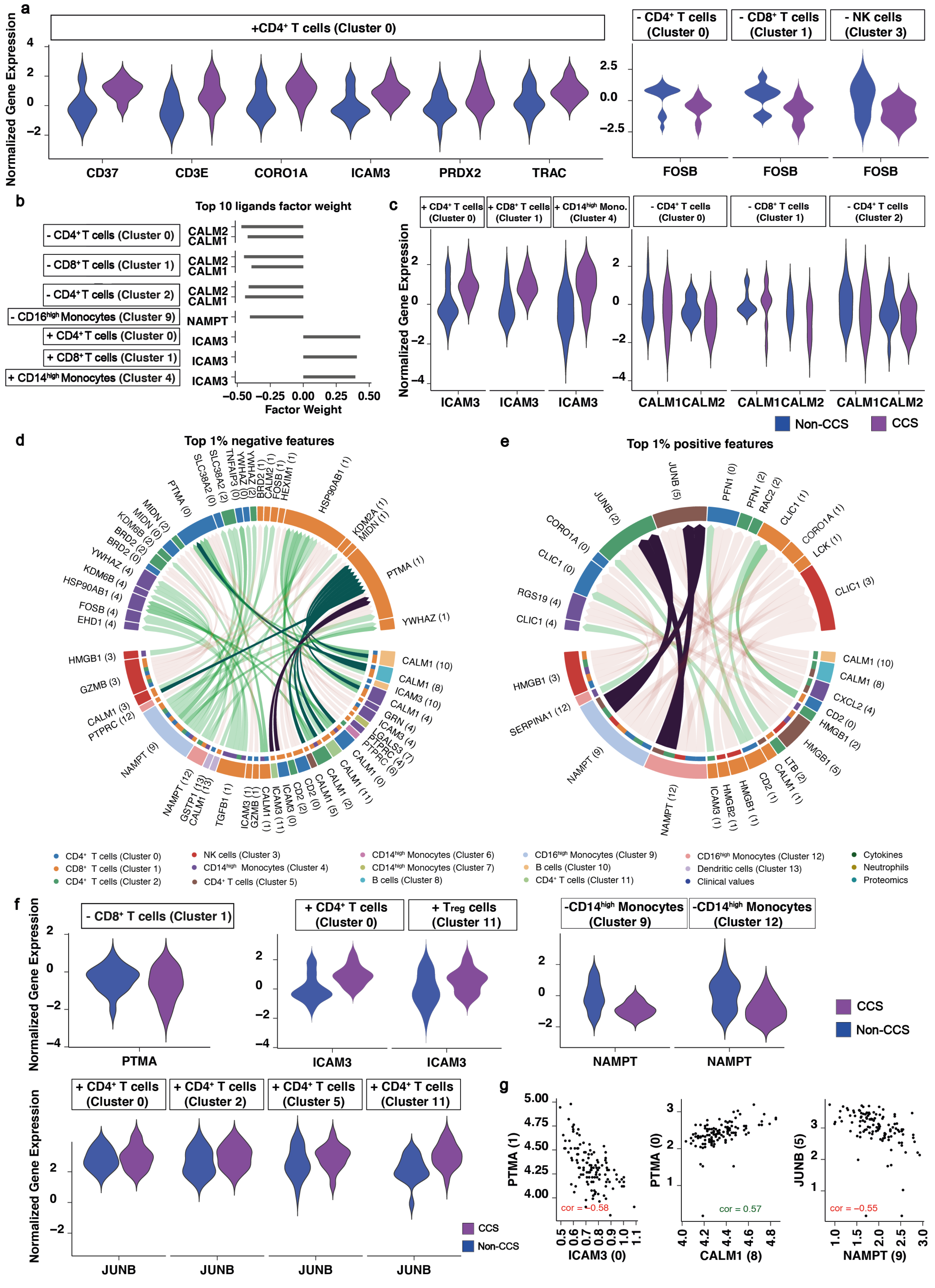

a
+CD4⁺ T cells (Cluster 0)
- CD4⁺ T cells (Cluster 0)
- CD8⁺ T cells (Cluster 1)
- NK cells (Cluster 3)
Normalized Gene Expression
CD37
CD3E
CORO1A
ICAM3
PRDX2
TRAC
FOSB
b
Top 10 ligands factor weight
- CD4⁺ T cells (Cluster 0)
- CD8⁺ T cells (Cluster 1)
- CD4⁺ T cells (Cluster 2)
- CD16high Monocytes (Cluster 9)
+ CD4⁺ T cells (Cluster 0)
+ CD8⁺ T cells (Cluster 1)
+ CD14high Monocytes (Cluster 4)
CALM2
CALM1
NAMPT
ICAM3
Factor Weight
c
+ CD4⁺ T cells (Cluster 0)
+ CD8⁺ T cells (Cluster 1)
+ CD14high Mono. (Cluster 4)
- CD4⁺ T cells (Cluster 0)
- CD8⁺ T cells (Cluster 1)
- CD4⁺ T cells (Cluster 2)
CALM1CALM2
Non-CCS
CCS
d
Top 1% negative features
e
Top 1% positive features
CD4⁺ T cells (Cluster 0)
CD8⁺ T cells (Cluster 1)
CD4⁺ T cells (Cluster 2)
NK cells (Cluster 3)
CD14high Monocytes (Cluster 4)
CD4⁺ T cells (Cluster 5)
CD14high Monocytes (Cluster 6)
CD14high Monocytes (Cluster 7)
B cells (Cluster 8)
CD16high Monocytes (Cluster 9)
B cells (Cluster 10)
CD4⁺ T cells (Cluster 11)
CD16high Monocytes (Cluster 12)
Dendritic cells (Cluster 13)
Clinical values
Cytokines
Neutrophils
Proteomics
f
- CD8⁺ T cells (Cluster 1)
+ CD4⁺ T cells (Cluster 0)
+ Treg cells (Cluster 11)
-CD14high Monocytes (Cluster 9)
-CD14high Monocytes (Cluster 12)
PTMA
ICAM3
NAMPT
+ CD4⁺ T cells (Cluster 0)
+ CD4⁺ T cells (Cluster 2)
+ CD4⁺ T cells (Cluster 5)
+ CD4⁺ T cells (Cluster 11)
JUNB
g
PTMA (1)
ICAM3 (0)
cor = −0.58
PTMA (0)
CALM1 (8)
cor = 0.57
JUNB (5)
NAMPT (9)
cor = −0.55

Suppl. Fig. 20

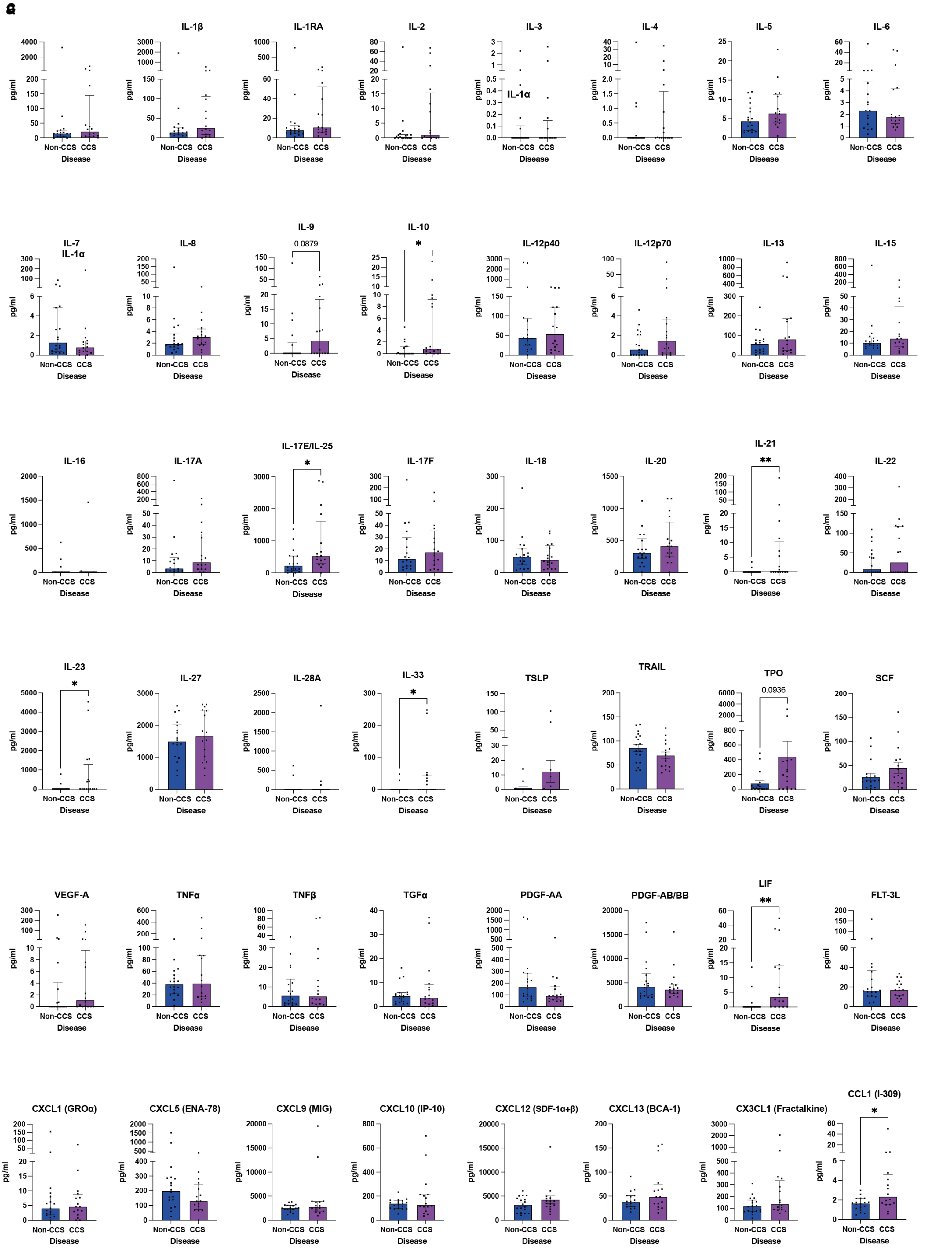
a
IL-1α
IL-1β
IL-1RA
IL-2
IL-3
IL-4
IL-5
IL-6
IL-7
IL-8
IL-9
0.0879
IL-10
*
IL-12p40
IL-12p70
IL-13
IL-15
IL-16
IL-17A
IL-17E/IL-25
*
IL-17F
IL-18
IL-20
IL-21
**
IL-22
IL-23
*
IL-27
IL-28A
IL-33
*
TSLP
TRAIL
TPO
0.0936
SCF
VEGF-A
TNFα
TNFβ
TGFα
PDGF-AA
PDGF-AB/BB
LIF
**
FLT-3L
CXCL1 (GROα)
CXCL5 (ENA-78)
CXCL9 (MIG)
CXCL10 (IP-10)
CXCL12 (SDF-1α+β)
CXCL13 (BCA-1)
CX3CL1 (Fractalkine)
CCL1 (I-309)
*
pg/ml
Non-CCS
CCS
Disease

#### Suppl. Fig. 21

**a**

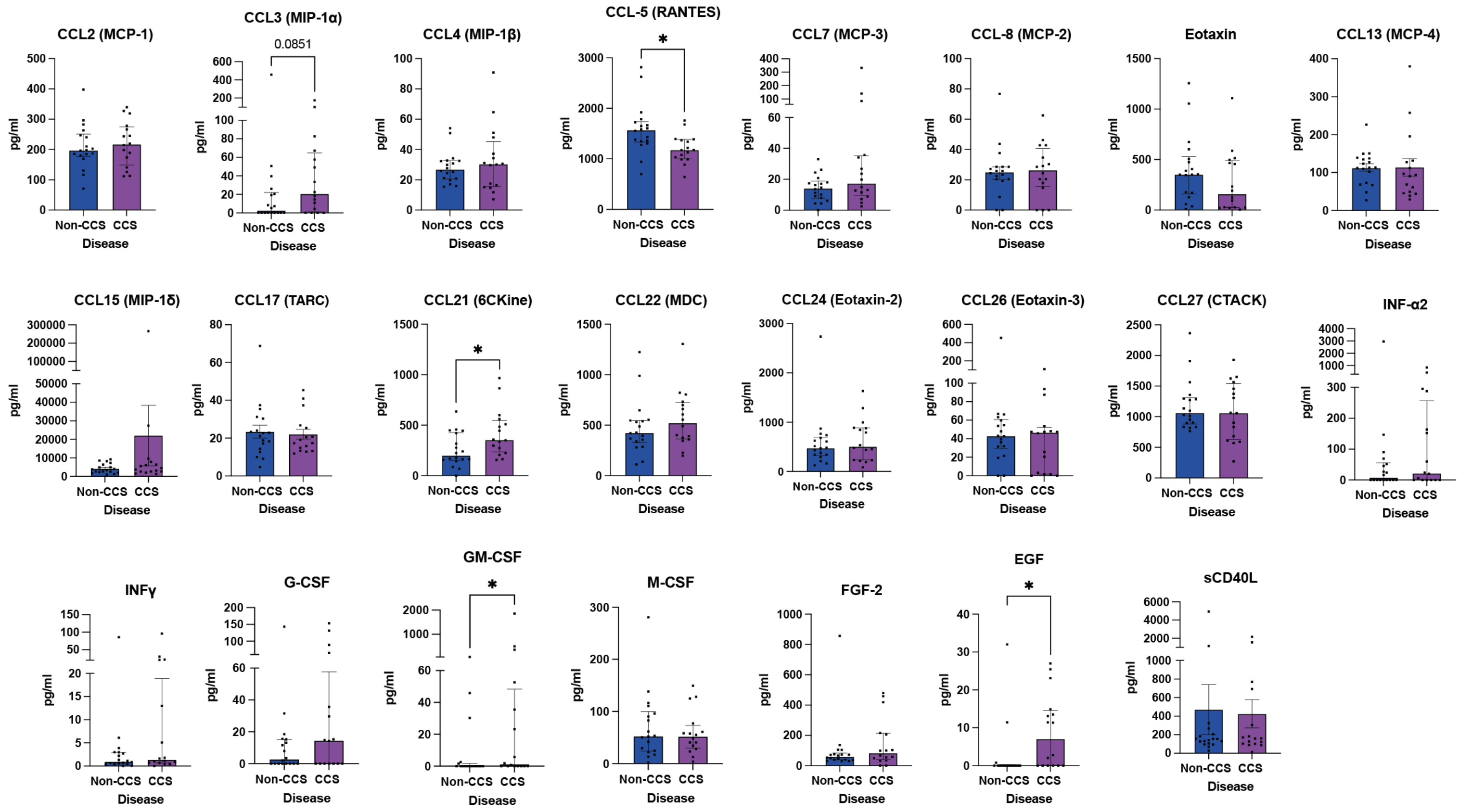
